# Risk prediction tools for pressure injury occurrence: An umbrella review of systematic reviews reporting model development and validation methods

**DOI:** 10.1101/2024.05.07.24306999

**Authors:** Bethany Hillier, Katie Scandrett, April Coombe, Tina Hernandez-Boussard, Ewout Steyerberg, Yemisi Takwoingi, Vladica Velickovic, Jacqueline Dinnes

**Affiliations:** Department of Applied Health Sciences, College of Medicine and Health, University of Birmingham, Edgbaston, Birmingham, UK; NIHR Birmingham Biomedical Research Centre, University Hospitals Birmingham NHS Foundation Trust and University of Birmingham, Birmingham, UK; Department of Medicine, Stanford University, Stanford, CA USA; Department of Biomedical Data Sciences, Leiden University Medical Center, Leiden, The Netherlands; Evidence Generation Department, HARTMANN GROUP, Heidenheim, Germany; Institute of Public Health, Medical, Decision Making and Health Technology Assessment, UMIT, Hall, Tirol, Austria

**Keywords:** Development, internal, external validation, prediction, prognostic, pressure injury, ulcer, overview

## Abstract

**Background:** Pressure injuries (PIs) place a substantial burden on healthcare systems worldwide. Risk stratification of those who are at risk of developing PIs allows preventive interventions to be focused on patients who are at the highest risk. The considerable number of risk assessment scales and prediction models available underscore the need for a thorough evaluation of their development, validation and clinical utility.

Our objectives were to identify and describe available risk prediction tools for PI occurrence, their content and development and validation methods used.

**Methods:** The umbrella review was conducted according to Cochrane guidance. MEDLINE, Embase, CINAHL, EPISTEMONIKOS, Google Scholar and reference lists were searched to identify relevant systematic reviews. Risk of bias was assessed using adapted AMSTAR-2 criteria. Results were described narratively. All included reviews contributed to build a comprehensive list of risk prediction tools.

**Results:** We identified 32 eligible systematic reviews only seven of which described the development and validation of risk prediction tools for PI. Nineteen reviews assessed the prognostic accuracy of the tools and 11 assessed clinical effectiveness. Of the seven reviews reporting model development and validation, six included only machine learning models. Two reviews included external validations of models, although only one review reported any details on external validation methods or results. This was also the only review to report measures of both discrimination and calibration. Five reviews presented measures of discrimination, such as area under the curve (AUC), sensitivities, specificities, F1 scores and G-means. For the four reviews that assessed risk of bias assessment using the PROBAST tool, all models but one were found to be at high or unclear risk of bias.

**Conclusions:** Available tools do not meet current standards for the development or reporting of risk prediction models. The majority of tools have not been externally validated. Standardised and rigorous approaches to risk prediction model development and validation are needed.

**Registration:** The protocol was registered on the Open Science Framework (https://osf.io/tepyk).

## INTRODUCTION

Pressure injuries (PI) carry a significant healthcare burden. A recent meta-analysis estimated the global burden of PIs to be 13%, two-thirds of which are hospital-acquired PIs (HAPI).^1^ The average cost of a HAPI has been estimated as $11k per patient, totalling at least $27 billion a year in the United States based on 2.5 million reported cases.^2^ Length of hospital stay is a large contributing cost, with patients over the age of 75 who develop HAPI having on average a 10-day longer hospital stay compared to those without PI.^3^

PIs result from prolonged pressure, typically on bony areas like heels, ankles, and the coccyx, and are more common in those with limited mobility, including those who are bedridden or wheelchair users. PIs can develop rapidly, and pose a threat in community, hospital and long-term care settings. Multicomponent preventive strategies are needed to reduce PI incidence^4^ with timely implementation to both reduce harm and burden to healthcare systems.^5^ Where preventive measures fail or are not introduced in adequate time, PI treatment involves cleansing, debridement, topical and biophysical agents, biofilms, growth factors and dressings^6^ ^7^ ^8^, and in severe cases, surgery may be necessary.^5^ ^9^

A number of clinical assessment scales for assessing the risk of PI are available (e.g. Braden^10^ ^11^, Norton^12^, Waterlow^13^) but are limited by reliance on subjective clinical judgment. Statistical risk prediction models may offer improved accuracy over clinical assessment scales, however appropriate methods of development and validation are required.^14^ ^15^ ^16^ Although methods for developing risk prediction models have developed considerably,^14^ ^15^ ^17^ ^18^ methodological standards of available models have been shown to remain relatively low.^17^ ^19–22^ Machine learning (ML) algorithms to develop prediction models are increasingly commonplace, but these models are at similarly high risk of bias^23^ and do not necessarily offer any model performance benefit over the use of statistical methods such as logistic regression.^24^ Methods for systematic reviews of risk prediction model studies have also improved,^25–27^ with tools such as PROBAST (Prediction model Risk of Bias Assessment Tool)^28^ now available to allow critical evaluation of study methods.

Although several systematic reviews of PI risk assessment scales and risk prediction models for PI (subsequently referred to as risk prediction tools) are available^29–38^, these have been demonstrated to frequently focus on single or small numbers of scales or models, use variable review methods and show a lack of consensus about the accuracy and clinical effectiveness of available tools.^39^ We conducted an umbrella review of systematic reviews of risk prediction tools for PI to gain further insight into the methods used for tool development and validation, and to summarise the content of available tools.

## METHODS

### Protocol registration and reporting of findings

We followed guidance for conducting umbrella reviews provided in the Cochrane Handbook for Intervention Reviews.^40^ The review was reported in accordance with guidelines for Preferred Reporting Items for Systematic Reviews and Meta-Analyses (PRISMA)^41^ (see Appendix 1), adapted for risk prediction model reviews as required. The protocol was registered on the Open Science Framework (https://osf.io/tepyk).

### Literature search

Electronic searches of MEDLINE, Embase via Ovid and CINAHL Plus EBSCO from inception to June 2024 were developed, tested and conducted by an experienced information specialist (AC), employing well-established systematic review and prognostic search filters^42–44^ combined with specific keyword and controlled vocabulary terms relating to PIs. Additional simplified searches were undertaken in EPISTEMONIKOS and Google Scholar due to the more limited search functionality of these two sources. The reference lists of all publications reporting reviews of prediction tools (systematic or non-systematic) were reviewed to identify additional eligible systematic reviews and to populate a list of PI risk prediction tools. Title and abstract screening and full text screening were conducted independently and in duplicate by two of four reviewers (BH, JD, YT, KS). Any disagreements were resolved by discussion or referral to a third reviewer.

### Eligibility criteria for this umbrella review

Published English-language systematic reviews of risk prediction models developed for adult patients at risk of PI in any setting were included. Reviews of clinical risk assessment tools or models developed using statistical or ML methods were included, both with or without internal or external validation. The use of any PI classification system^6^ ^45–47^ as a reference standard was eligible. Reviews of the diagnosis or staging of those with suspected or existing PIs or chronic wounds, reviews of prognostic factor and predictor finding studies, and models exclusively using pressure sensor data were excluded.

Systematic reviews were required to report a comprehensive search of at least two electronic databases, and at least one other indicator of systematic methods (i.e. explicit eligibility criteria, formal quality assessment of included studies, sufficient data presented to allow results to be reproduced, or review stages (e.g. search screening) conducted independently in duplicate).

### Data extraction and quality assessment

Data extraction forms (Appendix 3) were developed using the CHARMS checklist (CHecklist for critical Appraisal and data extraction for systematic Reviews of prediction Modelling Studies) and Cochrane Prognosis group template.^48^ ^49^ One reviewer extracted data concerning: review characteristics, model details, number of studies and participants, study quality and results. Extractions were independently checked by a second reviewer. Where discrepancies in model or primary study details were noted between reviews, we accessed the primary model development publications where possible.

The methodological quality of included systematic reviews was assessed using AMSTAR-2 (A Measurement Tool to Assess Systematic Reviews)^50^, adapted for systematic reviews of risk prediction models (Appendix 4). Quality assessment and data extraction were conducted by one reviewer and checked by a second (BH, JD, KS), with disagreements resolved by consensus. Our adapted AMSTAR-2 contains six critical items, and limitations in any of these items reduce the overall validity of a review.^50^

### Synthesis methods

Reviews were considered according to whether any information concerning model development and validation was reported. This specifically refers to reporting methods of model development or validation, and/or the presentation of measures of both discrimination and calibration. This is in contrast to evaluations of prognostic accuracy, where models are applied at a binary threshold (e.g., for high or low risk), and present only discrimination metrics with no further consideration of model performance. Available data were tabulated, and a narrative synthesis provided.

All risk prediction models identified are listed in Appendix 5 Table S4, including those for which no information about model development or validation was provided at systematic review level. Risk prediction models were classified as ML-based or non-ML models, based on how they were classified in included systematic reviews, including cases where models such as logistic regression were treated as ML-based models. Where possible, the predictors included in the tools were extracted at review level and categorised into relevant groups in order to describe the candidate predictors associated with risk of PI. No statistical synthesis of systematic review results was conducted.

Reviews reporting results as prognostic accuracy (i.e. risk classification according to a binary decision) or clinical effectiveness (i.e. impact on patient management and outcomes) are reported elsewhere.^39^ Hereafter, the term clinical utility is used to encompass both accuracy and clinical effectiveness.

## RESULTS

### Characteristics of included reviews

Following de-duplication of search results, 7200 unique records remained, of which 118 were selected for full text assessment. We obtained the full text of 111 publications of which 32 met all eligibility criteria for inclusion (see Figure 1). Seven reviews reported details about model development and internal validation^36^ ^37^ ^51–55^, two of which also considered external validation^52^ ^54^; 19 reported accuracy data^29^ ^31–35^ ^38^ ^54^ ^56–66^; and 11 reported clinical effectiveness data.^30^ ^56^ ^58^ ^61^ ^66–72^ One review^54^ reported both model development and accuracy data, and four reviews reported both accuracy and effectiveness data.^56^ ^58^ ^61^ ^66^

**Figure 1.**
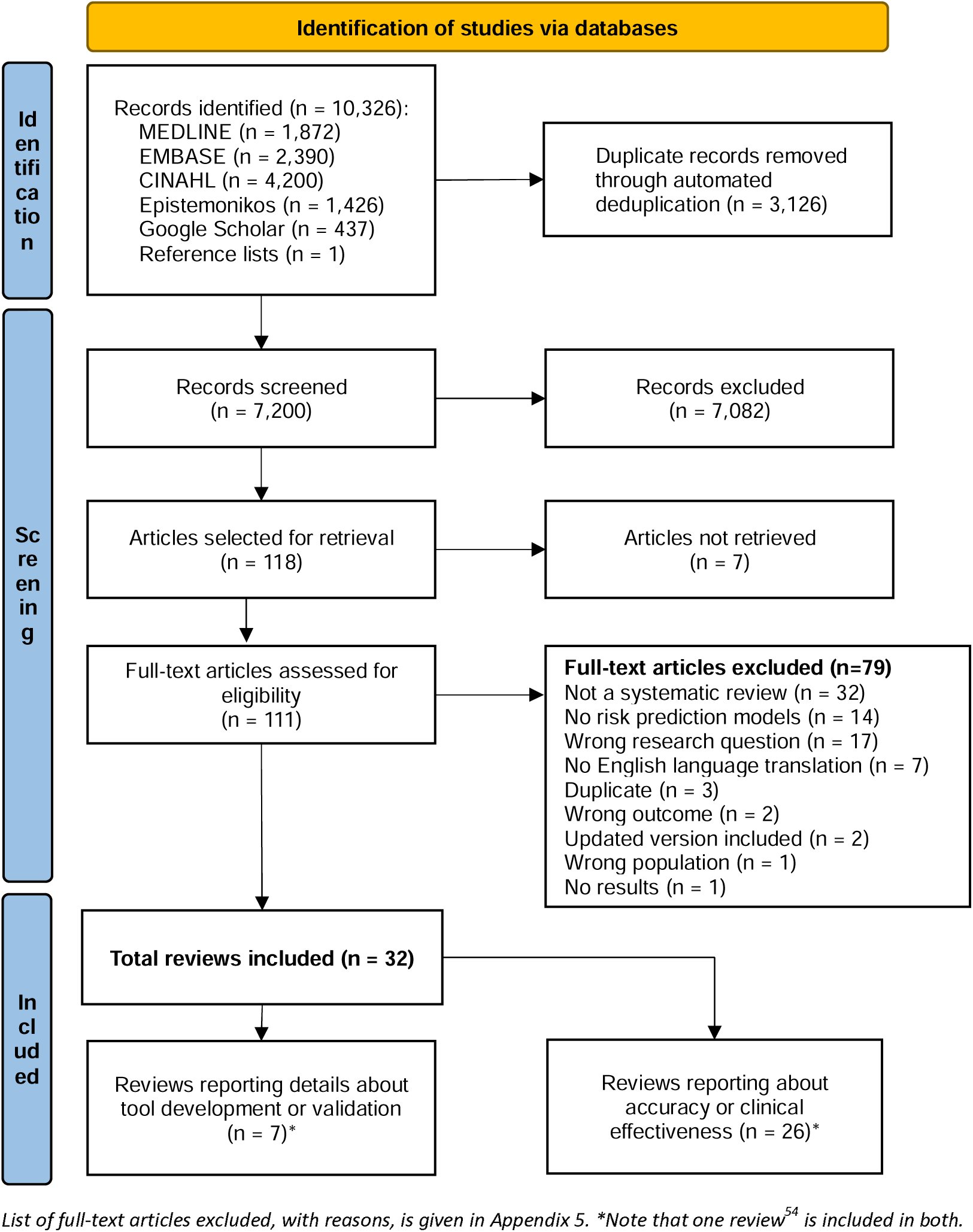
PRISMA^41^ flowchart: identification, screening and selection process

Table 1 provides a summary of systematic review methods for all 32 reviews according to whether or not they reported any tool development methods (see Appendix 5 for full details). The seven reviews reporting prediction tool development and validation were all published within the last six years (2019 to 2024) compared to reviews focused on the clinical utility of available tools (published from 2006 to 2024). Reviews focused on model development methods almost exclusively focused on ML-based models (all but one^60^ of the seven reviews limited inclusion to ML models), and frequently did not report study eligibility criteria related to study participants or setting (Table 1). In comparison, only two reviews (8%) concerning the clinical utility of models included ML-based models,^38^ ^54^ but more often reported eligibility criteria for population or setting: hospital settings (n = 3),^33^ ^38^ ^54^ or surgical settings (n=8),^34^ ^61^ ^63^ ^64^ ^70^ ^31^, hospital or acute settings (n=2)^67^ ^71^, long-term care settings (n=2)^29^ ^35^ or the elderly (n=1).^60^

**Table 1.**
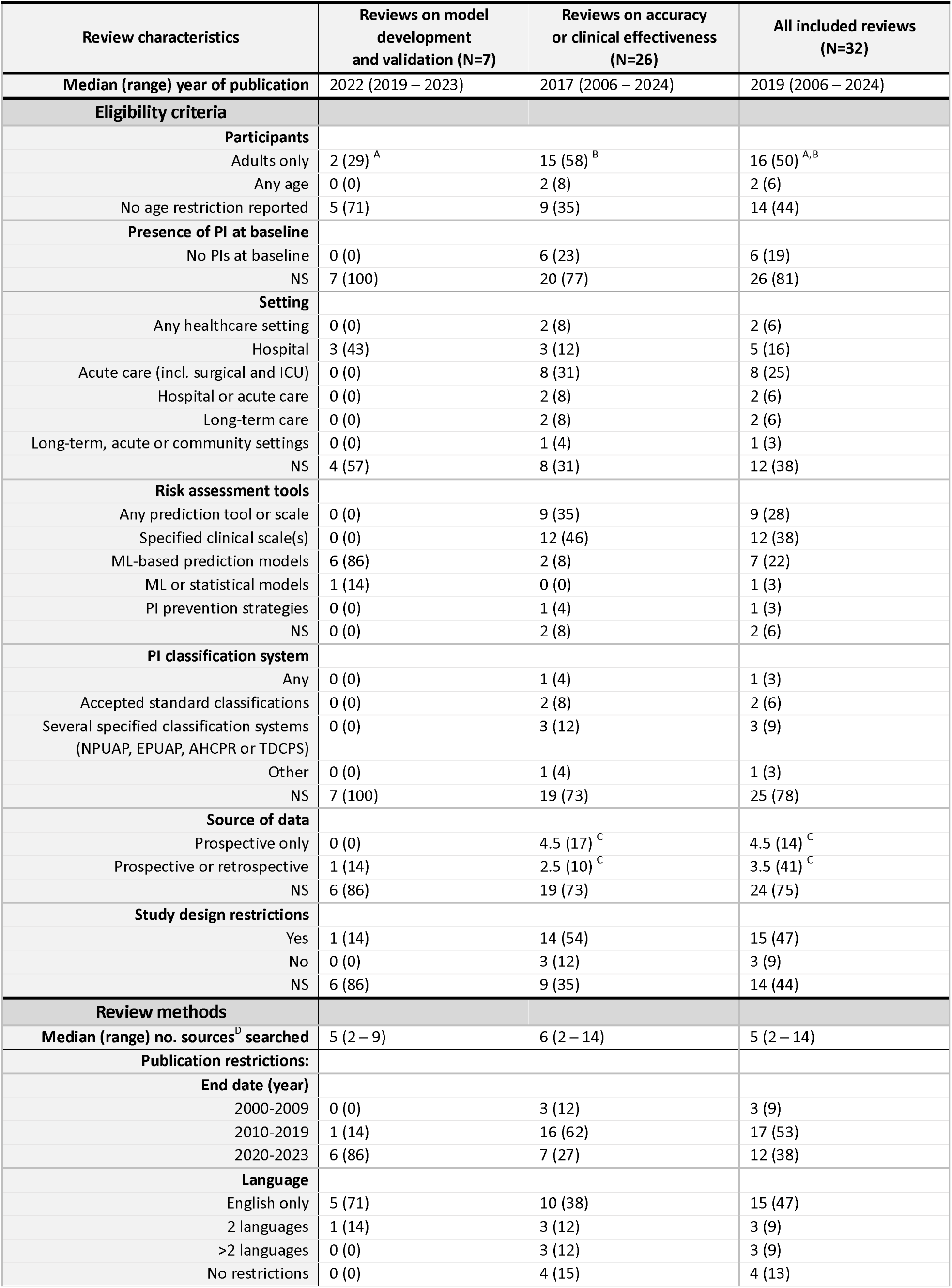
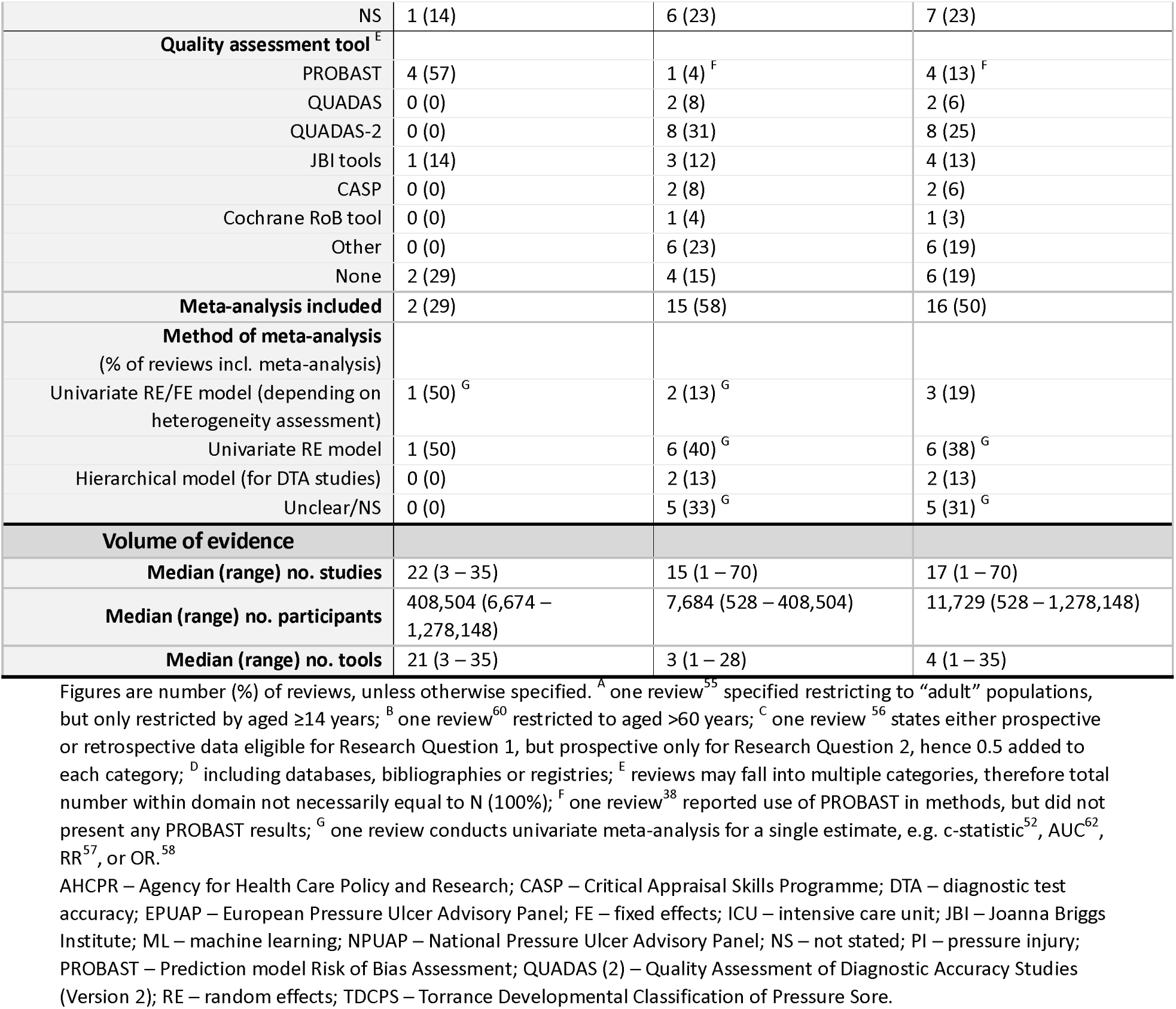
Summary of included systematic review characteristics.

On average, reviews about tool development included more studies than reviews of clinical utility (median 22 compared to 15), more participants (median 408,504 compared to 7,684) and covered more prediction tools (median 21 compared to 3) (Table 1). Ten reviews (38%) about clinical utility included only one risk assessment scale, whereas reviews of tool development included at least 3 different risk prediction models. The PROBAST tool for quality assessment of prediction model studies was used in 57% (n=4) of tool development reviews^37^ ^52–54^, whereas validated test-accuracy specific tools such as QUADAS were used less frequently (10/26, 38%) in reviews of clinical utility. Two reviews of tool development did not report any quality assessment of included studies (29%), compared to 4 (15%) of reviews of clinical utility. Meta-analysis was conducted in two of seven (29%) reviews of tool development compared to more than half of reviews of clinical utility (15, 58%).

### Methodological quality of included reviews

The quality of included reviews was generally low (Table 2; Appendix 5 for full assessments). The majority of reviews (71% (5/7) reviews on tool development and 78% (18/23) reviews on clinical utility) partially met the AMSTAR-2 criteria for the literature search (i.e. searched two databases, reported search strategy or key words, and justified language/publication restrictions), with only three (two reviews^56^ ^72^ on clinical utility, and one review^54^ on both tool development and clinical utility) meeting all criteria for ‘Yes’ (i.e. searching grey literature and reference lists, with the search conducted within 2 years of publication). Twenty-two reviews (69%) conducted study selection in duplicate (5/7 (71%) of reviews about tool development and 17/26 (65%) of clinical utility reviews). Conflicts of interest were reported in all seven tool development reviews and 77% of clinical utility reviews (20/26). Reviews scored poorly on the remaining AMSTAR-2 items, with around 50% or fewer reviews meeting the stipulated AMSTAR-2 criteria. Nine reviews (28%) used an appropriate method of quality assessment of included studies and provided itemisation of judgements per study. No review scored ‘Yes’ for all AMSTAR-2 items in either category.

**Table 2.**
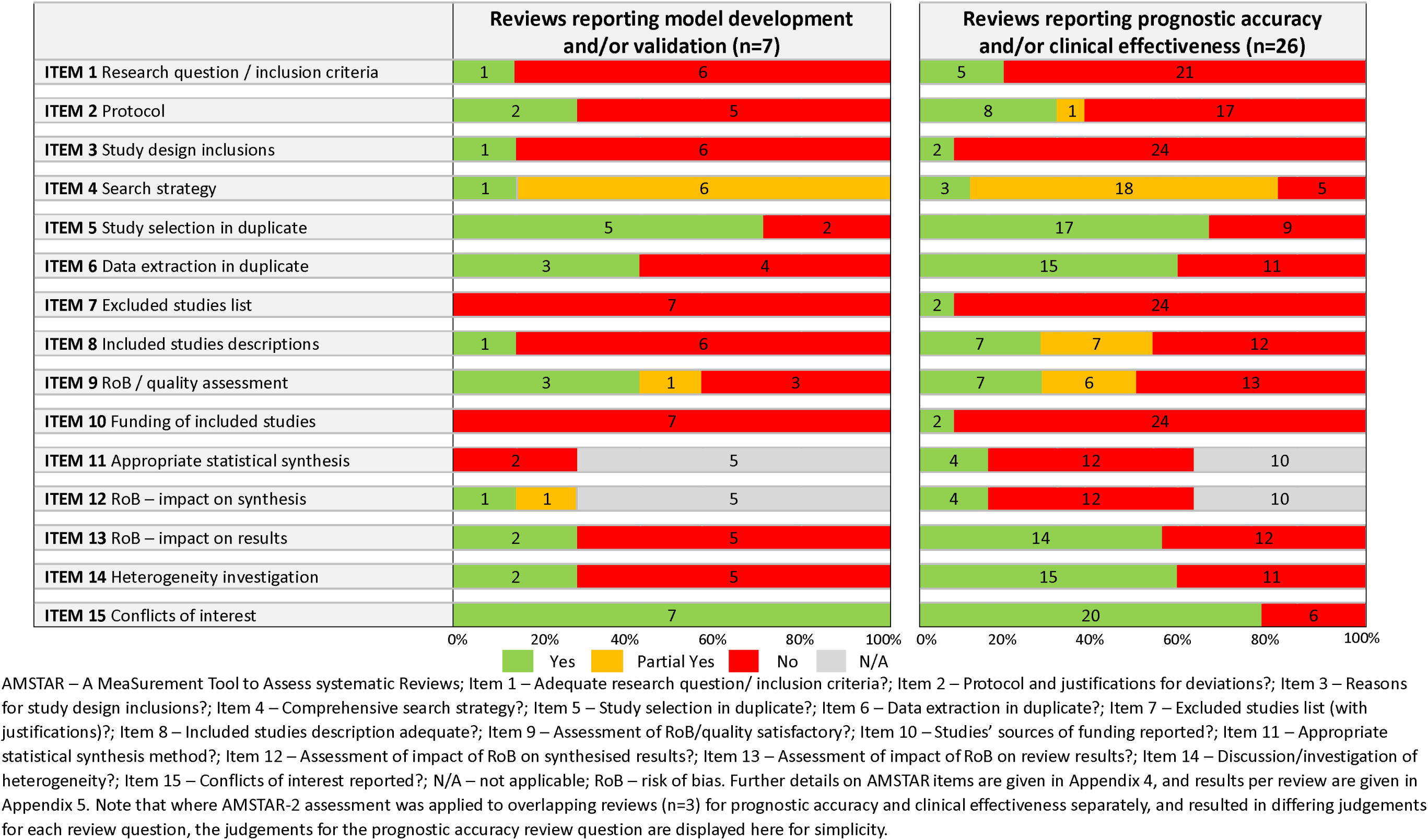
Summary of AMSTAR-2 assessment results.

### Findings

Of the 32 reviews, 26 reviews focused on the clinical utility (accuracy or effectiveness) of prediction tools. These clinical utility reviews provided no details about the development or validation of included models (except for one review^54^), and gave only limited detail about setting and study design (see Appendix 5). Reviews reporting the accuracy of prediction tools largely treated the tools as diagnostic tests to be applied at a single threshold (e.g., for high or low risk) and they did not focus on the broader aspects of prognostic model performance, such as calibration and the temporal relationship between prediction and the outcome, PI occurrence. These reviews included a total of 70 different prediction tools, predominantly derived by clinical experts, as opposed to empirically-derived models (that is, with statistical or ML methods). The methodology underlying their development is not always explicit, with scales in routine clinical usage apparently based on epidemiological evidence and clinical judgment about predictors that may not meet accepted principles for the development and reporting of risk prediction models. The most commonly included tools were the Braden^10^ ^11^ (included in 21 reviews), Waterlow^13^ (n=14 reviews), Norton^12^ (n=11 reviews), and Cubbin and Jackson scales^97^ ^98^ (n=8 reviews).

The seven systematic reviews that reported detailed information about model development and validation included 70 prediction models, 48 of which were unique to these seven reviews. Between three^51^ and 35^36^ model development studies were included; one review^52^ also included eight external validation studies and another review^54^ included one external validation study. Electronic health records (EHRs) were used for model development in all studies in one review^37^ and for the majority of models (>66%) in the remaining reviews, where reported.^51^ ^54^ ^55^ ^53^ Three reviews^52^ ^54^ ^55^ reported the use of prospectively or retrospectively collected data. No review included information about the thresholds used define whether a patient is at risk of developing PIs. Five reviews included detail about the predictors included in each model.

The largest review^36^ reported that logistic regression was the most commonly reported modelling approach (20/35 models), followed by random forest (n=18), decision tree (n=12) and support vector machine (n=12) approaches. Logistic regression was also the most frequently used approach in three other reviews (18/23^55^, 16/21^52^ and 15/22^53^). Primary studies frequently compared the use of different ML methods using the same datasets, such that ‘other’ ML methods were reported with little to no further detail (e.g. 19 studies in the review by Dweekat and colleagues^36^).

Approaches to internal validation were not well reported in the primary studies. One review^52^ found no information on internal validation for 76% (16/21) of studies; with re-sampling reported in two and tree-pruning, cross-validation and split sample reported in one study each. Another review^36^ reported finding no information about internal validation for 20% of studies (7/35) and the use of cross-validation (n=10), split sample (n=10) techniques, or both (n=8) for the remainder. Cross-validation was used in more than half (12/22) of studies in another.^53^

Only one review reported details on methods for selection of model predictors^52^: 29% (6/21) selected predictors by univariate analysis prior to modelling and 9 used stepwise selection for final model predictors; 11 (52%) clearly reported candidate predictors, and all 21 clearly reported final model predictors. Another review^54^ stated that feature selection (or predictor selection) was performed improperly and that some studies used univariate analyses to select predictors, but further details were not provided. One review^52^ reported 15 models (71%) with no information about missing data, and only two using imputation techniques (imputation using another data set, and multiple imputation by chained equations). Another review^54^ reported 7 models (39%) with no information about missing data, missing data excluded or negligible for 4 models (22%), and single or multiple imputation techniques used for 5 (28%) and 3 (17%) models, respectively.

Model performance measures were reported by three reviews^37^ ^52^ ^53^, all of which noted considerable variation in reported metrics and model performance including C-statistics (0.71 to 0.89 in 10 studies^53^), F1 score (0.02 to 0.99 in 9 studies^53^), G-means (0.628 to 0.822 in four studies^37^), and observed versus expected ratios (0.97 to 1 in 3 studies^52^). Four reviews^37^ ^53–55^ reported measures of discrimination associated with included models. Across reviews, reported sensitivities ranged between 0.04 and 1, specificities ranged between 0.69 and 1, and AUC values ranged between 0.50 and 1.

Shi and colleagues^52^ included eight external validations using data from long-term care (n=4) or acute hospital care (n=4) settings (Appendix 5 Table S5). All were judged to be at unclear (n=4) or high (n=4) risk of bias using PROBAST. Model performance metrics for five models (TNH-PUPP^89^, Berlowitz 11-item model^99^, Berlowitz MDS adjustment model^90^, interRAI PURS^88^, Compton ICU model^94^) included C-statistics between 0.61 and 0.9 and reported observed versus expected ratios were between 0.91 and 0.97. The review also reported external validation studies for the ‘SS scale’^100^ and the prePURSE study tool^91^, but no model performance metrics were given. A meta-analysis of C-statistics and O/E ratios was performed, including values from both development and external validation cohorts (Table 3). Parameters related to model development were not consistently reported: C-statistics ranged between 0.71 and 0.89 (n = 10 studies); observed versus expected ratios ranged between 0.97 and 1 (n=3 studies).

**Table 3.**
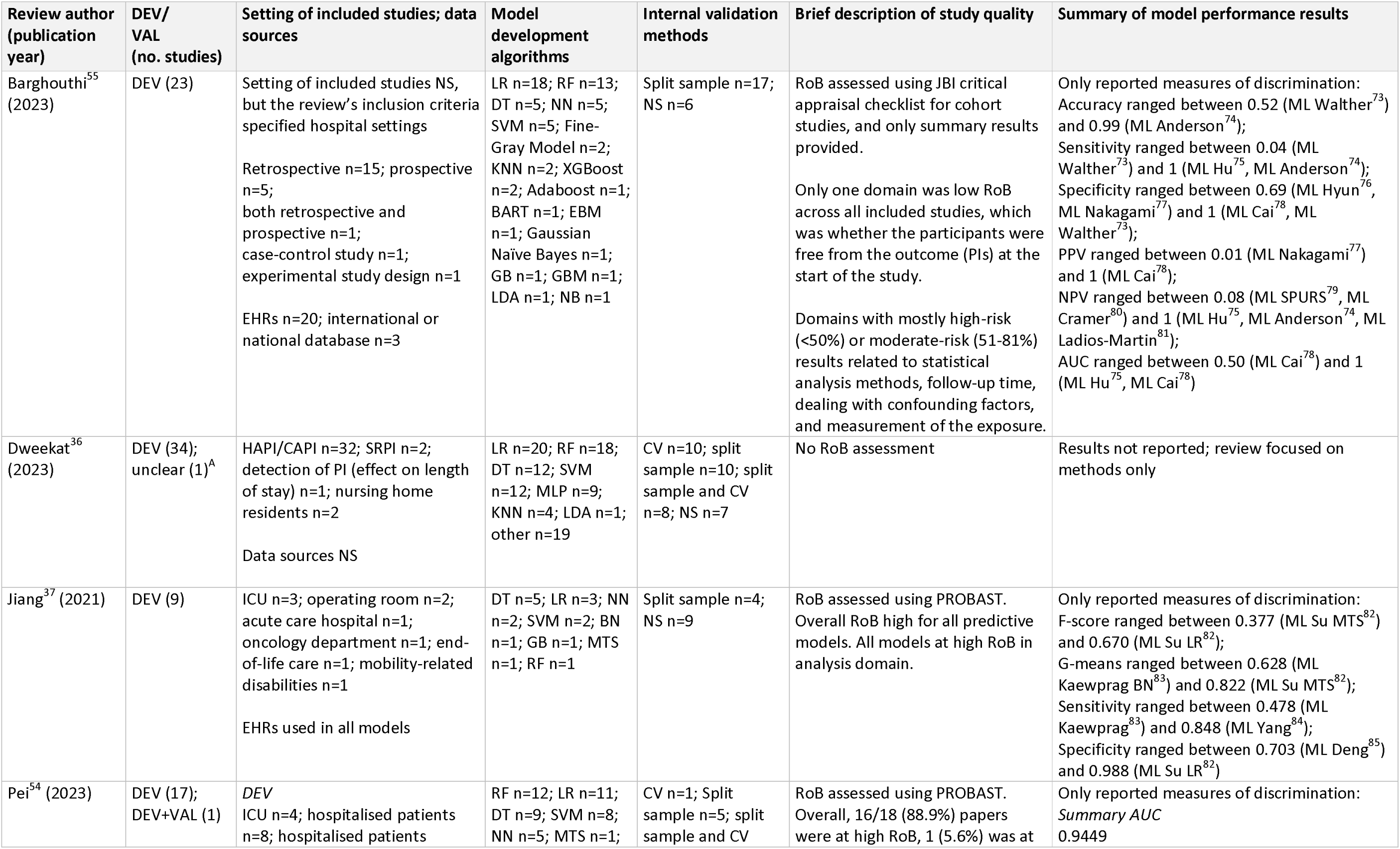
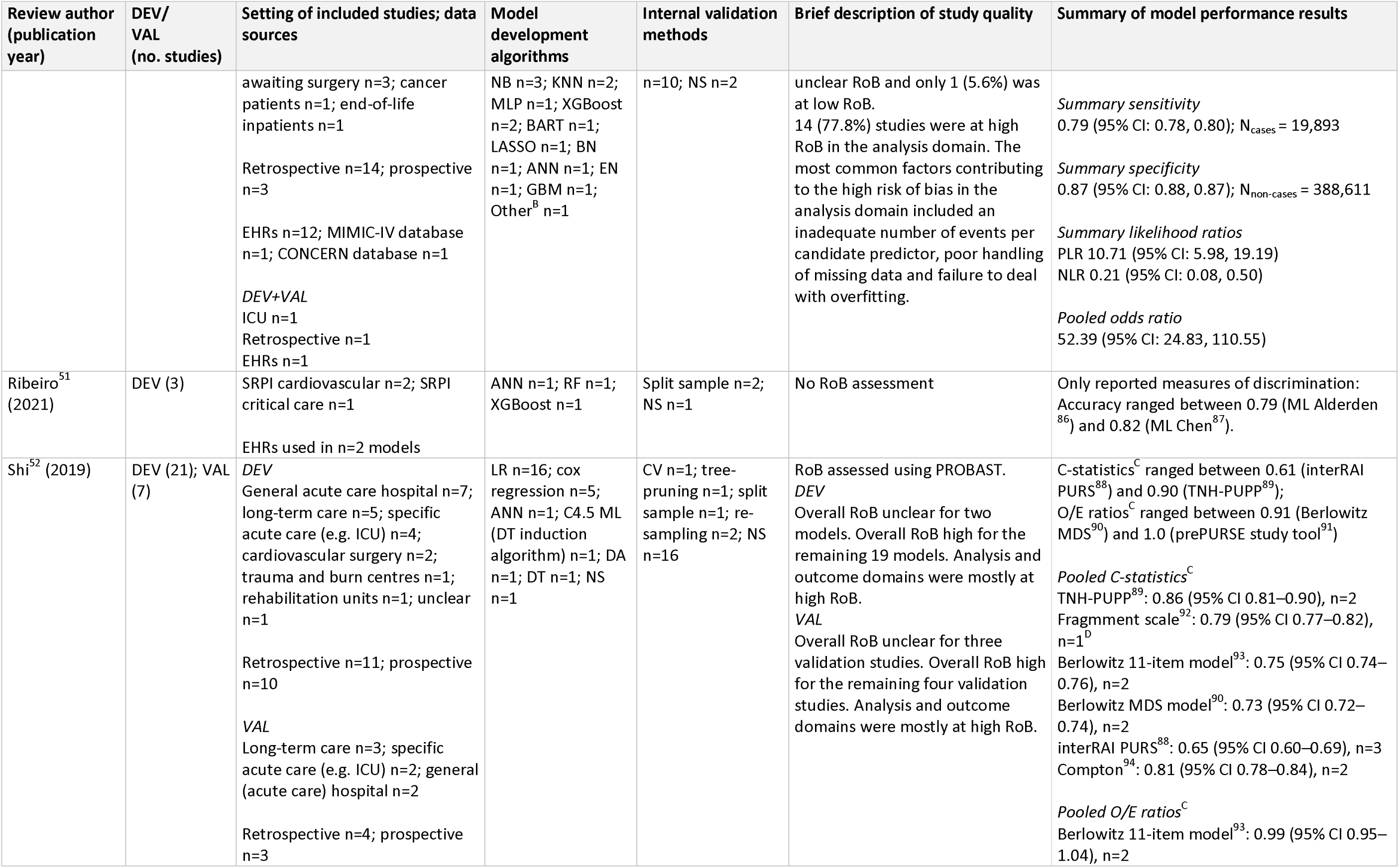
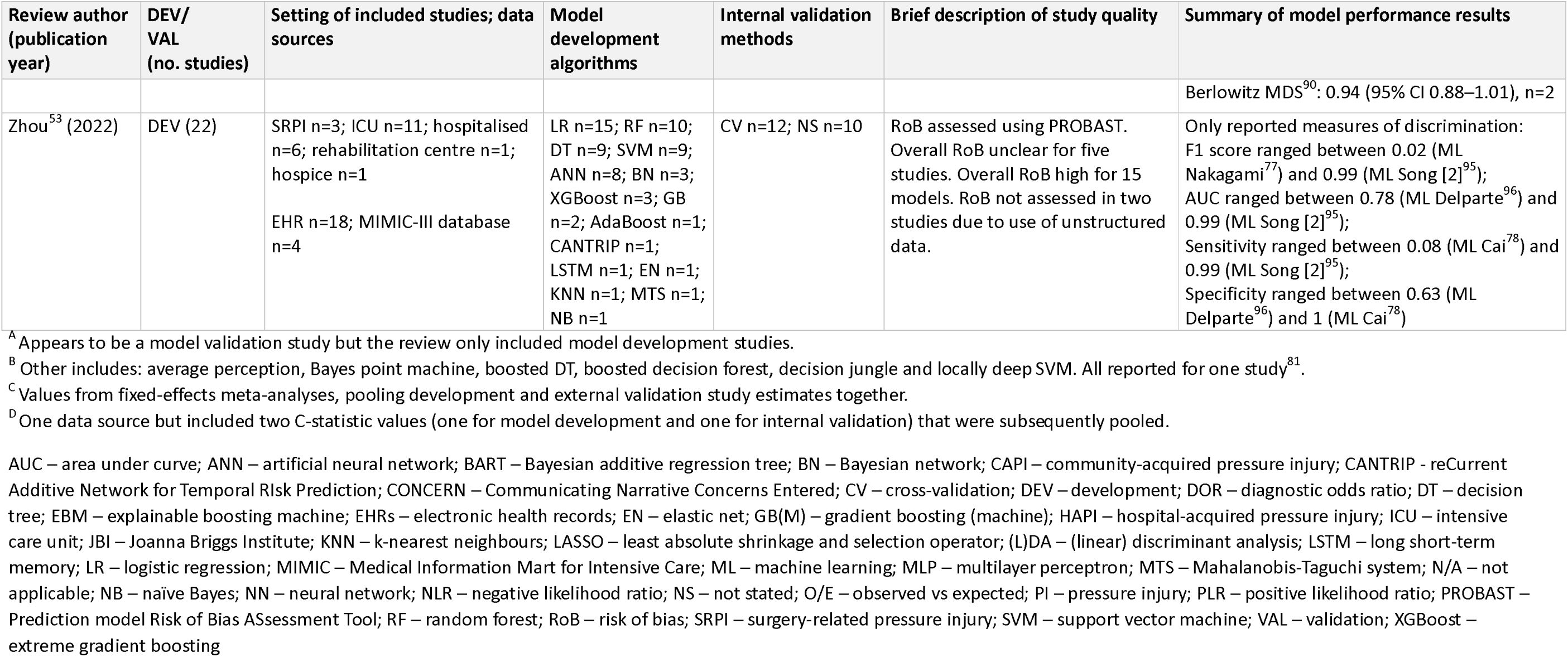
Results of reviews reporting model development and validation.

Pei and colleagues^54^ reported that one^81^ (1/18, 6%) of the model development studies included in their review also conducted an external validation. However, review authors presented accuracy metrics that originated from the internal validation, as opposed to the external validation (determined from inspection of the primary study). Additionally, no details on external validation methods and no measures of calibration were presented. Pei and colleagues^54^ judged this study to be of high risk of bias using PROBAST, as with the majority of studies (16/18, 89%) included in their review. More detailed information about individual models, including predictors, specific model performance metrics and sample sizes, is presented in Appendix 5.

### Included tools and predictors

A total of 124 risk prediction tools were identified (Table 4); 111 tools were identified from the 32 included systematic reviews and 13 were identified from screening the reference lists of literature reviews that used non-systematic methods that were considered during full text assessment. Full details obtained at review-level are reported in Appendix 5 Table S4.

**Table 4.** Summary of tool characteristics, extracted at review-level.

| Tool characteristics | ML-based models<br>(N=60, 48%) | Non-ML tools<br>(N=64, 52%) | Total<br>(N=124) |
| --- | --- | --- | --- |
| **No. of included reviews<sup>A</sup> considered in** |  |  |  |
| 0 | 0 (0) | 13 (20) | 13 (10) |
| 1 | 31 (52) | 23 (36) | 54 (44) |
| 2 | 6 (10) | 9 (14) | 15 (12) |
| >2 | 23 (38) | 19 (30) | 42 (34) |
| **Development study details** |  |  |  |
| **Median (range) year of publication** | 2020 (2000 – 2023) | 1998 (1962 – 2015) | 2008 (1962 – 2023) |
| **Source of data** |  |  |  |
| Prospective | 8 (13) | 18 (28) | 26 (21) |
| Retrospective | 41 (68) | 10 (16) | 51 (41) |
| NS | 11 (18) | 36 (56) | 47 (38) |
| **Setting** |  |  |  |
| Hospital | 16 (27) | 11 (17) | 27 (22) |
| Long-term care (incl. end-of-life and rehab) | 8 (13) | 14 (22) | 22 (18) |
| Acute care (incl. surgical and ICU) | 33 (55) | 24 (38) | 57 (46) |
| Mixed settings | 1 (2) | 1 (2) | 2 (2) |
| Other | 2 (3) | 2 (3) | 4 (3) |
| NS | 0 (0) | 12 (19) | 12 (10) |
| **Study population age** |  |  |  |
| Adults | 36 (60) | 34 (53) | 70 (56) |
| Any | 4 (7) | 3 (5) | 7 (6) |
| NS | 20 (33) | 27 (42) | 47 (38) |
| **Baseline condition** |  |  |  |
| PIs at baseline | 1 (2) | 0 (0) | 1 (1) |
| No PIs at baseline | 11 (18) | 19 (30) | 30 (24) |
| NS | 48 (80) | 45 (70) | 93 (75) |
| **Development methods** |  |  |  |
| **Development method/algorithm<sup>B</sup>** |  |  |  |
| ML algorithms | 48 (80) | 0 (0) | 48 (39) |
| Logistic regression | 40 (67) | 15 (23)<sup>C</sup> | 55 (44) |
| Cox regression | 0 (0) | 5 (8) | 5 (4) |
| Fine-Gray model | 2 (3) | 0 (0) | 2 (2) |
| Clinical expertise | 0 (0) | 2 (3) | 2 (2) |
| NS | 0 (0) | 44 (69)<sup>D</sup> | 44 (35) |
| **Internal validation method<sup>B</sup>** |  |  |  |
| Cross-validation | 21 (35) | 3 (5)<sup>G</sup> | 24 (19) |
| Data splitting | 28 (47) | 0 (0) | 28 (23) |
| Not done / NS | 22 (37)<sup>F</sup> | 61 (95) | 83 (67) |
| **Median (range) no. of final predictors<sup>E</sup>** | 7 (3 – 23) | 8 (3 – 12) | 7 (3 – 23) |
| **Study cohort** |  |  |  |
| **Median (range) total sample size** | 2674 (27 – 1252313) | 285 (15 – 31150) | 686 (15 – 1252313) |
| **Median (range) number of events** | 207 (8 – 86410) | 51 (9 – 1350) | 98 (8 – 86410) |
| **Median (range) proportion of events**<br>**(% of sample size)** | 10.43% (0.42% – 80.00%) | 14.84% (1.18% – 46.67%) | 14.69% (0.42% – 80.00%) |

Note that tools were categorised as ML or non-ML tools based on the descriptions from authors of the included systematic reviews that the tools were identified in. <sup>A</sup> the 32 included systematic reviews; <sup>B</sup> tools use multiple methods, therefore total number not equal to N (100%); <sup>C</sup> one study also used discriminant analysis for model development; <sup>D</sup> many seemed to use clinical expertise, but development methods were not clearly reported; <sup>E</sup> counting of final predictors may vary between models: some authors may count individual factors, while others consider domains or subscales; <sup>F</sup> one review<sup>36</sup> implies 5 models did not implement internal validation; <sup>G</sup> 'resampling' (not described further) was used for the development of 2 models; ML – machine learning; NS – not stated; ICU – intensive care unit; PI – pressure injury.

Tools were categorised as having been developed with (60/124, 48%) or without (64/124, 52%) the use of ML methods (as defined by review authors). Prospectively collected data was used for model development for 21% of tools (26/124), retrospectively collected data for 41% (51/124), or was not reported (47/124). Information about the study populations was poorly reported, however study setting was reported for 112 prediction tools. Twenty-seven tools were reported to have been developed in hospital inpatients, and 22 were developed in long-term care settings, rehabilitation units or nursing homes or hospices. Where reported (n=100), sample sizes ranged from 15^101^ to 1,252,313.^102^ The approach to internal validation used for the prediction tools (e.g. cross-validation or split sample) was not reported at review-level for over two thirds of tools (83/124, 67%).

We could extract information about the predictors for only 66 of the 124 tools (Table 5 and Appendix 5). The most frequently included predictor was age (33/66, 50%), followed by pre-disposing diseases/conditions (32/66, 48%), medical treatment/care received (28/66, 42%) and mobility (27/66, 41%). Tools often (31/66, 47%) included multiple pre-existing conditions or comorbidities and multiple types of treatment or medication as predictors. Other common predictors include laboratory values, continence, nutrition, body-related values (e.g. weight, height, body temperature), mental status, activity, gender and skin assessment (27% to 35% of tools). Ten tools incorporated scores from other established risk prediction scales as a predictor, with eight including Braden^10^ ^11^ scores, one including the Norton^12^ score and one including the Waterlow^13^ score.

**Table 5.** Predictor categories and frequency (%) of inclusion in N=66 tools.

| Predictor category | No. of tools predictor appears in |
| --- | --- |
| Age | 33 (50) |
| Pre-disposing conditions | 32 (48) |
| Receiving medical treatment/care | 28 (42) |
| Mobility | 27 (41) |
| Laboratory values | 23 (35) |
| Continence | 22 (33) |
| Nutrition | 22 (33) |
| Body | 21 (32) |
| Mental Status | 21 (32) |
| Activity | 21 (32) |
| Gender | 21 (32) |
| Skin | 18 (27) |
| General Health | 14 (21) |
| Braden<sup>10 11</sup> score | 8 (12) |
| Length of stay | 8 (12) |
| Pressure injury | 7 (11) |
| Surgery duration | 6 (9) |
| Ability to ambulate | 6 (9) |
| Medical unit, ward, visit | 5 (8) |
| Ethnicity or place of birth | 5 (8) |
| Friction, shear, pressure | 3 (5) |
| Body position | 3 (5) |
| Pain | 3 (5) |
| Hygiene | 2 (3) |
| Isolation | 2 (3) |
| Smoking | 2 (3) |
| Norton<sup>12</sup> or Waterlow<sup>13</sup> score | 2 (3) |
| 'Special' (not explained) | 2 (3) |

Figures are given as count (% out of 66 tools with information on predictors). Note that multiple predictors may fall within the same predictor category. For instance, the category 'skin' may encompass both 'skin moisture' and 'skin integrity', with the frequency count reflecting the entire predictor category rather than individual predictors.

Only one review^52^ reported the presentation format of included tools, coded as ‘score system’ (n=11), ‘formula equation’ (n=3), ‘nomogram scale’ (n=2), or ‘not reported’ (n=6).

## DISCUSSION

This umbrella review summarises data from 32 eligible systematic reviews of PI risk prediction tools. Quality assessment using an adaptation of AMSTAR-2 revealed that most reviews were conducted to a relatively poor standard. Critical flaws were identified, including inadequate or absent reporting of protocols (23/32, 72%), inappropriate statistical synthesis methods (13/17, 76%) and lack of consideration for risk of bias judgements when discussing review results (17/32, 53%). Despite the large number of risk prediction models identified, only seven reviews reported information about model development and validation, predominantly for ML-based prediction models. The remaining reviews reported the accuracy (sensitivity and specificity), or effectiveness of identified models. The studies included in the ‘accuracy’ reviews that we identified, typically reported a binary classification of participants as high or low risk of PI based on the risk prediction tool scores, rather than constituting external validations of models. For many (44/64, 69%) prediction tools that were developed without the use of ML, we were not able to determine whether reliable and robust statistical methods were used or whether models were essentially risk assessment tools developed based on expert knowledge. For nearly half (58/124, 47%) of the identified tools, predictors included in the final models were not reported. Details of study populations and settings were also lacking. It was not always clear from the reviews whether the poor reporting occurred at review level or in the original primary study publications.

Model development algorithms included logistic regression, decision trees and random forests, with a vast number of ML-based models having been developed in the last five years. Although logistic regression is considered a statistical approach^107^, it does share some characteristics with ML methods.^108^ Modern ML frameworks and libraries have streamlined the automation of logistic regression, including feature selection, hyperparameter optimisation, and cross-validation, solidifying its role within the ML ecosystem; however, logistic regression may still appear in non-ML contexts, as some developers continue to apply it using more traditional methods. Most (6/7, 86%) of our set of reviews reported the use of logistic regression as part of an ML-based approach, however this reflects the classifications used by included systematic reviews as opposed to our own assessment of the methods used in the primary studies, and may therefore be an overestimation of the use of ML models.

In contrast to logistic regression approaches, decision trees and random forests may not produce a quantitative risk probability. Instead, they commonly categorise patients into binary ‘at risk’ or ‘not at risk’ groups. Although the risk probabilities generated in logistic regression prediction models can be useful for clinical decision making, it was not possible to derive any information about thresholds used to define ‘at risk’ or ‘not at risk’, and for most reviews, it was unclear what the final model comprised of. This lack of transparency poses potential hurdles in applying these models effectively in clinical settings.

A recent systematic review of risk of bias in ML-developed prediction models found that most models are of poor methodological quality and are at high risk of bias.^23^ In our set of reviews, of the four reviews that conducted a risk of bias assessment using the PROBAST tool, all models but one^103^ were found to be at high or unclear risk of bias.^37^ ^52–54^ This raises significant concerns about the accuracy of clinical risk predictions. This issue is particularly critical in light of emerging evidence^104^ on skin tone classification versus ethnicity/race-based methods in predicting pressure ulcer risk. These results underscore the need for developing bias-free predictive models to ensure accurate and equitable healthcare outcomes, especially in diverse patient populations.

Where the method of internal validation was reported, split-sample and cross-validation were the most commonly used techniques, however, detail was limited, and it was not possible to determine whether appropriate methods had been used. Although split-sample approaches have been favoured for model validation, more recent empirical work suggests that bootstrap-based optimism correction^105^ or cross-validation^106^ are preferred approaches. None of the included reviews reported the use of optimism correction approaches.

Only two reviews included external validations of previously developed models^52^ ^54^, however limited details of model performance were presented. External validation is necessary to ensure a model is both reproducible and generalisable^109^ ^110^, bringing the usefulness of the models included in these reviews into question. The PROGRESS framework suggests that multiple external validation studies should be conducted using independent datasets from different locations.^15^ In the two reviews that included model validation studies^52^ ^54^, it is unclear whether these studies were conducted in different locations. Where reported, they were all conducted in the same setting as the corresponding development study. PROGRESS also suggests that external validations are carried out in a variety of relevant settings. Shi and colleagues^52^ described four of eight validations as using ‘temporal’ data, which suggests that the validation population is largely the same as the development population but with use of data from different timeframes. This approach has been described as lying somewhere ‘between’ internal and external validation, further emphasising the need for well-designed external validation studies.^109^

Importantly, model recalibration was not reported for any external validations. Evidence suggests greater focus should be placed on large, well-designed external validation studies to validate and improve promising models (using recalibration and updating^111^), rather than developing a multitude of new ones.^15^ ^18^ Model validation and recalibration should be a continuous process, and this is something that future research should address. Following external validation, effectiveness studies should be conducted to assess the impact of model use on decision making, patient outcomes and costs.^15^

The effective use of prediction tools is also influenced by the way in which the model’s output is presented to the end-user. Only one review^52^ reported the presentation format of included tools, such as formula equations and nomograms. In conjunction with this, identifying and mitigating modifiable risk factors can help prevent PIs. Additional effort is needed in the development of risk prediction tools to extract predictors that are risk modifiers and provide end-users with this information, to make the predictions more interpretable and actionable.

Risk stratification in itself is not clinically useful unless it leads to an effective change in patient management. For instance, in high-risk groups, additional types of preventive interventions can be triggered, or default preventive measures can be applied more intensively (e.g., more frequent repositioning) based on the results of the risk assessment. While sensitivity and specificity are valid performance metrics, their optimisation must consider the cost of misclassification. Net benefit calculations, which can be visualised through decision curves,^112^ provide a more reliable means of evaluating the clinical utility of risk assessment for PIs across a range of thresholds at which clinical action is indicated. These calculations can assist in providing a balanced use of resources while maximising positive health outcomes, such as lowering incidence of PI.

It is also important to assess whether the tool can improve outcomes with existing preventive interventions and whether it integrates well into clinical workflows (i.e., clinical effectiveness). A well-developed tool with good calibration and discrimination properties may be of limited value if these practical concerns are not addressed. Therefore, model developers should check the expected value of prognosis and how the tool can guide prevention when employed in practice, before planning model development. If it’s determined that there is no value in predicting certain outcomes – that brings into question whether the model should even be developed.^113^

Despite the advances in methods for developing risk prediction models, scales developed using clinical expertise such as the Braden Scale^10^ ^11^, Norton Scale^12^, Waterlow Score^13^ and Cubbin-Jackson Scales^97^ ^98^ are extensively discussed in numerous clinical practice guidelines for patient risk assessment, and are commonly used in clinical practice.^6^ ^114^ Although guidelines recognise their low accuracy, they are still acknowledged, while other risk prediction models are not even considered. This may be due to the availability of at least some clinical trials evaluating the clinical utility of scales.^39^ Some scales, such as the Braden scale^10^ ^11^, are so widely used that they have become an integral component of risk assessment for PI in clinical practice, and have even been incorporated into EHRs. Their widespread use may impede the progress towards development, validation and evaluation of more accurate and innovative risk prediction models. Striking a balance between tradition and embracing advancements is crucial for effective implementation in healthcare settings and improving patient outcomes.

### Strengths and limitations

Our umbrella review is the first to systematically identify and evaluate systematic reviews of risk prediction models for PI. The review was conducted to a high standard, following Cochrane guidance^40^, and with a highly sensitive search strategy designed by an experienced information specialist. Although we excluded non-English publications due to time and resource constraints, where possible these publications were used to identify additional eligible risk prediction models. To some extent our review is limited by the use of AMSTAR-2 for quality assessment of included reviews. AMSTAR-2 was not designed for assessment of diagnostic or prognostic studies and, although we made some adaptations, many of the existing and amended criteria relate to the quality of reporting of the reviews as opposed to methodological quality. There is scope for further work to establish criteria for assessing systematic reviews of prediction models.

The main limitation, however, was the lack of detail about risk prediction models and risk prediction model performance that could be determined from the included systematic reviews. To be as comprehensive as possible in model identification, we were relatively generous in our definition of ‘systematic’, and this may have contributed to the often-poor level of detail provided by included reviews. It is likely, however, that reporting was poor in many of the primary studies contributing to these reviews. Excluding the ML-based models, more than half of available risk prediction scales or tools were published prior to the year 2000. The fact that the original versions of reporting guidelines for diagnostic accuracy studies^115^ and risk prediction models^116^ were not published until 2003 and 2015 respectively, is likely to have contributed to poor reporting. In contrast, the ML-based models were published between 2000 and 2023, with a median year of 2020. Reporting guidelines for development and validation of ML-based models are more recent^117^ ^118^, but aim to improve the reporting standards and understanding of evolving ML technologies in healthcare.

## CONCLUSIONS

There is a very large body of evidence reporting various risk prediction scales, tool and models for PI which has been summarised across multiple systematic reviews of varying methodological quality. Only five systematic reviews reported the development and validation of models to predict risk of PIs. It seems that for the most part, available models do not meet current standards for the development or reporting of risk prediction models. Furthermore, most available models, including ML-based models have not been validated beyond the original population in which they were developed. Identification of the optimal risk prediction model for PI from those currently available would require a high-quality systematic review of the primary literature, ideally limited to studies conducted to a high methodological standard. It is evident from our findings that there is still a lack of consensus on the optimal risk prediction model for PI, highlighting the need for more standardised and rigorous approaches in future research.

## Data Availability

All data produced in the present work are contained in the manuscript and supplementary file

## Declarations

### Ethics approval and consent to participate

Not applicable.

### Consent for publication

Not applicable.

### Availability of data and materials

All data produced in the present work are contained in the manuscript and supplementary file.

### Author Contributions

**Conceptualisation:** Bethany Hillier, Katie Scandrett, April Coombe, Tina Hernandez-Boussard, Ewout Steyerberg, Yemisi Takwoingi, Vladica Velickovic, Jacqueline Dinnes

**Data curation:** Bethany Hillier, Katie Scandrett, April Coombe, Jacqueline Dinnes

**Formal analysis:** Bethany Hillier, Katie Scandrett, Jacqueline Dinnes

**Funding acquisition:** Yemisi Takwoingi, Vladica Velickovic, Jacqueline Dinnes

**Investigation:** Bethany Hillier, Katie Scandrett, April Coombe, Yemisi Takwoingi, Jacqueline Dinnes

**Methodology:** Bethany Hillier, Katie Scandrett, April Coombe, Tina Hernandez-Boussard, Ewout Steyerberg, Yemisi Takwoingi, Vladica Velickovic, Jacqueline Dinnes

**Project administration:** Bethany Hillier, Yemisi Takwoingi, Jacqueline Dinnes

**Resources:** Bethany Hillier, Katie Scandrett

**Supervision:** Yemisi Takwoingi, Jacqueline Dinnes

**Writing – original draft:** Bethany Hillier, Katie Scandrett, April Coombe, Jacqueline Dinnes

**Writing – review & editing:** Bethany Hillier, Katie Scandrett, April Coombe, Tina Hernandez-Boussard, Ewout Steyerberg, Yemisi Takwoingi, Vladica Velickovic, Jacqueline Dinnes

## Acknowledgements

We would like to thank Mrs. Rosie Boodell (University of Birmingham, UK) for her help in acquiring the publications necessary to complete this piece of work.

