## Appendices for "Risk prediction tools for pressure injury occurrence: An umbrella review of systematic reviews reporting model development and validation methods"

### Appendix 1: PRISMA 2020 Checklist

| **Section and Topic** | **Item #** | **Checklist item** | **Location where item is reported** |
| --- | --- | --- | --- |
| **TITLE** | | |  |
| Title | 1 | Identify the report as a systematic review. | Title page and abstract |
| **ABSTRACT** | | |  |
| Abstract | 2 | See the PRISMA 2020 for Abstracts checklist. | Abstract |
| **INTRODUCTION** | | |  |
| Rationale | 3 | Describe the rationale for the review in the context of existing knowledge. | Introduction |
| Objectives | 4 | Provide an explicit statement of the objective(s) or question(s) the review addresses. | Introduction |
| **METHODS** | | |  |
| Eligibility criteria | 5 | Specify the inclusion and exclusion criteria for the review and how studies were grouped for the syntheses. | Methods: Literature search, Eligibility criteria for this umbrella review, Synthesis methods |
| Information sources | 6 | Specify all databases, registers, websites, organisations, reference lists and other sources searched or consulted to identify studies. Specify the date when each source was last searched or consulted. | Methods: Literature search  Appendix 2: Description of search strategies |
| Search strategy | 7 | Present the full search strategies for all databases, registers and websites, including any filters and limits used. | Appendix 2: Description of search strategies |
| Selection process | 8 | Specify the methods used to decide whether a study met the inclusion criteria of the review, including how many reviewers screened each record and each report retrieved, whether they worked independently, and if applicable, details of automation tools used in the process. | Methods: Literature search |
| Data collection process | 9 | Specify the methods used to collect data from reports, including how many reviewers collected data from each report, whether they worked independently, any processes for obtaining or confirming data from study investigators, and if applicable, details of automation tools used in the process. | Methods: Data extraction and quality assessment |
| Data items | 10a | List and define all outcomes for which data were sought. Specify whether all results that were compatible with each outcome domain in each study were sought (e.g. for all measures, time points, analyses), and if not, the methods used to decide which results to collect. | Methods: Data extraction and quality assessment, Synthesis methods  Appendix 3: Data extraction form |
|  | 10b | List and define all other variables for which data were sought (e.g. participant and intervention characteristics, funding sources). Describe any assumptions made about any missing or unclear information. | Appendix 3: Data extraction form  Methods: Synthesis |
| Study risk of bias assessment | 11 | Specify the methods used to assess risk of bias in the included studies, including details of the tool(s) used, how many reviewers assessed each study and whether they worked independently, and if applicable, details of automation tools used in the process. | Methods: Data extraction and quality assessment  Appendix 4: AMSTAR-2 Methodology Quality Appraisal. Adapted for application to reviews of prognostic model and accuracy studies. |
| Effect measures | 12 | Specify for each outcome the effect measure(s) (e.g. risk ratio, mean difference) used in the synthesis or presentation of results. | Methods: Synthesis methods |
| Synthesis methods | 13a | Describe the processes used to decide which studies were eligible for each synthesis (e.g. tabulating the study intervention characteristics and comparing against the planned groups for each synthesis (item #5)). | Methods: Synthesis methods |
|  | 13b | Describe any methods required to prepare the data for presentation or synthesis, such as handling of missing summary statistics, or data conversions. | -- |
|  | 13c | Describe any methods used to tabulate or visually display results of individual studies and syntheses. | Methods: Synthesis methods |
|  | 13d | Describe any methods used to synthesize results and provide a rationale for the choice(s). If meta-analysis was performed, describe the model(s), method(s) to identify the presence and extent of statistical heterogeneity, and software package(s) used. | Methods: Synthesis methods |
|  | 13e | Describe any methods used to explore possible causes of heterogeneity among study results (e.g. subgroup analysis, meta-regression). | -- |
|  | 13f | Describe any sensitivity analyses conducted to assess robustness of the synthesized results. | -- |
| Reporting bias assessment | 14 | Describe any methods used to assess risk of bias due to missing results in a synthesis (arising from reporting biases). | -- |
| Certainty assessment | 15 | Describe any methods used to assess certainty (or confidence) in the body of evidence for an outcome. | -- |
| **RESULTS** | | |  |
| Study selection | 16a | Describe the results of the search and selection process, from the number of records identified in the search to the number of studies included in the review, ideally using a flow diagram. | Results: Characteristics of included reviews  **Figure 1:** PRISMA flowchart |
|  | 16b | Cite studies that might appear to meet the inclusion criteria, but which were excluded, and explain why they were excluded. | **Appendix 5: Table S1.** Full-text articles excluded, with reasons |
| Study characteristics | 17 | Cite each included study and present its characteristics. | **Table 1.** Summary of included systematic review characteristics  **Appendix 5: Table S2.** Systematic review characteristics |
| Risk of bias in studies | 18 | Present assessments of risk of bias for each included study. | **Figure 2.** Summary of AMSTAR-2 assessment results  **Appendix 5: Table S3.** AMSTAR-2 assessment results per review |
| Results of individual studies | 19 | For all outcomes, present, for each study: (a) summary statistics for each group (where appropriate) and (b) an effect estimate and its precision (e.g. confidence/credible interval), ideally using structured tables or plots. | **Table 2.** Results of reviews reporting model development and validation  **Table 3.** Summary of tool characteristics, from review-level data |
| Results of syntheses | 20a | For each synthesis, briefly summarise the characteristics and risk of bias among contributing studies. | Results: Characteristics of included reviews, Methodological quality of included reviews  **Table 1.** Summary of included systematic review characteristics  **Figure 2.** Summary of AMSTAR-2 assessment results |
|  | 20b | Present results of all ~~statistical~~ syntheses conducted. If meta-analysis was done, present for each the summary estimate and its precision (e.g. confidence/credible interval) and measures of statistical heterogeneity. If comparing groups, describe the direction of the effect. | Results: Findings, Included tools and predictors  **Table 4.** Predictor categories and frequency (%) of inclusion in N=53 models.  **Appendix 5: Tables S4-S6** |
|  | 20c | Present results of all investigations of possible causes of heterogeneity among study results. | -- |
|  | 20d | Present results of all sensitivity analyses conducted to assess the robustness of the synthesized results. | -- |
| Reporting biases | 21 | Present assessments of risk of bias due to missing results (arising from reporting biases) for each synthesis assessed. | -- |
| Certainty of evidence | 22 | Present assessments of certainty (or confidence) in the body of evidence for each outcome assessed. | -- |
| **DISCUSSION** | | |  |
| Discussion | 23a | Provide a general interpretation of the results in the context of other evidence. | Discussion  Discussion: Other existing evidence |
|  | 23b | Discuss any limitations of the evidence included in the review. | Discussion: Strengths and limitations |
|  | 23c | Discuss any limitations of the review processes used. | Discussion: Strengths and limitations |
|  | 23d | Discuss implications of the results for practice, policy, and future research. | Discussion, Conclusions |
| **OTHER INFORMATION** | | |  |
| Registration and protocol | 24a | Provide registration information for the review, including register name and registration number, or state that the review was not registered. | Methods: Protocol registration and reporting of findings  OSF <https://osf.io/>tepyk |
|  | 24b | Indicate where the review protocol can be accessed, or state that a protocol was not prepared. | Methods: Protocol registration and reporting of findings  OSF <https://osf.io/>tepyk |
|  | 24c | Describe and explain any amendments to information provided at registration or in the protocol. | -- |
| Support | 25 | Describe sources of financial or non-financial support for the review, and the role of the funders or sponsors in the review. | Funding |
| Competing interests | 26 | Declare any competing interests of review authors. | Conflicting Interests |
| Availability of data, code and other materials | 27 | Report which of the following are publicly available and where they can be found: template data collection forms; data extracted from included studies; data used for all analyses; analytic code; any other materials used in the review. | **Appendix 3:** Data extraction form  **Appendix 5: Table S4-S6** |

*From:* Page MJ, McKenzie JE, Bossuyt PM, Boutron I, Hoffmann TC, Mulrow CD, et al. The PRISMA 2020 statement: an updated guideline for reporting systematic reviews. BMJ 2021;372:n71. doi: 10.1136/bmj.n71

For more information, visit: <http://www.prisma-statement.org/>

### Appendix 2: Description of search strategies

ORIGINAL SEARCH: JAN 2023

**Summary table of searches**

| **Source** | **Results before deduplication** | **Results after deduplication** |
| --- | --- | --- |
| MEDLINE | 1643 | 574 |
| EMBASE | 2060 | 1920 |
| CINAHL | 3720 | 3007 |
| EPISTEMONIKOS | 1194 | 574 |
| GOOGLE SCHOLAR | 357 | 226 |
| **TOTAL** | **8974** | **6301** |

SEARCH UPDATE: JUNE 2024

**Searches run from 01/01/23 - 06/24**

**Summary table of searches**

| **Source** | **Results before deduplication** | **Results after deduplication** |
| --- | --- | --- |
| MEDLINE | 229 | 186 |
| EMBASE | 330 | 175 |
| CINAHL | 480 | 383 |
| EPISTEMONIKOS | 232 | 118 |
| GOOGLE SCHOLAR | 80 | 36 |
| **TOTAL** | **1351** | **898** |

**Search approach and sources**

Search concepts:

i. pressure injury terms
ii. systematic review terms
iii. prediction model terms

Pressure injury (PI) terms were used from previous PI topic reviews and were developed in consultation with the wider review team and customer.

Established systematic review methodological filters were used in OVID Embase and OVID MEDLINE combining the appropriate McMasters best balance reviews filters^[1]^ combining the appropriate McMasters best balance systematic reviews filters^[2]^ with the appropriate CADTH systematic review filter^[3]^.

A number of existing methodological filters are available for prediction/prognostic model terms. The effect of using different combinations of these filters have been tested in order to ensure retrieval of relevant literature at a manageable volume. This testing has informed the choice of prognostic search filters used (Geersing)^[4]^ Haynes Best Balance^[5]^ and Ingui Best Balance^[6]^.

Searches were run in OVID MEDLINE, OVID Embase and EBSCO CINAHL Plus using the search concepts, systematic review and prediction/prognostic filters listed above or adaptations of these filters. No publication date or language restrictions were applied.

Epistemonikos was also searched using PI terms and key prognostic terms limited by publication type systematic review or broad synthesis. The Epistemonikos interface does not support the same search functionality available in OVID or EBSCO (for example adjacency operators are not supported). The Information Specialist ran several separate shorter searches to accommodate for the limitations of the interface. No publication date or language restrictions were applied.

In addition, Google Scholar was searched to pick up any potentially relevant papers not indexed in the other databases. The Google Scholar interface has limited search functionality. The Information Specialist ran several separate shorter searches to accommodate for the limitations of the interface. Searches were limited to review publication types published in the last eleven years only for pragmatic reasons as Google Scholar has poor export functionality.

“Connected papers” was also considered for inclusion, however it is a one ‘seed tool’, i.e., searching for one paper generates one map of connected papers. The platform is also only freely accessible for searching five ‘seed’ papers a month, which appears to be more of a limitation than would be beneficial for this set of reviews.

**MEDLINE ALL (OVID)**

**Date run: 31/01/23**

Database: Ovid MEDLINE(R) ALL <1946 to January 30, 2023>

Search Strategy:

--------------------------------------------------------------------------------

1	(decubit* or bedsore* or bed-sore* or pressure-ulcer* or pressure-wound*).tw. (17989)

2	((pressure* or bed or bedbound or bed-bound or bedridden or bed-ridden or deep tissue* or deep-tissue) adj3 (wound* or ulcer* or sore* or injur* or lesion*)).tw. (22136)

3	exp pressure ulcer/ or pressure/ae (15198)

4	1 or 2 or 3 (33198)

5	((supine or immobil*) adj3 (heal or healing or heals or healed or dress*)).tw. (220)

6	((supine or immobil*) adj3 (wound* or ulcer* or sore* or injur* or lesion*)).tw. (876)

7	((pressure or bedbound or bedridden or bed-bound or bed-ridden or deep tissue or deep-tissue) adj3 (heal or healing or heals or healed or dress*)).tw. (1863)

8	5 or 6 or 7 (2923)

9	4 or 8 (34931)

10	(systematic review or meta-analysis).pt. (299072)

11	review.pt. (3115171)

12	search:.tw. (617577)

13	meta-analys:.mp. (291117)

14	meta-analysis/ or systematic review/ or systematic reviews as topic/ or meta-analysis as topic/ or "meta analysis (topic)"/ or "systematic review (topic)"/ or exp technology assessment, biomedical/ or network meta-analysis/ (336517)

15	((systematic* adj3 (review* or overview*)) or (methodologic* adj3 (review* or overview*))).ti,ab,kf. (303236)

16	((quantitative adj3 (review* or overview* or synthes*)) or (research adj3 (integrati* or overview*))).ti,ab,kf. (15021)

17	((integrative adj3 (review* or overview*)) or (collaborative adj3 (review* or overview*)) or (pool* adj3 analy*)).ti,ab,kf. (37395)

18	(data synthes* or data extraction* or data abstraction*).ti,ab,kf. (38583)

19	(handsearch* or hand search*).ti,ab,kf. (10921)

20	(mantel haenszel or peto or der simonian or dersimonian or fixed effect* or latin square*).ti,ab,kf. (34465)

21	(met analy* or metanaly* or technology assessment* or HTA or HTAs or technology overview* or technology appraisal*).ti,ab,kf. (11813)

22	(meta regression* or metaregression*).ti,ab,kf. (13870)

23	(meta-analy* or metaanaly* or systematic review* or biomedical technology assessment* or bio-medical technology assessment*).mp,hw. (446528)

24	(medline or cochrane or pubmed or medlars or embase or cinahl).ti,ab,hw. (325867)

25	(cochrane or (health adj2 technology assessment) or evidence report).jw. (21207)

26	(comparative adj3 (efficacy or effectiveness)).ti,ab,kf. (17070)

27	(outcomes research or relative effectiveness).ti,ab,kf. (11017)

28	((indirect or indirect treatment or mixed-treatment or bayesian) adj3 comparison*).ti,ab,kf. (4214)

29	(multi* adj3 treatment adj3 comparison*).ti,ab,kf. (287)

30	(mixed adj3 treatment adj3 (meta-analy* or metaanaly*)).ti,ab,kf. (177)

31	umbrella review*.ti,ab,kf. (1305)

32	(multi* adj2 paramet* adj2 evidence adj2 synthesis).ti,ab,kf. (13)

33	(multiparamet* adj2 evidence adj2 synthesis).ti,ab,kf. (18)

34	(multi-paramet* adj2 evidence adj2 synthesis).ti,ab,kf. (11)

35	or/10-34 (3717455)

36	predict:.tw. or validat:.mp. or develop.tw. (3128744)

37	(stratification or ROC curve).ti,ab. or exp ROC curve/ or discriminat$.ti,ab. or c-statistic.ti,ab. or "Area under the curve".ti,ab. or AUC.ti,ab. or Calibration.ti,ab. or indices.ti,ab. or algorithm.ti,ab. or multivaria$.mp. (1567005)

38	Validat*.mp. or Predict$.ti. or Rule*.mp. or (Predict* and (Outcome* or Risk* or Model$)).mp. or ((History or Variable$ or Criteria or Scor$ or Characteristic$ or Finding$ or Factor$) and (Predict$ or Model$ or Decision$ or Identif$ or Prognos$)).mp. or (Decision$.mp. and ((Model$ or Clinical$).mp. or Logistic Models/)) or (Prognostic and (History or Variable$ or Criteria or Scor$ or Characteristic$ or Finding$ or Factor$ or Model$)).mp. [mp=title, book title, abstract, original title, name of substance word, subject heading word, floating sub-heading word, keyword heading word, organism supplementary concept word, protocol supplementary concept word, rare disease supplementary concept word, unique identifier, synonyms] (5961362)

39	36 or 37 (6673746)

40	39 or 38 (7368991)

41	9 and 35 and 40 (1643)

**MEDLINE ALL (OVID)**

**Date run: 20/06/24**

Database: Ovid MEDLINE(R) ALL <1946 to June 20, 2024>

Search Strategy:

--------------------------------------------------------------------------------

1	(decubit* or bedsore* or bed-sore* or pressure-ulcer* or pressure-wound*).tw.

2	((pressure* or bed or bedbound or bed-bound or bedridden or bed-ridden or deep tissue* or deep-tissue) adj3 (wound* or ulcer* or sore* or injur* or lesion*)).tw.

3	exp pressure ulcer/ or pressure/ae

4	1 or 2 or 3

5	((supine or immobil*) adj3 (heal or healing or heals or healed or dress*)).tw.

6	((supine or immobil*) adj3 (wound* or ulcer* or sore* or injur* or lesion*)).tw.

7	((pressure or bedbound or bedridden or bed-bound or bed-ridden or deep tissue or deep-tissue) adj3 (heal or healing or heals or healed or dress*)).tw.

8	5 or 6 or 7

9	4 or 8

10	(systematic review or meta-analysis).pt.

11	review.pt.

12	search:.tw.

13	meta-analys:.mp.

14	meta-analysis/ or systematic review/ or systematic reviews as topic/ or meta-analysis as topic/ or "meta analysis (topic)"/ or "systematic review (topic)"/ or exp technology assessment, biomedical/ or network meta-analysis/

15	((systematic* adj3 (review* or overview*)) or (methodologic* adj3 (review* or overview*))).ti,ab,kf.

16	((quantitative adj3 (review* or overview* or synthes*)) or (research adj3 (integrati* or overview*))).ti,ab,kf.

17	((integrative adj3 (review* or overview*)) or (collaborative adj3 (review* or overview*)) or (pool* adj3 analy*)).ti,ab,kf.

18	(data synthes* or data extraction* or data abstraction*).ti,ab,kf.

19	(handsearch* or hand search*).ti,ab,kf.

20	(mantel haenszel or peto or der simonian or dersimonian or fixed effect* or latin square*).ti,ab,kf.

21	(met analy* or metanaly* or technology assessment* or HTA or HTAs or technology overview* or technology appraisal*).ti,ab,kf.

22	(meta regression* or metaregression*).ti,ab,kf.

23	(meta-analy* or metaanaly* or systematic review* or biomedical technology assessment* or bio-medical technology assessment*).mp,hw.

24	(medline or cochrane or pubmed or medlars or embase or cinahl).ti,ab,hw.

25	(cochrane or (health adj2 technology assessment) or evidence report).jw.

26	(comparative adj3 (efficacy or effectiveness)).ti,ab,kf.

27	(outcomes research or relative effectiveness).ti,ab,kf.

28	((indirect or indirect treatment or mixed-treatment or bayesian) adj3 comparison*).ti,ab,kf.

29	(multi* adj3 treatment adj3 comparison*).ti,ab,kf.

30	(mixed adj3 treatment adj3 (meta-analy* or metaanaly*)).ti,ab,kf.

31	umbrella review*.ti,ab,kf.

32	(multi* adj2 paramet* adj2 evidence adj2 synthesis).ti,ab,kf.

33	(multiparamet* adj2 evidence adj2 synthesis).ti,ab,kf.

34	(multi-paramet* adj2 evidence adj2 synthesis).ti,ab,kf.

35	or/10-34

36	predict:.tw. or validat:.mp. or develop.tw.

37	(stratification or ROC curve).ti,ab. or exp ROC curve/ or discriminat$.ti,ab. or c-statistic.ti,ab. or "Area under the curve".ti,ab. or AUC.ti,ab. or Calibration.ti,ab. or indices.ti,ab. or algorithm.ti,ab. or multivaria$.mp.

38	Validat*.mp. or Predict$.ti. or Rule*.mp. or (Predict* and (Outcome* or Risk* or Model$)).mp. or ((History or Variable$ or Criteria or Scor$ or Characteristic$ or Finding$ or Factor$) and (Predict$ or Model$ or Decision$ or Identif$ or Prognos$)).mp. or (Decision$.mp. and ((Model$ or Clinical$).mp. or Logistic Models/)) or (Prognostic and (History or Variable$ or Criteria or Scor$ or Characteristic$ or Finding$ or Factor$ or Model$)).mp. [mp=title, book title, abstract, original title, name of substance word, subject heading word, floating sub-heading word, keyword heading word, organism supplementary concept word, protocol supplementary concept word, rare disease supplementary concept word, unique identifier, synonyms]

39	36 or 37

40	39 or 38

41	9 and 35 and 40

42	limit 41 to dt=20230101-20241231

43	limit 41 to ez=20230101-20241231

44	limit 42 to da=20230101-20241231

45	42 or 43 or 44

**Embase (OVID)**

**Date run: 31/01/23**

Database: Embase <1974 to 2023 January 30>

Search Strategy:

--------------------------------------------------------------------------------

1	(decubit* or bedsore* or bed-sore* or pressure-ulcer* or pressure-wound*).tw. (24186)

2	((pressure* or bed or bedbound or bed-bound or bedridden or bed-ridden or deep tissue* or deep-tissue) adj3 (wound* or ulcer* or sore* or injur* or lesion*)).tw. (28692)

3	exp decubitus/ (24330)

4	1 or 2 or 3 (45109)

5	((supine or immobil*) adj3 (heal or healing or heals or healed or dress*)).tw. (279)

6	((supine or immobil*) adj3 (wound* or ulcer* or sore* or injur* or lesion*)).tw. (1181)

7	((pressure or bedbound or bedridden or bed-bound or bed-ridden or deep tissue or deep-tissue) adj3 (heal or healing or heals or healed or dress*)).tw. (2463)

8	5 or 6 or 7 (3873)

9	4 or 8 (47397)

10	(systematic review or meta-analysis).pt. (0)

11	review.pt. (3006963)

12	search:.tw. (777617)

13	meta-analys:.mp. (420287)

14	meta-analysis/ or systematic review/ or systematic reviews as topic/ or meta-analysis as topic/ or "meta analysis (topic)"/ or "systematic review (topic)"/ or exp technology assessment, biomedical/ or network meta-analysis/ (600348)

15	((systematic* adj3 (review* or overview*)) or (methodologic* adj3 (review* or overview*))).ti,ab,kf. (374423)

16	((quantitative adj3 (review* or overview* or synthes*)) or (research adj3 (integrati* or overview*))).ti,ab,kf. (17492)

17	((integrative adj3 (review* or overview*)) or (collaborative adj3 (review* or overview*)) or (pool* adj3 analy*)).ti,ab,kf. (53239)

18	(data synthes* or data extraction* or data abstraction*).ti,ab,kf. (47755)

19	(handsearch* or hand search*).ti,ab,kf. (13352)

20	(mantel haenszel or peto or der simonian or dersimonian or fixed effect* or latin square*).ti,ab,kf. (45672)

21	(met analy* or metanaly* or technology assessment* or HTA or HTAs or technology overview* or technology appraisal*).ti,ab,kf. (19758)

22	(meta regression* or metaregression*).ti,ab,kf. (17239)

23	(meta-analy* or metaanaly* or systematic review* or biomedical technology assessment* or bio-medical technology assessment*).mp,hw. (709507)

24	(medline or cochrane or pubmed or medlars or embase or cinahl).ti,ab,hw. (427516)

25	(cochrane or (health adj2 technology assessment) or evidence report).jw. (30592)

26	(comparative adj3 (efficacy or effectiveness)).ti,ab,kf. (25197)

27	(outcomes research or relative effectiveness).ti,ab,kf. (15991)

28	((indirect or indirect treatment or mixed-treatment or bayesian) adj3 comparison*).ti,ab,kf. (7335)

29	(multi* adj3 treatment adj3 comparison*).ti,ab,kf. (423)

30	(mixed adj3 treatment adj3 (meta-analy* or metaanaly*)).ti,ab,kf. (256)

31	umbrella review*.ti,ab,kf. (1367)

32	(multi* adj2 paramet* adj2 evidence adj2 synthesis).ti,ab,kf. (28)

33	(multiparamet* adj2 evidence adj2 synthesis).ti,ab,kf. (21)

34	(multi-paramet* adj2 evidence adj2 synthesis).ti,ab,kf. (23)

35	or/10-34 (3937855)

36	validat:.mp. or index.tw. or model.tw. (5261274)

37	(stratification or ROC curve).ti,ab. or exp receiver operating characteristic/ or discriminat$.ti,ab. or c-statistic.ti,ab. or "Area under the curve".ti,ab. or AUC.ti,ab. or Calibration.ti,ab. or indices.ti,ab. or algorithm.ti,ab. or multivaria$.mp. (2151295)

38	Validat*.mp. or Predict$.ti. or Rule*.mp. or (Predict* and (Outcome* or Risk* or Model$)).mp. or ((History or Variable$ or Criteria or Scor$ or Characteristic$ or Finding$ or Factor$) and (Predict$ or Model$ or Decision$ or Identif$ or Prognos$)).mp. or (Decision$.mp. and ((Model$ or Clinical$).mp. or Statistical model/)) or (Prognostic and (History or Variable$ or Criteria or Scor$ or Characteristic$ or Finding$ or Factor$ or Model$)).mp. [mp=title, abstract, heading word, drug trade name, original title, device manufacturer, drug manufacturer, device trade name, keyword heading word, floating subheading word, candidate term word] (8141367)

39	36 or 37 (9046587)

40	39 or 38 (10998837)

47	9 and 35 and 40 (2060)

**Embase (OVID)**

**Date run: 20/06/24**

Database: Embase <1974 to 2024 June 20>

Search Strategy:

--------------------------------------------------------------------------------

1	(decubit* or bedsore* or bed-sore* or pressure-ulcer* or pressure-wound*).tw.

2	((pressure* or bed or bedbound or bed-bound or bedridden or bed-ridden or deep tissue* or deep-tissue) adj3 (wound* or ulcer* or sore* or injur* or lesion*)).tw.

3	exp decubitus/

4	1 or 2 or 3

5	((supine or immobil*) adj3 (heal or healing or heals or healed or dress*)).tw.

6	((supine or immobil*) adj3 (wound* or ulcer* or sore* or injur* or lesion*)).tw.

7	((pressure or bedbound or bedridden or bed-bound or bed-ridden or deep tissue or deep-tissue) adj3 (heal or healing or heals or healed or dress*)).tw.

8	5 or 6 or 7

9	4 or 8

10	(systematic review or meta-analysis).pt.

11	review.pt.

12	search:.tw.

13	meta-analys:.mp.

14	meta-analysis/ or systematic review/ or systematic reviews as topic/ or meta-analysis as topic/ or "meta analysis (topic)"/ or "systematic review (topic)"/ or exp technology assessment, biomedical/ or network meta-analysis/

15	((systematic* adj3 (review* or overview*)) or (methodologic* adj3 (review* or overview*))).ti,ab,kf.

16	((quantitative adj3 (review* or overview* or synthes*)) or (research adj3 (integrati* or overview*))).ti,ab,kf.

17	((integrative adj3 (review* or overview*)) or (collaborative adj3 (review* or overview*)) or (pool* adj3 analy*)).ti,ab,kf.

18	(data synthes* or data extraction* or data abstraction*).ti,ab,kf.

19	(handsearch* or hand search*).ti,ab,kf.

20	(mantel haenszel or peto or der simonian or dersimonian or fixed effect* or latin square*).ti,ab,kf.

21	(met analy* or metanaly* or technology assessment* or HTA or HTAs or technology overview* or technology appraisal*).ti,ab,kf.

22	(meta regression* or metaregression*).ti,ab,kf.

23	(meta-analy* or metaanaly* or systematic review* or biomedical technology assessment* or bio-medical technology assessment*).mp,hw.

24	(medline or cochrane or pubmed or medlars or embase or cinahl).ti,ab,hw.

25	(cochrane or (health adj2 technology assessment) or evidence report).jw.

26	(comparative adj3 (efficacy or effectiveness)).ti,ab,kf.

27	(outcomes research or relative effectiveness).ti,ab,kf.

28	((indirect or indirect treatment or mixed-treatment or bayesian) adj3 comparison*).ti,ab,kf.

29	(multi* adj3 treatment adj3 comparison*).ti,ab,kf.

30	(mixed adj3 treatment adj3 (meta-analy* or metaanaly*)).ti,ab,kf.

31	umbrella review*.ti,ab,kf.

32	(multi* adj2 paramet* adj2 evidence adj2 synthesis).ti,ab,kf.

33	(multiparamet* adj2 evidence adj2 synthesis).ti,ab,kf.

34	(multi-paramet* adj2 evidence adj2 synthesis).ti,ab,kf.

35	or/10-34

36	validat:.mp. or index.tw. or model.tw.

37	(stratification or ROC curve).ti,ab. or exp receiver operating characteristic/ or discriminat$.ti,ab. or c-statistic.ti,ab. or "Area under the curve".ti,ab. or AUC.ti,ab. or Calibration.ti,ab. or indices.ti,ab. or algorithm.ti,ab. or multivaria$.mp.

38	Validat*.mp. or Predict$.ti. or Rule*.mp. or (Predict* and (Outcome* or Risk* or Model$)).mp. or ((History or Variable$ or Criteria or Scor$ or Characteristic$ or Finding$ or Factor$) and (Predict$ or Model$ or Decision$ or Identif$ or Prognos$)).mp. or (Decision$.mp. and ((Model$ or Clinical$).mp. or Statistical model/)) or (Prognostic and (History or Variable$ or Criteria or Scor$ or Characteristic$ or Finding$ or Factor$ or Model$)).mp. [mp=title, abstract, heading word, drug trade name, original title, device manufacturer, drug manufacturer, device trade name, keyword heading word, floating subheading word, candidate term word] (8141367)

39	36 or 37

40	39 or 38

47	9 and 35 and 40

48	limit 47 to dc=20230101-20243112

**CINAHL PLUS (EBSCOhost)**
*SEARCH DATE 02/02/23*

S1	( ((TI decubit* OR AB decubit*) OR (TI bedsore* OR AB bedsore*) OR (TI bed-sore* OR AB bed-sore*) OR (TI pressure-ulcer* OR AB pressure-ulcer*) OR (TI pressure-wound* OR AB pressure-wound*)) ) OR ( (((TI pressure* OR AB pressure*) OR (TI bed OR AB bed) OR (TI bedbound OR AB bedbound) OR (TI bed-bound OR AB bed-bound) OR (TI bedridden OR AB bedridden) OR (TI bed-ridden OR AB bed-ridden) OR (TI "deep tissue*" OR AB "deep tissue*") OR (TI deep-tissue OR AB deep-tissue)) N3 ((TI wound* OR AB wound*) OR (TI ulcer* OR AB ulcer*) OR (TI sore* OR AB sore*) OR (TI injur* OR AB injur*) OR (TI lesion* OR AB lesion*))) ) OR ( (MH "pressure ulcer"+) OR (MH pressure) )	Expanders - Apply equivalent subjects (28 401)

S2	( ((TI heal OR AB heal) OR (TI healing OR AB healing) OR (TI heals OR AB heals) OR (TI healed OR AB healed) OR (TI dress* OR AB dress*))) ) OR ( (((TI supine OR AB supine) OR (TI immobil* OR AB immobil*)) N3 ((TI wound* OR AB wound*) OR (TI ulcer* OR AB ulcer*) OR (TI sore* OR AB sore*) OR (TI injur* OR AB injur*) OR (TI lesion* OR AB lesion*))) ) OR ( (((TI pressure OR AB pressure) OR (TI bedbound OR AB bedbound) OR (TI bedridden OR AB bedridden) OR (TI bed-bound OR AB bed-bound) OR (TI bed-ridden OR AB bed-ridden) OR (TI "deep tissue" OR AB "deep tissue") OR (TI deep-tissue OR AB deep-tissue)) N3 ((TI heal OR AB heal) OR (TI healing OR AB healing) OR (TI heals OR AB heals) OR (TI healed OR AB healed) OR (TI dress* OR AB dress*))) )	Expanders – Apply equivalent subjects (72 286)

S3	S1 OR S2	Expanders - Apply equivalent subjects (96245)

S4	( (PT "systematic review" OR PT meta-analysis) ) OR ( (MH meta-analysis) OR (MH "systematic review") OR (MH "systematic reviews as topic") OR (MH "meta-analysis as topic") OR (MH "meta analysis (topic)") OR (MH "systematic review (topic)") OR (MH "technology assessment, biomedical"+) OR (MH "network meta-analysis") ) OR ( (((TI systematic* OR AB systematic* OR SU systematic*) N3 ((TI review* OR AB review* OR SU review*) OR (TI overview* OR AB overview* OR SU overview*))) OR ((TI methodologic* OR AB methodologic* OR SU methodologic*) N3 ((TI review* OR AB review* OR SU review*) OR (TI overview* OR AB overview* OR SU overview*)))) ) OR ( (((TI quantitative OR AB quantitative OR SU quantitative) N3 ((TI review* OR AB review* OR SU review*) OR (TI overview* OR AB overview* OR SU overview*) OR (TI synthes* OR AB synthes* OR SU synthes*))) OR ((TI research OR AB research OR SU research) N3 ((TI integrati* OR AB integrati* OR SU integrati*) OR (TI overview* OR AB overview* OR SU overview*)))) ) OR ( (((TI integrative OR AB integrative OR SU integrative) N3 ((TI review* OR AB review* OR SU review*) OR (TI overview* OR AB overview* OR SU overview*))) OR ((TI collaborative OR AB collaborative OR SU collaborative) N3 ((TI review* OR AB review* OR SU review*) OR (TI overview* OR AB overview* OR SU overview*))) OR ((TI pool* OR AB pool* OR SU pool*) N3 (TI analy* OR AB analy* OR SU analy*))) ) OR ( ((TI "data synthes*" OR AB "data synthes*" OR SU "data synthes*") OR (TI "data extraction*" OR AB "data extraction*" OR SU "data extraction*") OR (TI "data abstraction*" OR AB "data abstraction*" OR SU "data abstraction*")) ) OR ( ((TI handsearch* OR AB handsearch* OR SU handsearch*) OR (TI "hand search*" OR AB "hand search*" OR SU "hand search*")) ) OR ( ((TI "mantel haenszel" OR AB "mantel haenszel" OR SU "mantel haenszel") OR (TI peto OR AB peto OR SU peto) OR (TI "der simonian" OR AB "der simonian" OR SU "der simonian") OR (TI dersimonian OR AB dersimonian OR SU dersimonian) OR (TI "fixed effect*" OR AB "fixed effect*" OR SU "fixed effect*") OR (TI "latin square*" OR AB "latin square*" OR SU "latin square*")) ) OR ( ((TI "met analy*" OR AB "met analy*" OR SU "met analy*") OR (TI metanaly* OR AB metanaly* OR SU metanaly*) OR (TI "technology assessment*" OR AB "technology assessment*" OR SU "technology assessment*") OR (TI HTA OR AB HTA OR SU HTA) OR (TI HTAs OR AB HTAs OR SU HTAs) OR (TI "technology overview*" OR AB "technology overview*" OR SU "technology overview*") OR (TI "technology appraisal*" OR AB "technology appraisal*" OR SU "technology appraisal*")) ) OR ( ((TI "meta regression*" OR AB "meta regression*" OR SU "meta regression*") OR (TI metaregression* OR AB metaregression* OR SU metaregression*)) ) OR ( (meta-analy* OR metaanaly* OR "systematic review*" OR "biomedical technology assessment*" OR "bio-medical technology assessment*") ,hw. ) OR ( ((TI medline OR AB medline) OR (TI cochrane OR AB cochrane) OR (TI pubmed OR AB pubmed) OR (TI medlars OR AB medlars) OR (TI embase OR AB embase) OR (TI cinahl OR AB cinahl)) ,hw. )	Expanders - Apply equivalent subjects (237782)

S5	( (cochrane OR ( health N2 "technology assessment") OR "evidence report") .jw. ) OR ( ((TI comparative OR AB comparative OR SU comparative) N3 ((TI efficacy OR AB efficacy OR SU efficacy) OR (TI effectiveness OR AB effectiveness OR SU effectiveness))) ) OR ( ((TI "outcomes research" OR AB "outcomes research" OR SU "outcomes research") OR (TI "relative effectiveness" OR AB "relative effectiveness" OR SU "relative effectiveness")) ) OR ( (((TI indirect OR AB indirect OR SU indirect) OR (TI "indirect treatment" OR AB "indirect treatment" OR SU "indirect treatment") OR (TI mixed-treatment OR AB mixed-treatment OR SU mixed-treatment) OR (TI bayesian OR AB bayesian OR SU bayesian)) N3 (TI comparison* OR AB comparison* OR SU comparison*)) ) OR ( ((TI multi* OR AB multi* OR SU multi*) N3 (TI treatment OR AB treatment OR SU treatment) N3 (TI comparison* OR AB comparison* OR SU comparison*)) ) OR ( (TI "umbrella review*" OR AB "umbrella review*" OR SU "umbrella review*") ) OR ( ((TI multi* OR AB multi* OR SU multi*) N2 (TI paramet* OR AB paramet* OR SU paramet*) N2 (TI evidence OR AB evidence OR SU evidence) N2 (TI synthesis OR AB synthesis OR SU synthesis)) ) OR ( ((TI multiparamet* OR AB multiparamet* OR SU multiparamet*) N2 (TI evidence OR AB evidence OR SU evidence) N2 (TI synthesis OR AB synthesis OR SU synthesis)) ) OR ( ((TI multi-paramet* OR AB multi-paramet* OR SU multi-paramet*) N2 (TI evidence OR AB evidence OR SU evidence) N2 (TI synthesis OR AB synthesis OR SU synthesis))	Expanders - Apply equivalent subjects (19108)

S6	((TI search: OR AB search:))	Expanders - Apply equivalent subjects (123419)

S7	PT review	Expanders - Apply equivalent subjects (356003)

S8	S4 OR S5 OR S6 OR S7	Expanders - Apply equivalent subjects (645360)

S9	S3 AND S8	Expanders - Apply equivalent subjects (10379)

S100	(TX validat*) or (TI index or model) or (AB index or model) Expanders - Apply equivalent subjects (1286240)xpanders - Apply

S11	TI ( stratification or "ROC curve" or discriminat" or c-statistic" or "Area under the curve" or AUC or Calibration* or indices* or algorithm* or multivaria* ) OR AB ( stratification or "ROC curve" or discriminat" or c-statistic" or "Area under the curve" or AUC or Calibration* or indices* or algorithm* or multivaria* )	Expanders - Apply equivalent subjects (294871)

S12	(MH "ROC Curve")	Expanders - Apply equivalent subjects (33393)

S13	TI Validat* or Predict* or Rule* or (Predict* and (Outcome* or Risk* or Model*)) or ((History or Variable* or Criteria or Scor* or Characteristic* or Finding* or Factor*) and (Predict* or Model* or Decision* or Identif* or Prognos*)) or (Decision* and ((Model* or Clinical*) or (Prognostic and (History or Variable* or Criteria or Scor* or Characteristic* or Finding* or Factor* or Model*))	Expanders - Apply equivalent subjects (144228)

S14	AB Validat* or Predict* or Rule* or (Predict* and (Outcome* or Risk* or Model*)) or ((History or Variable* or Criteria or Scor* or Characteristic* or Finding* or Factor*) and (Predict* or Model* or Decision* or Identif* or Prognos*)) or (Decision* and ((Model* or Clinical*) or (Prognostic and (History or Variable* or Criteria or Scor* or Characteristic* or Finding* or Factor* or Model*))	Expanders - Apply equivalent subjects (1482692)

S15	(MH "Models, Statistical+")	Expanders - Apply equivalent subjects (40243)

S16	S10 OR S11 OR S12 OR S13 OR S14 OR S15	Expanders - Apply equivalent subjects (2119713)

S17	S9 AND S16 (3720)

**CINAHL PLUS (EBSCOhost)**
*SEARCH DATE 21/06/24*

S1	( ((TI decubit* OR AB decubit*) OR (TI bedsore* OR AB bedsore*) OR (TI bed-sore* OR AB bed-sore*) OR (TI pressure-ulcer* OR AB pressure-ulcer*) OR (TI pressure-wound* OR AB pressure-wound*)) ) OR ( (((TI pressure* OR AB pressure*) OR (TI bed OR AB bed) OR (TI bedbound OR AB bedbound) OR (TI bed-bound OR AB bed-bound) OR (TI bedridden OR AB bedridden) OR (TI bed-ridden OR AB bed-ridden) OR (TI "deep tissue*" OR AB "deep tissue*") OR (TI deep-tissue OR AB deep-tissue)) N3 ((TI wound* OR AB wound*) OR (TI ulcer* OR AB ulcer*) OR (TI sore* OR AB sore*) OR (TI injur* OR AB injur*) OR (TI lesion* OR AB lesion*))) ) OR ( (MH "pressure ulcer"+) OR (MH pressure) )	Expanders - Apply equivalent subjects

S2	( ((TI heal OR AB heal) OR (TI healing OR AB healing) OR (TI heals OR AB heals) OR (TI healed OR AB healed) OR (TI dress* OR AB dress*))) ) OR ( (((TI supine OR AB supine) OR (TI immobil* OR AB immobil*)) N3 ((TI wound* OR AB wound*) OR (TI ulcer* OR AB ulcer*) OR (TI sore* OR AB sore*) OR (TI injur* OR AB injur*) OR (TI lesion* OR AB lesion*))) ) OR ( (((TI pressure OR AB pressure) OR (TI bedbound OR AB bedbound) OR (TI bedridden OR AB bedridden) OR (TI bed-bound OR AB bed-bound) OR (TI bed-ridden OR AB bed-ridden) OR (TI "deep tissue" OR AB "deep tissue") OR (TI deep-tissue OR AB deep-tissue)) N3 ((TI heal OR AB heal) OR (TI healing OR AB healing) OR (TI heals OR AB heals) OR (TI healed OR AB healed) OR (TI dress* OR AB dress*))) )	Expanders – Apply equivalent subjects

S3	S1 OR S2	Expanders - Apply equivalent subjects

S4	( (PT "systematic review" OR PT meta-analysis) ) OR ( (MH meta-analysis) OR (MH "systematic review") OR (MH "systematic reviews as topic") OR (MH "meta-analysis as topic") OR (MH "meta analysis (topic)") OR (MH "systematic review (topic)") OR (MH "technology assessment, biomedical"+) OR (MH "network meta-analysis") ) OR ( (((TI systematic* OR AB systematic* OR SU systematic*) N3 ((TI review* OR AB review* OR SU review*) OR (TI overview* OR AB overview* OR SU overview*))) OR ((TI methodologic* OR AB methodologic* OR SU methodologic*) N3 ((TI review* OR AB review* OR SU review*) OR (TI overview* OR AB overview* OR SU overview*)))) ) OR ( (((TI quantitative OR AB quantitative OR SU quantitative) N3 ((TI review* OR AB review* OR SU review*) OR (TI overview* OR AB overview* OR SU overview*) OR (TI synthes* OR AB synthes* OR SU synthes*))) OR ((TI research OR AB research OR SU research) N3 ((TI integrati* OR AB integrati* OR SU integrati*) OR (TI overview* OR AB overview* OR SU overview*)))) ) OR ( (((TI integrative OR AB integrative OR SU integrative) N3 ((TI review* OR AB review* OR SU review*) OR (TI overview* OR AB overview* OR SU overview*))) OR ((TI collaborative OR AB collaborative OR SU collaborative) N3 ((TI review* OR AB review* OR SU review*) OR (TI overview* OR AB overview* OR SU overview*))) OR ((TI pool* OR AB pool* OR SU pool*) N3 (TI analy* OR AB analy* OR SU analy*))) ) OR ( ((TI "data synthes*" OR AB "data synthes*" OR SU "data synthes*") OR (TI "data extraction*" OR AB "data extraction*" OR SU "data extraction*") OR (TI "data abstraction*" OR AB "data abstraction*" OR SU "data abstraction*")) ) OR ( ((TI handsearch* OR AB handsearch* OR SU handsearch*) OR (TI "hand search*" OR AB "hand search*" OR SU "hand search*")) ) OR ( ((TI "mantel haenszel" OR AB "mantel haenszel" OR SU "mantel haenszel") OR (TI peto OR AB peto OR SU peto) OR (TI "der simonian" OR AB "der simonian" OR SU "der simonian") OR (TI dersimonian OR AB dersimonian OR SU dersimonian) OR (TI "fixed effect*" OR AB "fixed effect*" OR SU "fixed effect*") OR (TI "latin square*" OR AB "latin square*" OR SU "latin square*")) ) OR ( ((TI "met analy*" OR AB "met analy*" OR SU "met analy*") OR (TI metanaly* OR AB metanaly* OR SU metanaly*) OR (TI "technology assessment*" OR AB "technology assessment*" OR SU "technology assessment*") OR (TI HTA OR AB HTA OR SU HTA) OR (TI HTAs OR AB HTAs OR SU HTAs) OR (TI "technology overview*" OR AB "technology overview*" OR SU "technology overview*") OR (TI "technology appraisal*" OR AB "technology appraisal*" OR SU "technology appraisal*")) ) OR ( ((TI "meta regression*" OR AB "meta regression*" OR SU "meta regression*") OR (TI metaregression* OR AB metaregression* OR SU metaregression*)) ) OR ( (meta-analy* OR metaanaly* OR "systematic review*" OR "biomedical technology assessment*" OR "bio-medical technology assessment*") ,hw. ) OR ( ((TI medline OR AB medline) OR (TI cochrane OR AB cochrane) OR (TI pubmed OR AB pubmed) OR (TI medlars OR AB medlars) OR (TI embase OR AB embase) OR (TI cinahl OR AB cinahl)) ,hw. )

S5	( (cochrane OR ( health N2 "technology assessment") OR "evidence report") .jw. ) OR ( ((TI comparative OR AB comparative OR SU comparative) N3 ((TI efficacy OR AB efficacy OR SU efficacy) OR (TI effectiveness OR AB effectiveness OR SU effectiveness))) ) OR ( ((TI "outcomes research" OR AB "outcomes research" OR SU "outcomes research") OR (TI "relative effectiveness" OR AB "relative effectiveness" OR SU "relative effectiveness")) ) OR ( (((TI indirect OR AB indirect OR SU indirect) OR (TI "indirect treatment" OR AB "indirect treatment" OR SU "indirect treatment") OR (TI mixed-treatment OR AB mixed-treatment OR SU mixed-treatment) OR (TI bayesian OR AB bayesian OR SU bayesian)) N3 (TI comparison* OR AB comparison* OR SU comparison*)) ) OR ( ((TI multi* OR AB multi* OR SU multi*) N3 (TI treatment OR AB treatment OR SU treatment) N3 (TI comparison* OR AB comparison* OR SU comparison*)) ) OR ( (TI "umbrella review*" OR AB "umbrella review*" OR SU "umbrella review*") ) OR ( ((TI multi* OR AB multi* OR SU multi*) N2 (TI paramet* OR AB paramet* OR SU paramet*) N2 (TI evidence OR AB evidence OR SU evidence) N2 (TI synthesis OR AB synthesis OR SU synthesis)) ) OR ( ((TI multiparamet* OR AB multiparamet* OR SU multiparamet*) N2 (TI evidence OR AB evidence OR SU evidence) N2 (TI synthesis OR AB synthesis OR SU synthesis)) ) OR ( ((TI multi-paramet* OR AB multi-paramet* OR SU multi-paramet*) N2 (TI evidence OR AB evidence OR SU evidence) N2 (TI synthesis OR AB synthesis OR SU synthesis))

S6	((TI search: OR AB search:))

S7	PT review

S8	S4 OR S5 OR S6 OR S7

S9	S3 AND S8

S100	(TX validat*) or (TI index or model) or (AB index or model) s - Apply

S11	TI ( stratification or "ROC curve" or discriminat" or c-statistic" or "Area under the curve" or AUC or Calibration* or indices* or algorithm* or multivaria* ) OR AB ( stratification or "ROC curve" or discriminat" or c-statistic" or "Area under the curve" or AUC or Calibration* or indices* or algorithm* or multivaria* )

S12	(MH "ROC Curve")

S13	TI Validat* or Predict* or Rule* or (Predict* and (Outcome* or Risk* or Model*)) or ((History or Variable* or Criteria or Scor* or Characteristic* or Finding* or Factor*) and (Predict* or Model* or Decision* or Identif* or Prognos*)) or (Decision* and ((Model* or Clinical*) or (Prognostic and (History or Variable* or Criteria or Scor* or Characteristic* or Finding* or Factor* or Model*))

S14	AB Validat* or Predict* or Rule* or (Predict* and (Outcome* or Risk* or Model*)) or ((History or Variable* or Criteria or Scor* or Characteristic* or Finding* or Factor*) and (Predict* or Model* or Decision* or Identif* or Prognos*)) or (Decision* and ((Model* or Clinical*) or (Prognostic and (History or Variable* or Criteria or Scor* or Characteristic* or Finding* or Factor* or Model*))

S15	(MH "Models, Statistical+")

S16	S10 OR S11 OR S12 OR S13 OR S14 OR S15

S17	S9 AND S16

S18	(EM 20230101-20241212) OR (ZD "in process" AND RD 20230101-20241212)

S19	S17 AND S18

**EPISTEMONIKOS**

**Date run: 31/01/23**

**Date update run: 21/06/24**

For update, all searches limited by date added to database: From: 01/01/2023 To: 21/06/2024

**Search 1**

(title:(decubit* OR bedsore* OR bed-sore* OR pressure-ulcer* OR pressure-wound*) OR abstract:(decubit* OR bedsore* OR bed-sore* OR pressure-ulcer* OR pressure-wound*)) AND (title:(stratification OR "ROC curve" OR "ROC curves" OR "receiver operating characteristic" OR discriminat* OR c-statistic OR "Area under the curve" OR AUC OR Calibration OR indices OR algorithm OR multivaria* OR Validat* OR Predict* OR Rule* OR Risk* OR Model* OR Criteria OR Scor* OR Characteristic* OR Finding* OR Factor* OR Decision* OR Prognos* OR Index OR model OR prevent*) OR abstract:(stratification OR "ROC curve" OR "ROC curves" OR "receiver operating characteristic" OR discriminat* OR c-statistic OR "Area under the curve" OR AUC OR Calibration OR indices OR algorithm OR multivaria* OR Validat* OR Predict* OR Rule* OR Risk* OR Model* OR Criteria OR Scor* OR Characteristic* OR Finding* OR Factor* OR Decision* OR Prognos* OR Index OR model OR prevent*))
Limit by publication type: systematic review (106) (Update: 21) or broad synthesis (3) (Update: 0)

**Search 2**

(title:("pressure ulcer" OR "pressure ulcers" OR "pressure sore" OR "pressure sores" OR "pressure lesion" OR "pressure lesions" OR "pressure injury" OR "pressure injuries") OR abstract:("pressure ulcer" OR "pressure ulcers" OR "pressure sore" OR "pressure sores" OR "pressure lesion" OR "pressure lesions" OR "pressure injury" OR "pressure injuries")) AND (title:(stratification OR "ROC curve" OR "ROC curves" OR "receiver operating characteristic" OR discriminat* OR c-statistic OR "Area under the curve" OR AUC OR Calibration OR indices OR algorithm OR multivaria* OR Validat* OR Predict* OR Rule* OR Risk* OR Model* OR Criteria OR Scor* OR Characteristic* OR Finding* OR Factor* OR Decision* OR Prognos* OR Index OR model OR prevent*) OR abstract:(stratification OR "ROC curve" OR "ROC curves" OR "receiver operating characteristic" OR discriminat* OR c-statistic OR "Area under the curve" OR AUC OR Calibration OR indices OR algorithm OR multivaria* OR Validat* OR Predict* OR Rule* OR Risk* OR Model* OR Criteria OR Scor* OR Characteristic* OR Finding* OR Factor* OR Decision* OR Prognos* OR Index OR model OR prevent*))
Limit by publication type: systematic review (709) (Update: 129) or broad synthesis (35) (Update: 25)

**Search 3**

(title:("deep-tissue wound" OR "deep-tissue wounds" OR "deep-tissue ulcer" OR "deep-tissue ulcers" OR "deep-tissue sore" OR "deep-tissue sores" OR "deep-tissue lesion" OR "deep-tissue lesions" OR "deep-tissue injury" OR "deep-tissue injuries") OR abstract:("deep-tissue wound" OR "deep-tissue wounds" OR "deep-tissue ulcer" OR "deep-tissue ulcers" OR "deep-tissue sore" OR "deep-tissue sores" OR "deep-tissue lesion" OR "deep-tissue lesions" OR "deep-tissue injury" OR "deep-tissue injuries")) AND (title:(stratification OR "ROC curve" OR "ROC curves" OR "receiver operating characteristic" OR discriminat* OR c-statistic OR "Area under the curve" OR AUC OR Calibration OR indices OR algorithm OR multivaria* OR Validat* OR Predict* OR Rule* OR Risk* OR Model* OR Criteria OR Scor* OR Characteristic* OR Finding* OR Factor* OR Decision* OR Prognos* OR Index OR model OR prevent*) OR abstract:(stratification OR "ROC curve" OR "ROC curves" OR "receiver operating characteristic" OR discriminat* OR c-statistic OR "Area under the curve" OR AUC OR Calibration OR indices OR algorithm OR multivaria* OR Validat* OR Predict* OR Rule* OR Risk* OR Model* OR Criteria OR Scor* OR Characteristic* OR Finding* OR Factor* OR Decision* OR Prognos* OR Index OR model OR prevent*))
Limit by publication type: systematic review (0) (Update: 0) or broad synthesis (0) (Update: 0)

**Search 4**

(title:("deep tissue wound" OR "deep tissue wounds" OR "deep tissue ulcer" OR "deep tissue ulcers" OR "deep tissue sore" OR "deep tissue sores" OR "deep tissue lesion" OR "deep tissue lesions" OR "deep tissue injury" OR "deep tissue injuries") OR abstract:("deep tissue wound" OR "deep tissue wounds" OR "deep tissue ulcer" OR "deep tissue ulcers" OR "deep tissue sore" OR "deep tissue sores" OR "deep tissue lesion" OR "deep tissue lesions" OR "deep tissue injury" OR "deep tissue injuries")) AND (title:(stratification OR "ROC curve" OR "ROC curves" OR "receiver operating characteristic" OR discriminat* OR c-statistic OR "Area under the curve" OR AUC OR Calibration OR indices OR algorithm OR multivaria* OR Validat* OR Predict* OR Rule* OR Risk* OR Model* OR Criteria OR Scor* OR Characteristic* OR Finding* OR Factor* OR Decision* OR Prognos* OR Index OR model OR prevent*) OR abstract:(stratification OR "ROC curve" OR "ROC curves" OR "receiver operating characteristic" OR discriminat* OR c-statistic OR "Area under the curve" OR AUC OR Calibration OR indices OR algorithm OR multivaria* OR Validat* OR Predict* OR Rule* OR Risk* OR Model* OR Criteria OR Scor* OR Characteristic* OR Finding* OR Factor* OR Decision* OR Prognos* OR Index OR model OR prevent*))
Limit by publication type: systematic review (5) (Update: 1) or broad synthesis (2) (Update: 0)

**Search 5**

(title:("bed wound" OR "bed wounds" OR "bed ulcer" OR "bed ulcers" OR "bed sore" OR "bed sores" OR "bed lesion" OR "bed lesions" OR "bed injury" OR "bed injuries") OR abstract:("bed wound" OR "bed wounds" OR "bed ulcer" OR "bed ulcers" OR "bed sore" OR "bed sores" OR "bed lesion" OR "bed lesions" OR "bed injury" OR "bed injuries")) AND (title:(stratification OR "ROC curve" OR "ROC curves" OR "receiver operating characteristic" OR discriminat* OR c-statistic OR "Area under the curve" OR AUC OR Calibration OR indices OR algorithm OR multivaria* OR Validat* OR Predict* OR Rule* OR Risk* OR Model* OR Criteria OR Scor* OR Characteristic* OR Finding* OR Factor* OR Decision* OR Prognos* OR Index OR model OR prevent*) OR abstract:(stratification OR "ROC curve" OR "ROC curves" OR "receiver operating characteristic" OR discriminat* OR c-statistic OR "Area under the curve" OR AUC OR Calibration OR indices OR algorithm OR multivaria* OR Validat* OR Predict* OR Rule* OR Risk* OR Model* OR Criteria OR Scor* OR Characteristic* OR Finding* OR Factor* OR Decision* OR Prognos* OR Index OR model OR prevent*))
Limit by publication type: systematic review (14) (Update: 0) or broad synthesis (0) (Update: 1)

**Search 6**

(title:("bed bound" OR bed-bound OR bedridden OR bed-ridden OR "bed ridden") OR abstract:("bed bound" OR bed-bound OR bedridden OR bed-ridden OR "bed ridden")) AND (title:(stratification OR "ROC curve" OR "ROC curves" OR "receiver operating characteristic" OR discriminat* OR c-statistic OR "Area under the curve" OR AUC OR Calibration OR indices OR algorithm OR multivaria* OR Validat* OR Predict* OR Rule* OR Risk* OR Model* OR Criteria OR Scor* OR Characteristic* OR Finding* OR Factor* OR Decision* OR Prognos* OR Index OR model OR prevent*) OR abstract:(stratification OR "ROC curve" OR "ROC curves" OR "receiver operating characteristic" OR discriminat* OR c-statistic OR "Area under the curve" OR AUC OR Calibration OR indices OR algorithm OR multivaria* OR Validat* OR Predict* OR Rule* OR Risk* OR Model* OR Criteria OR Scor* OR Characteristic* OR Finding* OR Factor* OR Decision* OR Prognos* OR Index OR model OR prevent*))
Limit by publication type: systematic review (30) (Update: 3) or broad synthesis (1) (Update: 1)

**Search 7**

(title:(stratification OR "ROC curve" OR "ROC curves" OR "receiver operating characteristic" OR discriminat* OR c-statistic OR "Area under the curve" OR AUC OR Calibration OR indices OR algorithm OR multivaria* OR Validat* OR Predict* OR Rule* OR Risk* OR Model* OR Criteria OR Scor* OR Characteristic* OR Finding* OR Factor* OR Decision* OR Prognos* OR Index OR model OR prevent*) OR abstract:(stratification OR "ROC curve" OR "ROC curves" OR "receiver operating characteristic" OR discriminat* OR c-statistic OR "Area under the curve" OR AUC OR Calibration OR indices OR algorithm OR multivaria* OR Validat* OR Predict* OR Rule* OR Risk* OR Model* OR Criteria OR Scor* OR Characteristic* OR Finding* OR Factor* OR Decision* OR Prognos* OR Index OR model OR prevent*)) AND (title:( wound* OR ulcer* OR sore* OR injur* OR lesion*) OR abstract:( wound* OR ulcer* OR sore* OR injur* OR lesion*)) AND (title:(supine OR immobil*) OR abstract:(supine OR immobil*))
Limit by publication type: systematic review (234) (Update: 39) or broad synthesis (13) (Update: 3)

**Search 8**

(title:(stratification OR "ROC curve" OR "ROC curves" OR "receiver operating characteristic" OR discriminat* OR c-statistic OR "Area under the curve" OR AUC OR Calibration OR indices OR algorithm OR multivaria* OR Validat* OR Predict* OR Rule* OR Risk* OR Model* OR Criteria OR Scor* OR Characteristic* OR Finding* OR Factor* OR Decision* OR Prognos* OR Index OR model OR prevent*) OR abstract:(stratification OR "ROC curve" OR "ROC curves" OR "receiver operating characteristic" OR discriminat* OR c-statistic OR "Area under the curve" OR AUC OR Calibration OR indices OR algorithm OR multivaria* OR Validat* OR Predict* OR Rule* OR Risk* OR Model* OR Criteria OR Scor* OR Characteristic* OR Finding* OR Factor* OR Decision* OR Prognos* OR Index OR model OR prevent*)) AND (title:(heal OR healing OR heals OR healed OR dress*) OR abstract:(heal OR healing OR heals OR healed OR dress*)) AND (title:(supine OR immobil*) OR abstract:(supine OR immobil*))
Limit by publication type: systematic review (39) (Update: 9) or broad synthesis (3) (Update: 0)

**GOOGLE SCHOLAR 1/02/23**

**Update run: 24/06/24**, limit to years 2023-2024

allintitle: prevent OR prevention OR risk OR predict OR prevents OR risks OR prediction OR predicts OR prognosis OR prognostic "pressure injury" -ulcer -ulcers Limit to review and years 2013-2023 (66)

(Update: 31)

allintitle: prevent OR prevention OR risk OR predict OR prevents OR risks OR prediction OR predicts OR prognosis OR prognostic "pressure injuries" -ulcer -ulcers Limit to review and years 2013-2023 (33)

(Update: 14)

allintitle: prevent OR prevention OR risk OR predict OR prevents OR risks OR prediction OR predicts OR prognosis OR prognostic "pressure ulcer” Limit to review and years 2013-2023 (120)

(Update: 14)

allintitle: prevent OR prevention OR risk OR predict OR prevents OR risks OR prediction OR predicts OR prognosis OR prognostic "pressure ulcers” Limit to review and years 2013-2023 (102)

(Update: 14)

allintitle: prevent OR prevention OR risk OR predict OR prevents OR risks OR prediction OR predicts OR prognosis OR prognostic "pressure sore” Limit to review and years 2013-2023 (3)

(Update: 0)

allintitle: prevent OR prevention OR risk OR predict OR prevents OR risks OR prediction OR predicts OR prognosis OR prognostic "pressure sores” Limit to review and years 2013-2023 (2)

(Update: 0)

allintitle: prevent OR prevention OR risk OR predict OR prevents OR risks OR prediction OR predicts OR prognosis OR prognostic "pressure wound” Limit to review and years 2013-2023 (25)

(Update: 4)

allintitle: prevent OR prevention OR risk OR predict OR prevents OR risks OR prediction OR predicts OR prognosis OR prognostic "pressure wounds” Limit to review and years 2013-2023 (0)

(Update: 0)

allintitle: prevent OR prevention OR risk OR predict OR prevents OR risks OR prediction OR predicts OR prognosis OR prognostic "bedsore” Limit to review and years 2013-2023 (1)

(Update: 0)

allintitle: prevent OR prevention OR risk OR predict OR prevents OR risks OR prediction OR predicts OR prognosis OR prognostic "bedsores” Limit to review and years 2013-2023 (1)

(Update: 1)

allintitle: prevent OR prevention OR risk OR predict OR prevents OR risks OR prediction OR predicts OR prognosis OR prognostic "bed sore” Limit to review and years 2013-2023 (0)

(Update: 0)

allintitle: prevent OR prevention OR risk OR predict OR prevents OR risks OR prediction OR predicts OR prognosis OR prognostic "bed sores” Limit to review and years 2013-2023 (0)

(Update: 0)

allintitle: prevent OR prevention OR risk OR predict OR prevents OR risks OR prediction OR predicts OR prognosis OR prognostic "decubitus” Limit to review and years 2013-2023 (4)

(Update: 2)

### Appendix 3: Data extraction form

| **Data Extraction Items** | | | |
| --- | --- | --- | --- |
|  | Extractor | | |
| **Publication information:** | Review Title;  First Author;  Publication Year;  Umbrella review eligibility (D/V, ACC, CE);  Comments;  Primary studies fundings reported?^A^;  Conflicts of Interest reported?^A^ | | |
| **Eligibility Criteria:** | Population;  Setting;  Prediction models/tools;  Model outcome (and classification if specified);  Interventions^B^;  Comparators^B^;  Outcomes of interest^B^;  Inclusion criteria incorporated PICO, PIRT or POII?^A^;  Source of data (prospective/retrospective);  Phase of development of models;  Study design;  Did they explain reasons for study design inclusions?^A^;  Exclusion criteria | | |
| **Review methods:** | Review protocol;  Protocol and justifications for deviations from?^A^;  Databases searched;  Adequate search strategy?^A^;  Search cut-off date;  Publication restrictions;  Quality assessment tool;  Suitable quality assessment tool?;  Study selection method;  Study selection in duplicate?^A^;  Quality assessment method;  Data extraction method;  Data extraction in duplicate?^A^;  Synthesis method;  Appropriate method of statistical synthesis, if applicable?^A^ | | |
| **Review results:** | PRISMA diagram provided?;  Excluded studies list (with justifications)?^A^;  N models per review;  N studies per review;  N participants in review;  How were the results presented? (e.g. outcomes reported);  Description of included studies provided? (summary table, tabulated per study, narrative only);  Description of included studies adequate?^A^;  Study quality described? (summary table, tabulated per study, narrative only);  Assessment of RoB satisfactory?^A^;  Assessment of impact of RoB on synthesised results?^A^;  Assessment of impact of RoB on review results?^A^;  Discussion/investigation of heterogeneity?^A^;  Models included;  Brief description of included studies;  Brief description of study quality | | |
| **D/V Reviews**  **(Re: prognostic studies)** | | **Accuracy Reviews**  **(Re: accuracy studies)** | **Clinical Effectiveness Reviews**  **(Re: effectiveness studies)** |
|  | | 2x2 tables presented for each study? |  |
|  | | Cut-off points specified for each study? |  |
|  | | List Author, year of primary studies included in review |  |
| Summary estimates:  Overall model performance (e.g. R-squared, Brier score)  Model calibration (e.g. calibration plot, slope, intercept, O/E ratio)  Model discrimination (e.g. c-statistic, AUC)  (results from statistical synthesis) | | Summary estimates:  Sensitivity (incl. n), specificity (incl. N), likelihood ratios, DOR, AUROC, predictive values  Summary Sensitivity (incl. n)  (results from statistical synthesis) | Summary of statistical synthesis of results  (e.g. effect on incidence of PI, treatment outcome, or other patient-relevant outcomes) |
| Summary of narrative synthesis of results | | Summary of narrative synthesis of results | Summary of narrative synthesis of results |
| AUC – area under the curve; AUROC – area under the receiver operating characteristic curve; AMSTAR – A MeaSurement Tool to Assess systematic Reviews; CE – clinical effectiveness; DOR – diagnostic odds ratio; ACC – test accuracy; D/V – development/validation; O/E – observed/expected; PI – pressure injury; PICO – population, intervention, comparator, outcome; PIRT – population, index test, reference standard, target condition; POII – population, outcome, intended use, intended timing; PRISMA – Preferred Reporting Items for Systematic Reviews and Meta-Analyses; RoB – risk of bias.  ^A^ AMSTAR-2 Items.  ^B^ applicable to clinical effectiveness reviews only. | | | |

### Appendix 4: AMSTAR-2 Methodology Quality Appraisal. Adapted for application to reviews of prognostic model and accuracy studies.

|  | AMSTAR-2 Adapted | |
| --- | --- | --- |
|  | Questions | Guidance |
| Item 1. | 1. Did the research questions and inclusion criteria for the review include the components one of the following: PICO, PIRT, or POII? **Y/N** | For intervention reviews: Population, Intervention, Comparator, Outcome  For prognostic accuracy reviews: Population, Index test, Reference standard, Target condition (PIRT) Population, Outcome to be predicted, Intended use of model, Intended moment in time (POII) |
| **Item 2*** | 2. Did the report of the review contain an explicit statement that the review methods were established prior to the conduct of the review and did the report justify any significant deviations from the protocol? **Y/PY/N** | For Partial Yes (PY): The authors state that they had a written protocol or guide that included ALL the following:   - review question(s), - a search strategy, - inclusion/exclusion criteria, - a risk of bias assessment.   For Yes: As for partial yes, plus the protocol should be registered and should also have specified:   - a meta-analysis/synthesis plan, if appropriate, - and a plan for investigating causes of heterogeneity, - justification for any deviations from the protocol. |
| Item 3. | 3. Did the review authors explain their selection of the study designs for inclusion in the review? **Y/N** | For Yes, the review should give an explanation for including types of studies included in the review, for example:  For the development/validation review: development studies, validation studies or both.  For the accuracy/effectiveness review: single group (prospective/retrospective), two/multi group (i.e. diagnostic case-control), RCTs, NSRs |
| **Item 4*** | 4. Did the review authors use a comprehensive literature search strategy? **Y/PY/N** | For Partial Yes (all the following):   - searched at least 2 databases (relevant to research question), - provided key word and/or search strategy, - justified publication restrictions (e.g. language).   For Yes, should also have (all the following):   - searched the reference lists / bibliographies of included studies, - searched trial/study registries, - included/consulted content experts in the field where relevant, - searched for grey literature, - conducted search within 24 months of completion of the review. |
| Item 5. | 5. Did the review authors perform study selection in duplicate? **Y/N** | For Yes, either ONE of the following: at least two reviewers independently agreed on selection of eligible studies and achieved consensus on which studies to include, OR two reviewers selected a sample of eligible studies and achieved good agreement (at least 80 percent), with the remainder selected by one reviewer. |
| Item 6. | 6. Did the review authors perform data extraction in duplicate? **Y/N** | For Yes, either ONE of the following: at least two reviewers achieved consensus on which data to extract from included studies, OR two reviewers extracted data from a sample of eligible studies and achieved good agreement (at least 80 percent), with the remainder extracted by one reviewer. |
| **Item 7*** | 7. Did the review authors provide a list of excluded studies and justify the exclusions? **Y/PY/N** | For Partial Yes: provided a list of all potentially relevant studies that were read in full-text form but excluded from the review  For Yes, must also have: Justified the exclusion from the review of each potentially relevant study |
| Item 8. | 8. Did the review authors describe the included studies in adequate detail? **Y/PY/N** | For Partial Yes (ALL the following per included study):   - described PICO/PIRT/POII (whichever applicable), - and described research designs   For Yes, should also have ALL the following per included study:   - described PICO/PIRT/POII (whichever applicable) in detail, - described study’s setting - and timeframe for follow-up |
| **Item 9*** | 9. Did the review authors use a satisfactory technique for assessing the risk of bias (RoB) in  individual studies that were included in the review? **Y/PY/N** | **RCTs** For Partial Yes, must have reported summary findings and assessed RoB from:   - unconcealed allocation, - and lack of blinding of patients and assessors when assessing outcomes (unnecessary for objective outcomes such as all-cause mortality)   For Yes, must also have given itemisation of quality judgements per study, and assessed RoB from:   - allocation sequence that was not truly random, - and selection of the reported result from among multiple measurements or analyses of a specified outcome |
|  |  | **NRS** For Partial Yes, must have reported summary findings and assessed RoB from:   - confounding, - and from selection bias   For Yes, must also have given itemisation of quality judgements per study, and assessed RoB from:   - methods used to ascertain exposures and outcomes, - and selection of the reported result from among multiple measurements or analyses of a specified outcome |
|  |  | **Accuracy studies** For Partial Yes, must have assessed RoB with a recognised tool (e.g. QUADAS-2) and given summary of result across domains  For Yes, must also have also given itemisation of quality judgements per study. |
|  |  | **Prognostic studies** For Partial Yes, must have assessed RoB with a recognised tool (e.g. PROBAST, QUIPS) and given summary of result across domains  For Yes, must also have also given itemisation of quality judgements per study. |
| Item 10. | 10. Did the review authors report on the sources of funding for the studies included in the review? **Y/N** | For Yes: Must have reported on the sources of funding for individual studies included in the review. Note: Reporting that the reviewers looked for this information, but it was not reported by study authors also qualifies |
| **Item 11*** | 11. If meta-analysis was performed did the review authors use appropriate methods for statistical combination of results? **Y/N/ 'No MA conducted'** | For Yes: The authors justified combining the data in a meta-analysis AND they used an appropriate weighted technique to combine study results and adjusted for heterogeneity if present. AND investigated the causes of any heterogeneity |
| Item 12. | 12. If meta-analysis was performed, did the review authors assess the potential impact of RoB in individual studies on the results of the meta-analysis or other evidence synthesis? **Y/N/ 'No MA conducted'** | For Yes: included only low risk of bias studies OR, if the pooled estimate was based on studies at variable RoB, the authors performed sensitivity analyses to investigate possible impact of RoB on summary estimates |
| **Item 13*** | 13. Did the review authors account for RoB in individual studies when interpreting/ discussing the results of the review? **Y/N** | For Yes: included only low risk of bias RCTs OR, if RCTs with moderate or high RoB, or NRSs were included the review provided a discussion of the likely impact of RoB on the results |
| Item 14. | 14. Did the review authors provide a satisfactory explanation for, and discussion of, any heterogeneity observed in the results of the review? **Y/N** | For Yes: There was no significant heterogeneity, OR if heterogeneity was present, the authors performed an investigation of main sources of heterogeneity in the results, if applicable, and particularly any between-study heterogeneity and discussed the impact of this |
| Item 15. | 15. Did the review authors report any potential sources of conflict of interest, including any funding they received for conducting the review? **Y/N** | For Yes: The authors reported no competing interests, OR the authors described their funding sources and how they managed potential conflicts of interest |
|  | * Critical domains identified by AMSTAR-2 developers.^1^  DEV/VAL – development/validation; MA – Meta-Analysis; N – No; NRS – non-randomised study; PICO – population, intervention, comparator, outcome; PIRT – population, index test, reference standard, target condition; POII – population, outcome, intended use, intended time; PROBAST – Prediction model Risk Of Bias ASsessment Tool; PY – Partial Yes; QUADAS – Quality Assessment of Diagnostic Accuracy Studies; QUIPS – Quality In Prognosis Studies; RCT – randomised controlled trial; RoB – risk of bias; Y – Yes. | |

### Appendix 5: Detailed results tables

#### Table S1. Full-text articles excluded, with reasons

|  | **Author, year** | **Title** | **Major reason for exclusion** |
| --- | --- | --- | --- |
| 1 | Alves, 2014 ^2^ | *Assessment of risk for pressure ulcers in intensive care units: an integrative review* | Not a systematic review |
| 2 | Anthony, 2008 ^3^ | *Norton, Waterlow and Braden scores: a review of the literature and a comparison between the scores and clinical judgement* | Not a systematic review |
| 3 | Barradas Cavalcante, 2016 ^4^ | *Updating pf the assistance protocol for pressure prevention: evidence based practice* | Not a systematic review |
| 4 | Charalambous, 2018 ^5^ | *Evaluation of the Validity and Reliability of the Waterlow Pressure Ulcer Risk Assessment Scale* | Not a systematic review |
| 5 | de Laat, 2006 ^6^ | *Epidemiology, risk and prevention of pressure ulcers in critically ill patients: a literature review* | Not a systematic review |
| 6 | do Egito Cavalcanti de Farias, 2022 ^7^ | *Risk factors for the development of pressure injury in the elderly: integrative review* | Not a systematic review |
| 7 | Feuchtinger, 2005 ^8^ | *Pressure ulcer risk factors in cardiac surgery: A review of the research literature* | Not a systematic review |
| 8 | Garcia-Fernandez, 2014 ^9^ | *A new theoretical model for the development of pressure ulcers and other dependence-related lesions* | Not a systematic review |
| 9 | Garrubba, 2017 ^10^ | *Effectiveness of the Braden risk screening tool for pressure injuries: systematic review* | Not a systematic review |
| 10 | Kelechi, 2013 ^11^ | *Review of pressure ulcer risk assessment scales* | Not a systematic review |
| 11 | Keller, 2002 ^12^ | *Pressure ulcers in intensive care patients: A review of risks and prevention* | Not a systematic review |
| 12 | Ladd, 2018 ^13^ | *A systematic review of pressure ulcers in burn patients: Risk factors, demographics, and treatment modalities* | Not a systematic review |
| 13 | Lepisto, 2006 ^14^ | *Developing a Pressure Ulcer Risk Assessment Scale for Patients in Long-Term Care* | Not a systematic review |
| 14 | Mendes Coqueiro, 2013 ^15^ | *Multiple risk factors and preventive strategies of pressure ulcers: systematic review* | Not a systematic review |
| 15 | Michel, 2012 ^16^ | *As of 2012, what are the key predictive risk factors for pressure ulcers? Developing French guidelines for clinical practice* | Not a systematic review |
| 16 | Ming, 2012 ^17^ | *Systematic review of pressure ulcer risk assessment scales for using in ICU patients* | Not a systematic review |
| 17 | Mordiffi, 2010 ^18^ | *Evaluating the effects of using the mobility assessment sub-scale within the Braden Scale on pressure ulcer incidence and preventive interventions in adult acute care settings: A systematic review* | Not a systematic review |
| 18 | Mortenson, 2008 ^19^ | *A review of scales for assessing the risk of developing a pressure ulcer in individuals with SCI* | Not a systematic review |
| 19 | Nadeem, 2021 ^20^ | *Utility of the Waterlow scale in acute care settings: A literature review* | Not a systematic review |
| 20 | O’Tuathail, 2011 ^21^ | *Evaluation of three commonly used pressure ulcer risk assessment scales* | Not a systematic review |
| 21 | Rodriguez Torres, 2007 ^22^ | *Clinical judgement or assessment scales to identify patients at risk of developing pressure ulcers?* | Not a systematic review |
| 22 | Sales de Almeida, 2020 ^23^ | *Pressure injury prevention scales in intensive care units: an integrative review* | Not a systematic review |
| 23 | Santos, 2015 ^24^ | *Development of the nursing diagnosis risk for pressure ulcer* | Not a systematic review |
| 24 | Satekova, 2014 ^25^ | *Validity of pressure ulcer risk assesment scales: Review* | Not a systematic review |
| 25 | Shahin, 2007 ^26^ | *Predictive validity of pressure ulcer risk assessment tools in intensive care patients* | Not a systematic review |
| 26 | Smet, 2019 ^27^ | *The Belgian pressure ulcer risk assessment project: Is assessing mobility and skin status a more accurate, reliable, and feasible approach to assess pressure ulcer risk in hospitalised patients?* | Not a systematic review |
| 27 | Solati, 2016 ^28^ | *Predictive values of Braden and Waterlow scales to assess the risk of pressure ulcer* | Not a systematic review |
| 28 | Taylor, 1988 ^29^ | *Assessment tools for the identification of patients at risk for the development of pressure sores: a review* | Not a systematic review |
| 29 | Tran, 2016 ^30^ | *Prevention of Pressure Ulcers in the Acute Care Setting: New Innovations and Technologies* | Not a systematic review |
| 30 | Tschannen, 2020 ^31^ | *The pressure injury predictive model: A framework for hospital-acquired pressure injuries* | Not a systematic review |
| 31 | Walsh, 2011 ^32^ | *Investigating the reliability and validity of the Waterlow risk assessment scale: A literature review* | Not a systematic review |
| 32 | Xu, 2018 ^33^ | *Risk assessment tools for pressure injury in intensive care patients: a review* | Not a systematic review |
| 33 | Alderden, 2017 ^34^ | *Risk factors for pressure injuries among critical care patients: A systematic review* | No risk prediction models |
| 34 | Barbosa da Silva, 2020 ^35^ | *Pressure ulcers in individuals with spinal cord injury: risk factors in neurological rehabilitation* | No risk prediction models |
| 35 | Di Prinzio, 2019 ^36^ | *Risk factors for the development and recurrence of pressure ulcers in patients with spinal cord injury: A systematic review* | No risk prediction models |
| 36 | Haisley, 2020 ^37^ | *Postoperative pressure injuries in adults having surgery under general anaesthesia: systematic review of perioperative risk factors* | No risk prediction models |
| 37 | Ham, 2014 ^38^ | *Pressure ulcers from spinal immobilization in trauma patients: A systematic review* | No risk prediction models |
| 38 | Lima, 2021 ^39^ | *Risk factors and preventive interventions for pressure injuries in cancer patients* | No risk prediction models |
| 39 | Lima Serrano, 2017 ^40^ | *Risk factors for pressure ulcer development in Intensive Care Units: Systematic review* | No risk prediction models |
| 40 | Marin, 2013 ^41^ | *A systematic review of risk factors for the development and recurrence of pressure ulcers in people with spinal cord injuries* | No risk prediction models |
| 41 | Rao, 2016 ^42^ | *Risk Factors Associated With Pressure Ulcer Formation in Critically Ill Cardiac Surgery Patients: A Systematic Review* | No risk prediction models |
| 42 | Reenalda, 2009 ^43^ | *Clinical use of interface pressure to predict pressure ulcer development: a systematic review* | No risk prediction models |
| 43 | Shi, 2018 ^44^ | *Skin status for predicting pressure ulcer development: A systematic review and meta-analyses* | No risk prediction models |
| 44 | Siping, 2022 ^45^ | *Risk factors of intraoperative acquired pressure injury: A systematic review and meta-analysis* | No risk prediction models |
| 45 | Wynn, 2022 ^46^ | *Risk factors for the development and evolution of deep tissue injuries: A systematic review* | No risk prediction models |
| 46 | Zhang, 2022 ^47^ | *Prevalence and Risk Factors of Postoperative Pressure Ulcers: A Systematic Review and Meta-analysis of Diagnostic Test* | No risk prediction models |
| 47 | Bulfone, 2018 ^48^ | *Perioperative Pressure Injuries: A Systematic Literature Review* | Wrong research question |
| 48 | Chung, 2022 ^49^ | *Risk Factors for Pressure Injuries in Adult Patients: A Narrative Synthesis* | Wrong research question |
| 49 | Chung, 2022 ^50^ | *Risk factors for pressure ulcers in adult patients: A meta-analysis on sociodemographic factors and the Braden scale* | Wrong research question |
| 50 | Coleman, 2013 ^51^ | *Patient risk factors for pressure ulcer development: Systematic review* | Wrong research question |
| 51 | Dube, 2022 ^52^ | *Risk factors associated with heel pressure ulcer development in adult population: A systematic literature review* | Wrong research question |
| 52 | Ferris, 2019 ^53^ | *Pressure ulcers in patients receiving palliative care: A systematic review* | Wrong research question |
| 53 | Floyd, 2018 ^54^ | *Effectiveness of pressure ulcer protocols with the Braden Scale for elderly patients in the intensive care unit: A Systematic Review* | Wrong research question |
| 54 | Gelis, 2009 ^55^ | *Pressure ulcer risk factors in persons with SCI: Part I: Acute and rehabilitation stages* | Wrong research question |
| 55 | Gelis, 2009 ^56^ | *Pressure ulcer risk factors in persons with spinal cord injury part 2: the chronic stage* | Wrong research question |
| 56 | Liu, 2024 ^57^ | *Effects of predictive nursing interventions on pressure*  *ulcer in older bedridden patients: A meta-analysis* | Wrong research question |
| 57 | Moore, 2023 ^58^ | *A systematic review of movement monitoring devices to aid the prediction of pressure ulcers in at-risk adults* | Wrong research question |
| 58 | Mordiffi, 2011 ^59^ | *Use of mobility subscale for risk assessment of pressure ulcer incidence and preventive interventions: A systematic review* | Wrong research question |
| 59 | Nixon, 2015 ^60^ | *Pressure UlceR Programme Of reSEarch (PURPOSE): using mixed methods (systematic reviews, prospective cohort, case study, consensus and psychometrics) to identify patient and organisational risk, develop a risk assessment tool and patient-reported outcome Quality of Life and Health Utility measures* | Wrong research question |
| 60 | Richardson, 2015 ^61^ | *Part 1: Pressure ulcer assessment - the development of Critical Care Pressure Ulcer Assessment Tool made Easy (CALCULATE)* | Wrong research question |
| 61 | Teixeira, 2022 ^62^ | *Risk factors for pressure injury in critically ill polytraumatized patients: A systematic review* | Wrong research question |
| 62 | Ting, 2021 ^63^ | *E-Health Decision Support Technologies in the Prevention and Management of Pressure Ulcers: A Systematic Review* | Wrong research question |
| 63 | Toffaha, 2023 ^64^ | *Leveraging artificial intelligence and decision support systems in hospital-acquired pressure injuries prediction: A comprehensive review* | Wrong research question |
| 64 | Fuentelsaz Gallego, 2005 ^65^ | *Review of literature on pressure ulcers in people aged 65 or over* | No English language translation |
| 65 | Garcia-Fernandez, 2013 ^66^ | *Risk assessment scales for pressure ulcer in intensive care units: A systematic review with metaanalysis* | No English language translation |
| 66 | Kottner, 2008 ^67^ | *Interrater reliability of the Braden scale* | No English language translation |
| 67 | Nunes de Sousa, 2023 ^68^ | *SCALES USED TO MEASURE PRESSURE INJURY RISK IN HOSPITALIZED PATIENTS: A REVIEW* | No English language translation |
| 68 | Pancorbo-Hidalgo, 2008 ^69^ | *Pressure ulcers risk assessment: clinical practice in Spain and a meta-analysis of scales effectiveness* | No English language translation |
| 69 | Park, 2014 ^70^ | *Predictive validity of the Braden Scale for pressure ulcer risk: a meta-analysis* | No English language translation |
| 70 | Yang, 2019 ^71^ | *Predictive validity of the Munro Scale for pressure injuries in surgical patients: A meta-analysis* | No English language translation |
| 71 | De Queiroz, 2022 ^7^ | *Risk factors for the development of pressure injury in the elderly: integrative review/Fatores de risco o para desenvolvimento de lesão por pressão em idosos: revisão integrativa* | Duplicate |
| 72 | Garcia-Fernandez, 2013 ^66^ | *Risk assessment scales for pressure ulcers in intensive care units: A systematic review with meta-analysis* | Duplicate |
| 73 | Nixon, 2015 ^60^ | *Pressure UlceR Programme Of reSEarch (PURPOSE): using mixed methods (systematic reviews, prospective cohort, case study, consensus and psychometrics) to identify patient and organisational risk, develop a risk assessment tool and patient-reported outcome Quality of Life and Health Utility measures* | Duplicate |
| 74 | Nayar, 2021 ^72^ | *Waterlow score for risk assessment in surgical patients: a systematic review* | Wrong outcome |
| 75 | Zahia, 2020 ^73^ | *Pressure injury image analysis with machine learning techniques: A systematic review on previous and possible future methods* | Wrong outcome |
| 76 | Moore, 2008 ^74^ | *Risk assessment tools for the prevention of pressure ulcers* | Updated version included |
| 77 | Moore, 2014 ^75^ | *Risk assessment tools for the prevention of pressure ulcers* | Updated version included |
| 78 | Liao, 2018 ^76^ | *Predictive accuracy of the Braden Q Scale in risk assessment for paediatric pressure ulcer: A meta-analysis* | Wrong population |
| 79 | Ribeiro, 2013 ^77^ | *How effective is the development of skin care in critically ill patients using the Braden Scale scores aiming to prevent the incidence of pressure ulcers? Sistematic Literature Review* | No results |

#### Table S2. Systematic review characteristics

| **Review author**  (publication year)  **Review question** | **Eligibility criteria** | | | **Review methods** | | | | | **Volume of evidence** | |
| --- | --- | --- | --- | --- | --- | --- | --- | --- | --- | --- |
|  | **Population; setting** | **Prediction tools; PI classification system** | **Study design** | **Databases searched** | **Publication restrictions**  Year; language; publication type | **Quality assessment tool** | **Meta-analysis included; method of meta-analysis** | **N relevant studies in review**  (n participants) | | **N tools included** |
| Barghouthi^78^ (2023)  Model development | “Adult” inpatients (age ≥14y); hospital | ML; NS | NS | CINAHL; PubMed; Science Direct; IEEE; Cochrane; Google Scholar | 2017-2023; English; NS | JBI appraisal checklist for cohort studies | No | 23 (706393) | | 23 |
| Baris^79^ (2015)  Effectiveness | Turkish populations only; NS | Braden; NS | NS | Turkish MEDLINE; PubMed; ScienceDirect; Google Scholar; YOK Thesis Search; Reference Directory of Turkey; Medicine Directory of Turkish Clinics; ULAKBIM National Database; National Library Bibliography of Turkish Articles | 1998-2012; English, Turkish; NS | None | No | 16 (2273^a^) | | 2 |
| Chen^80^ (2023)  Accuracy | Patients with a critical illness; ICU | Cubbin & Jackson; NS | Diagnostic studies (presenting TP, FP, TN and FN results) with any research design | EBSCO; PubMed; Ovid; Web of Science; Cochrane databases; Wangfang Data; China National  Knowledge Infrastructure | Database inception - 2021; NS; reviews and expert opinions excluded | QUADAS-II | Yes; meta-analysis method unclear, SROC analysis | 9 (7684) | | 1 |
| Chen^81^ (2016)  Accuracy | NS; long-term care | Braden; NS | NS | PubMed; Web of Science | Inception-2015; English; NS | QUADAS | Yes; DerSimonian and Laird random-effects model, SROC analysis | 8 (41489) | | 1 |
| Chou^82^ (2013)  Accuracy  Effectiveness | Adults (age ≥18y); acute care hospital, long-term and rehabilitation facilities, operative and postoperative, community (home care and wheelchair users) | PI risk assessment tools; NS | KQ1^b^: controlled or comparative randomised and nonrandomised trials, controlled or comparative observational studies KQ2^b^: prospective studies of predictive validity (case-control excluded) | MEDLINE; CINAHL; Cochrane Library; grant databases; clinical trial registries | 1946-2021 (MEDLINE), 1988-2012 (CINAHL), inception- 2012 (Cochrane library); English; conference abstracts excluded | Criteria consistent with AHRQ Methods Guide for Effectiveness and Comparative Effectiveness Reviews | No; presented median accuracy results | KQ1^b^: 3 KQ2^b^: 47 | | KQ1^b^: 4 KQ2^b^: 20 |
| Dweekat^83^ (2023)  Model development | NS; NS | ML; NS | NS | PubMed; Web of Science; Scopus; Science Direct | 2007-2022; English; article papers, review papers, conference proceedings | None | No | 35 (664719) | | 35 |
| Garcia-Fernandez^84^ (2014)  Accuracy | No PIs at baseline, no age restriction; NS | PI risk assessment tools; NS | Controlled clinical trials, prospective cohort | Cochrane Library; Center for Reviews and Dissemination University of York; LILACS; CUIDEN Plus; Spanish Medical Index | 1962-2010; no restriction; peer-reviewed journal article | CASP for RCT/cohort studies | Yes; random-effects model | 70 (30327) | | 28 |
| Gaspar^85^ (2019)  Effectiveness | Adult inpatients; hospital wards or any acute unit | PI prevention strategies; NS | Prospective or retrospective; cross-sectional, comparative, pre-test and post-test, quasi-experimental, experimental, RCT, mixed-method | MEDLINE; CINAHL; PubMed; Web of Science; EBSCO Nursing & Allied Health; Cochrane Central Register of Controlled Trials; Library, Information Science & Technology Abstracts; MedicLatina | 2009-2018; English, French, Portuguese, Spanish; peer-reviewed | Evidence-Based Librarianship Critical Appraisal checklist | No | 1 (1231) | | 2 |
| He^86^ (2012)  Accuracy | NS; surgical | Braden; NS | Studies assessing predictive validity | PubMed; Web of Science | Not stated-2011; NS; NS | QUADAS | Yes; DerSimonian and Laird random-effects model, SROC analysis | 3 (609) | | 1 |
| Health Quality Ontario^87^ (2009)  Effectiveness | Any population at risk of developing PIs; NS | PI risk assessment tools; NS | Systematic reviews, RCTs, non-randomised controlled clinical trials | MEDLINE; MEDLINE In-Process; CINAHL; EMBASE; Cochrane Library; other non-indexed citations | 1997-2008; English; NS | Criteria name not given | No | 3 (528) | | 3 |
| Huang^88^ (2021)  Accuracy | Inpatients aged ≥18y, no PIs at admission; NS | Braden; accepted standards (NPUAP, EPUAP, AHCPR, ICD-9, Bergstrom, others) | Cross-sectional, cohort | PubMed; CINAHL; EMBASE; Web of Science; Cochrane Library; bibliographies | Inception-2020; NS; NS | QUADAS-II | Yes; bivariate model, SROC analysis | 60 (49326) | | 1 |
| Jiang^89^ (2021)  Development | Any population; NS | ML; NS | NS | CINAHL; PubMed; EMBASE; Web of Science; Cochrane Library; China National Knowledge Infrastructure; Wanfang database; VIP database; China Biomedical Literature Database | NS-2020; English, Chinese; review papers, opinion papers, editorials, discussion papers, dissertations, conference abstracts excluded | PROBAST | No | 9 (1278148) | | 9 |
| Kottner^90^ (2009)  Effectiveness (reliability) | NS; NS | Waterlow; NS | Inter- and intrarater reliability and agreement | MEDLINE; EMBASE; CINAHL | 1985-2008; English, German; original research | Own criteria | No | 8 | | 2 |
| Lovegrove^91^ (2021)  Effectiveness | Adults (age ≥18y); acute hospital care | PI risk assessment tools; NS | Primary research | MEDLINE; EMBASE; EBSCO CINAHL; EBSCO; Scopus; Web of Science | 2010-2020; English; conference abstracts, posters excluded | JBI tools  or analytical cross-sectional study appraisal checklist | No | 5 (1910) | | 5 |
| Lovegrove^92^ (2018)  Effectiveness | Adults; hospital or acute care | PI risk assessment tools; NS | Primary research | MEDLINE; CINAHL; Scopus; Web of Science | 2007-2017; English; non-research publications excluded | JBI tools | No | 20 | | 5^b^ |
| Mehicic^93^ (2024)  Accuracy  Effectiveness (reliability, measurement error and convergent validity) | Adults (age ≥18y); ICU  For reliability assessment: sample of nurse-raters required | Braden; NS | Primary quantitative or mixed-methods research studies | CINAHL; EMBASE; MEDLINE; Scopus; Web of Science | Database inception - 2023; English language; Peer-reviewed | COSMIN RoB checklist | No | 34 (59325) | | 1 |
| Moore^94^ (2019)  Effectiveness | People without PIs, any age; any healthcare setting | PI risk assessment tools; validated PI staging system | RCTs or cluster-RCTs | MEDLINE; EMBASE; CINAHL; Cochrane Wounds Specialised Register; Cochrane Central Register of Controlled Trials | Start date between 1937-1974, until 2018; no restrictions; no restrictions | Cochrane RoB tool | No | 2 (1487) | | 3 |
| Pancorbo-Hidalgo^95^ (2006)  Accuracy  Effectiveness | No PIs at baseline; NS | PI risk assessment tools; NS | Controlled clinical trials, prospective cohort | MEDLINE; CINAHL; EBSCO; ScienceDirect; Current contents; DARE; Indice medico espanol; LILACS; CUIDEN; Cochrane Library; Springer; InterSciencia; ProQuest; Pascal | 1966-2003; Spanish, English, French, Portuguese; no restrictions | CASP Guide for clincial trials; critical assessment guide for PI assessment and prevention for cohort studies | Yes; weighted average values using inverse of variance for weights, DerSimonian and Laird random-effects model | 33 | | 13 |
| Park^96^ (2016a)  Accuracy | NS; NS | Modified Braden, Waterlow, Norton, Cubbin & Jackson; NPUAP, EPUAP, AHCPR, Torrence Developmental Classification of Pressure Sore | NS | MEDLINE; EMBASE; CINAHL; Cochrane Library; KoreaMed; NDSL; KERIS | NS-2013; NS; NS | QUADAS-II | Yes; random-effects model, SROC analysis | 17 (6143) | | 5 |
| Park^97^ (2016b)  Accuracy | Elderly (age ≥60y); NS | Braden, Waterlow, Norton; NS | NS | MEDLINE; EMBASE; CINAHL; Cochrane database; KoreaMed | 1966-2013; NS; NS | QUADAS-II | Yes; random-effects model, SROC analysis | 29 (11729) | | 3 |
| Park^98^ (2015)  Accuracy | Adults (age ≥18y) with no PIs at baseline; hospitalised | Braden; NPUAP, AHCPR, others | Prospective | MEDLINE; EMBASE; CINAHL; KoreaMed;  Cochrane Library; National Digital Science Library; Korea Education and Research Information Service | NS-2013; NS; NS | QUADAS-II | Yes; random-effects model, SROC analysis | 21 (6070) | | 1 |
| Pei^99^ (2023)  Model development & validation  Accuracy | Adult; hospital | ML (if >1 model per study, only the *‘best’* was included); NS | NS | PubMed; Embase; Cochrane Library; Web of Science; CINAHL; Grey literature; and “other databases” | Database inception - 2022; English and Chinese; peer-reviewed articles or full-length conference proceedings | PROBAST | Yes; random-effects model, SROC analysis | 18 (408504) | | 18 |
| Qu^100^ (2022)  Accuracy | Adults with no PIs at baseline; hospital inpatients | ML; Munoz and Posthauer (2021) PI stage or as defined by the study authors | Diagnostic trials, crossover trials, cluster-controlled trials | MEDLINE; EMBASE; EBSCO; Web of Science | Start date between 1985-2010, until 2021; English; NS | QUADAS-II; PROBAST | Yes; fixed-effects or random-effects model dependent on heterogeneity assessment, ANOVA model for Bayesian network meta-analysis for diagnostic test accuracy | 24 (221541) | | 24 |
| Ribeiro^101^ (2021)  Model development | Bedridden patients (at risk of PI); NS | ML; NS | NS | Scopus; Web of Science | 2010-2021; English; review papers, opinion paper, extended abstracts excluded | None | No | 3 (6674) | | 3 |
| Shi^102^ (2019)  Model development & validation | Any; NS | Empirically derived multivariable models, including ML models; NS | Objectives 1 & 2: prospective or retrospective longitudinal  Objective 3: RCTs, non-randomised trials, prospective or retrospective 'before-and-after' | MEDLINE; CINAHL; ProQuest | 1946-2017 (MEDLINE), 1937-2017 (CINAHL), inception-2017 (ProQuest); no restrictions; no restrictions | PROBAST | Yes; fixed-effects or random-effects model dependent on heterogeneity assessment | 23 (72326) | | 21 |
| Tayyib^103^ (2013)  Accuracy  Effectiveness | Adults; ICU | NS; NPUAP/EPUAP | Quantitative | MEDLINE; PubMed; CINHAL; EBSCOHost; Cochrane Library; ProQuest; Google Scholar | 2000-2012; English; journals, books, handbooks, abstracts | None | No | 11 (2119) | | 9 |
| Wang^104^ (2022)  Accuracy | Any age; any healthcare setting | NS; NS | Primary research and sample size, except case reports or case series | PubMed; EMBASE; CINAHL; Cochrane Library | Inception-2021; English; NS | JBI tools; NOS | Yes; fixed-effects or random-effects model dependent on heterogeneity assessment | 2 (992) | | 2 |
| Wei^105^ (2020)  Accuracy | Adults (age >18y); ICU | Braden; NS | NS | PubMed; Web of Science; Cochrane Library; SinoMed; CNKI; Wanfang | NS-2019; no restrictions; NS | QUADAS-II | Yes; DerSimonian and Laird random-efects model | 11 (10044) | | 1 |
| Wilchesky^106^ (2015)  Accuracy | NS; long-term care | Braden; NS | NS | MEDLINE; PubMed; EMBASE; PsychINFO | 1985-2013; English; journal articles (reviews and opinion papers excluded) | None | Yes; DerSimonian and Laird random-effects model | 9 (40361) | | 1 |
| Zhang^107^ (2021)  Accuracy | Inpatient aged >18y; ICU (stay >24h) | PI risk assessment tools; standard for judging the occurrence of PI had to be described | Cohort, case-control | PubMed/MEDLINE; EMBASE; CINAHL; Web of Science; Cochrane Library; China Biomedical Literature Service System; VIP Database; CNKI | Inception-2019; no restrictions; NS | QUADAS-II | Yes; hierarchal SROC model | 23 (15199) | | 15 |
| Zhou^108^ (2022)  Model development | Any inpatient; hospital | ML; NS | NS | PubMed; EMBASE; CINHAL; Web of Science; Scopus | 2010-2021; English; NS | PROBAST | No | 22 (234105) | | 22 |
| Zimmerman^109^ (2018)  Accuracy | Adult inpatients; ICU | Any scale or index; NS | NS | MEDLINE; CINAHL COCHRANE; El Banco de Datos de Enfermería; nursing database; LILACS | 1962-2016; English, Portuguese, Spanish; NS | None | No | 13 | | 11 |

AHRQ – Agency for Healthcare Research and Quality; AHCPR – Agency for Health Care Policy and Research; CASP – Critical Appraisal Skills Programme; CNKI – China National Knowledge Infrastructure; CUIDEN – Bibliographic Database Index Foundation including scientific production on Health Care in Latin American; DARE – Database of Abstracts of Reviews of Effects; DEV – model development study; EPUAP – European Pressure Ulcer Advisory Panel; HCW – health care worker; ICU – intensive care unit; ICD-9 – International Classification of Diseases Ninth Edition; IEEE – Institute of Electrical and Electronics Engineers; JBI – Joanna Briggs Institute; LILACS – Latin America and Caribbean Health Sciences Literature; ML – machine learning; NOS – Newcastle Ottowa Scale; NPUAP – National Pressure Ulcer Advisory Panel; NS – not stated; PI – pressure injury; PROBAST – Prediction model Risk of Bias Assessment; QUADAS – Quality Assessment of Diagnostic Accuracy Studies; SROC – summary receiver operating curve; ULAKBIM – Turkish Academic Network and Information Center; VAL – model validation study.

^a^Patients and HCWs

^b^KQ1 – key question 1 looks at effectiveness of risk assessment tools; KQ2 – key question 2 looks at diagnostic accuracy/validity of risk assessment tools

^b^Version of modified Norton scale cannot be determine.

#### Table S3. AMSTAR-2 assessment results per review

| **Review author**  (pub. year) | **ITEM 1** | **ITEM 2** | **ITEM 3** | **ITEM 4** | **ITEM 5** | **ITEM 6** | **ITEM 7** | **ITEM 8** | **ITEM 9** | **ITEM 10** | **ITEM 11** | **ITEM 12** | **ITEM 13** | **ITEM 14** | **ITEM 15** | **Overall confidence** |
| --- | --- | --- | --- | --- | --- | --- | --- | --- | --- | --- | --- | --- | --- | --- | --- | --- |
| **Model development and validation reviews** | | | | | | | | | | | | | | | | |
| Barghouthi^78^  (2023) | N | N | N | N | Y | N | N | N | N | N | N/A | N/A | N | N | Y | **Critically Low** Y=2/13  PY=0/13  N=11/13 |
| Dweekat^83^ (2023) | N | N | N | PY | Y | N | N | N | N | N | N/A | N/A | N | N | Y | **Critically Low***  Y=2/13  PY=1/13  N=10/13 |
| Jiang^89^  (2021) | N | N | N | PY | Y | N | N | N | PY | N | N/A | N/A | N | N | Y | **Critically Low** Y=2/13  PY=2/13  N=9/13 |
| Pei^99^  (2023) | N | Y | N | Y | N | Y | N | N | Y | N | N | N | Y | Y | Y | **Critically Low**  Y=7/15  PY=0/15  N=8/15 |
| Ribeiro^101^ (2021) | N | N | N | PY | N | Y | N | N | N | N | N/A | N/A | N | N | Y | **Critically Low** Y=2/13  PY=1/13  N=10/13 |
| Shi^102^  (2019) | Y | Y | Y | PY | Y | N | N | Y | Y | N | N | Y | Y | Y | Y | **Low**  Y=10/15  PY=1/15  N=4/15 |
| Zhou^108^  (2022) | N | N | N | PY | Y | Y | N | N | Y | N | N/A | N/A | N | N | Y | **Critically Low** Y=4/13  PY=2/13  N=8/13 |
| **Summary** | **1/7 Yes** | **2/7 Yes** | **1/7 Yes** | **1/7 Yes**  **6/7 PY** | **5/7 Yes** | **3/7 Yes** | **0/7 Yes** | **1/7 Yes** | **3/7 Yes**  **1/7 PY** | **0/7 Yes** | **0/2 Yes** | **1/2 Yes** | **2/7 Yes** | **2/7 Yes** | **7/7 Yes** |  |
| **Prognostic accuracy reviews** | | | | | | | | | | | | | | | | |
| Chen^80^  (2023) | N | N | N | PY | Y | Y | N | PY | Y | N | N | N | N | N | Y | **Critically Low**  Y=3/15  PY=2/15  N=10/15 |
| Chen^81^  (2016) | N | N | N | PY | Y | N | N | PY | PY | N | N | N | N | Y | Y | **Critically Low**  Y=3/15  PY=3/15  N=9/15 |
| Chou^82^ (2013) | Y | Y | N | Y | Y | N | Y | Y | PY | Y | N | Y | Y | N | Y | **Low**  Y=10/15  PY=1/15  N=4/15 |
| Garcia-Fernandez^84^ (2014) | N | N | Y | PY | N | N | N | N | N | N | Y | Y | Y | N | N | **Critically Low**  Y=4/15  PY=1/15  N=10/15 |
| He^86^  (2012) | N | N | N | N | Y | N | N | N | Y | N | N | N | N | Y | Y | **Critically Low**  Y=4/15  PY=0/15  N=11/15 |
| Huang^88^ (2021) | N | Y | N | PY | Y | Y | N | N | Y | N | Y | N | N | Y | Y | **Critically Low** Y=7/15  PY=1/15  N=7/15 |
| Mehicic^93^  (2024) | N | Y | N | N | Y | Y | N | N | N | N | NA | NA | N | Y | Y | **Critically Low**  Y=5/13  PY=0/13  N=8/13 |
| Pancorbo-Hidalgo^110^ (2006) | N | N | Y | PY | N | Y | N | PY | N | N | N | Y | Y | N | N | **Critically Low**  Y=4/15  PY=2/15  N=9/15 |
| Park^96^ (2016a) | N | N | N | PY | N | N | N | PY | N | N | N | N | N | Y | Y | **Critically Low**  Y=2/15  PY=2/15  N=11/15 |
| Park^97^ (2016b) | N | N | N | PY | N | Y | N | PY | N | N | N | N | N | Y | Y | **Critically Low**  Y=3/15  PY=2/15  N=10/15 |
| Park^98^  (2015) | N | N | N | PY | Y | Y | N | PY | N | N | N | Y | Y | Y | Y | **Critically Low**  Y=6/15  PY=2/15  N=7/15 |
| Pei^99^  (2023) | N | Y | N | Y | N | Y | N | N | Y | N | N | N | Y | N | Y | **Critically Low**  Y=6/15  PY=0/15  N=9/15 |
| Qu^100^  (2022) | N | Y | N | N | Y | Y | N | N | Y | N | N | N | Y | N | Y | **Critically Low**  Y=6/15  PY=0/15  N=9/15 |
| Tayyib^103^ (2013) | N | N | N | PY | N | N | N | N | N | N | NA | NA | N | N | N | **Critically Low**  Y=0/13  PY=1/13  N=12/13 |
| Wang^104^ (2022) | N | N | N | PY | Y | Y | N | N | N | N | Y | N | Y | N | Y | **Critically Low**  Y=5/15  PY=1/15  N=9/15 |
| Wei^105^  (2020) | N | N | N | PY | Y | Y | N | N | PY | N | N | N | Y | Y | N | **Critically Low**  Y=4/15  PY=2/15  N=9/15 |
| Wilchesky^106^ (2015) | N | N | N | PY | N | N | N | N | N | N | N | N | N | Y | Y | **Critically Low**  Y=2/15  PY=1/15  N=12/15 |
| Zhang^107^ (2021) | N | Y | N | PY | Y | Y | N | PY | Y | N | Y | N | Y | Y | Y | **Low**  Y=8/15  PY=2/15  N=5/15 |
| Zimmerman ^109^ (2018) | N | N | N | N | Y | N | N | N | N | N | NA | NA | N | N | N | **Critically Low** Y=1/13  PY=0/13  N=12/13 |
| **Summary** | **1/19 Yes** | **6/19 Yes** | **2/19 Yes** | **2/19 Yes**  **13/19 PY** | **12/19 Yes** | **11/19 Yes** | **1/19 Yes** | **1/19 Yes**  **7/19 PY** | **6/19 Yes**  **3/19 PY** | **1/19 Yes** | **4/16 Yes** | **4/16 Yes** | **9/19 Yes** | **10/19 Yes** | **14/19 Yes** |  |
| **Clinical effectiveness reviews** | | | | | | | | | | | | | | | | |
| Baris^79^  (2015) | N | N | N | PY | N | Y | N | N | N | N | NA | NA | N | N | N | **Critically Low** Y=1/13  PY=1/13  N=11/13 |
| Chou^82^ (2013) | Y | Y | N | Y | Y | N | Y | Y | Y | Y | NA | NA | Y | N | Y | **Moderate**  Y=10/13  PY=0/13  N=3/13 |
| Gaspar^85^ (2019) | Y | N | N | PY | Y | N | N | Y | N | N | NA | NA | N | Y | Y | **Critically Low**  Y=5/13  PY=1/13  N=7/13 |
| Health Quality Ontario^87^ (2009) | N | N | N | N | N | N | N | Y | N | N | NA | NA | Y | N | Y | **Critically Low**  Y=3/13  PY=0/13  N=10/13 |
| Kottner^90^ (2009) | N | N | N | PY | Y | Y | N | Y | PY | N | NA | NA | Y | Y | Y | **Critically Low**  Y=6/13  PY=2/13  N=5/13 |
| Lovegrove^91^ (2021) | Y | PY | N | PY | Y | Y | N | Y | PY | N | NA | NA | Y | Y | Y | **Low**  Y=7/13  PY=3/13  N=2/13 |
| Lovegrove^92^ (2018) | N | Y | N | PY | Y | Y | N | Y | PY | N | NA | NA | Y | Y | Y | **Low**  Y=7/13  PY=2/13  N=4/13 |
| Mehicic^93^  (2024) | Y | Y | N | N | Y | Y | N | N | PY | N | NA | NA | Y | N | Y | **Critically Low**  Y=6/13  PY=1/13  N=6/13 |
| Moore^94^ (2019) | Y | Y | N | Y | Y | N | Y | Y | Y | Y | NA | NA | Y | Y | Y | **High**  Y=11/13  PY=0/13  N=2/13 |
| Pancorbo-Hidalgo^110^ (2006) | N | N | Y | PY | N | Y | N | Y | PY | N | NA | NA | Y | N | N | **Critically Low** Y=4/13  PY=3/13  N=6/13 |
| Tayyib^103^ (2013) | N | N | N | PY | N | N | N | PY | N | N | NA | NA | N | N | N | **Critically Low** Y=0/13  PY=2/13  N=11/13 |
| **Summary** | **4/11 Yes** | **4/11 Yes**  **1/11 PY** | **1/11 Yes** | **2/11 Yes**  **7/11 PY** | **7/11 Yes** | **6/11 Yes** | **2/11 Yes** | **8/11 Yes**  **1/11 PY** | **2/11 Yes**  **5/11 PY** | **2/11 Yes** | **11/11 NA** | **11/11 NA** | **8/11 Yes** | **5/11 Yes** | **8/11 Yes** |  |
| Item 1 – Adequate research question/ inclusion criteria?; Item 2 – Protocol and justifications for deviations?; Item 3 – Reasons for study design inclusions?; Item 4 – Comprehensive search strategy?; Item 5 – Study selection in duplicate?; Item 6 – Data extraction in duplicate?; Item 7 – Excluded studies list (with justifications)?; Item 8 – Included studies description adequate?; Item 9 – Assessment of RoB/quality satisfactory?; Item 10 – Studies’ sources of funding reported?; Item 11 – Appropriate statistical synthesis method?; Item 12 – Assessment of impact of RoB on synthesised results?; Item 13 – Assessment of impact of RoB on review results?; Item 14 – Discussion/investigation of heterogeneity?; Item 15 – Conflicts of interest reported?  * Note that many items were not as applicable here, as the Dweekat 2023^83^ review is a methodological review; N – No; N/A – Not Applicable; PY – Partial Yes; RoB – Risk of Bias; Y – Yes. Further details on AMSTAR items are given in Appendix 4. | | | | | | | | | | | | | | | | |

#### Table S4. Risk prediction tool characteristics, ascertained at review level

| **Name of tool (publication year)** | **Considered in included systematic reviews** | **Country; setting; patients; data source** | **Type of model; model development algorithm** | **Prediction horizon** | **Interval validation method** | **Predictors/domains in final model** | **N patients (n events)** | **Patient characteristics** | **Performance metrics** |
| --- | --- | --- | --- | --- | --- | --- | --- | --- | --- |
| Abruzzese (1985)^111^ | No | NS | Statistical; NS | NS | NS | General health; mental status; activity; mobility; continence; nutrition; oral nutrition intake; oral fluid intake; pre-disposing diseases (vascular disease, neuropathies, diabetes, anemias etc.) | NS | NS | NS |
| Admission PU-FIM model (2014)^112^ | Yes ^102^ | NS; rehabilitation units for SCI; NS; prospective | ML; LR, recursive partitioning analysis | Mean follow-up 36.5 (SD 31.4) days | NS | NS | 159 (21) | Mean (SD) age: 46.9 (19.1) Female: 22.0% | C-statistic: 0.77 (95% CI 0.65–0.86) |
| Andersen (1982)^113^ | Yes ^82 84 110^ | Denmark; acute hospital inpatient; adult; prospective | Statistical; NS | 10 days in-hospital observation; 3-months total observation | NS | NS | 3398 (40) | NS | NS |
| Arnell (1983)^114^ | Yes ^84^ | NS; NS; adult; NS | Statistical; NS | NS | NS | NS | NS | NS | NS |
| Berlowitz – 11-item  (1996)^115^ | Yes ^102^ | USA; long-term care; adult veterans; retrospective | Statistical; LR | 6 months | NS | NS | 31150 (1350) | Mean (SD) age: 70 (11.6) Female: 3% | O/E ratio: 1.0 (95% CI 0.95-1.05)  C-statistic: 0.75 (95% CI 0.74–0.76) |
| Berlowitz MDS risk-adjustment model (2001)^116^ | Yes ^102^ | USA; long-term care; adult nursing home residents; retrospective | Statistical; LR | 3 months | NS | NS | 14607 (905) | Mean (SD) age: 82.5 (10.9) Female: 75.4% | O/E ratio: 0.97 (95% CI 0.91-1.04)  C-statistic: 0.73 (95% CI 0.71–0.75) |
| Braden (1987)^117^ | Yes ^59 79 81 82 84 86 88 91-94 97 98 103-107 109 110^ | USA; hospital; elderly; NS | Statistical; NS | 12 weeks | NS | Sensory perception; moisture; activity; mobility; nutrition; friction and shear | 102 (28) | NS | NS |
| Braden – Baldwin 2-item (1998)^118^ | Yes ^102^ | NS; trauma and burn centres; NS; prospective | Statistical; LR | 26.5 days | NS | NS | 36 (11) | Mean (SD) age: 31.8 (10.9) Female: 27.8% | NS |
| Braden – Bergquist 2-item (2001)^119^ | Yes ^102^ | USA; long-term care; adult ($>$60y); retrospective | Statistical; Cox regression | Mean follow-up 60.6 (SD 94.3) days | NS | NS | 1684 (107) | Mean (SD) age: 76.4 (8.6) Female: 62.4% | NS |
| Braden – Bergquist 3-item (2001)^119^ | Yes ^102^ | USA; long-term care; adult ($>$60y); retrospective | Statistical; Cox regression | Mean 60.6 follow-up (SD 94.3) days | NS | NS | 1684 (107) | Mean (SD) age: 76.4 (8.6) Female: 62.4% | NS |
| Braden modified by Choi & Song (1991)^120 121^ | Yes ^82 84 96 103 107 109^ | Korea; ICU (neurological problems); NS; prospective | Statistical; NS | NS | NS | Body temperature; amount of medication (analgesics, sedation, anticoagulants); sensory perception; activity and mobility; moisture; nutrition; friction and shear | 146 (17) | Female: 39% | NS |
| Braden modified by Halfens/4-factor model (2000)^122^ | Yes ^84 103 107 109^ | Netherlands; hospital inpatients; NS; prospective | Statistical; stepwise LR | NS | NS | Sensory perception; moisture; friction and shear; age | 320 (47) | Mean age: 61 Female: 48%  Ethnicity: white 100% | NS |
| Braden modified by Kwong (2005)^123^ | Yes ^82 84^ | China; acute hospital inpatient; any; prospective | Statistical; NS | Mean of 11 days (range 5-21 days) | NS | NS | 429 (9) | Mean (SD) age: 54 (17) Female: 41% | NS |
| Braden modified by Pang & Wong (1998)^124^ | Yes ^96^ | NS; rehabilitation hospital (medical and orthopaedic); NS; NS | Statistical; NS | 2 weeks | NS | NS | 138 (NS, 20%) | NS | NS |
| Braden modified by Schue (1998)^125^ | Yes ^102^ | NS; rehabilitation units; adult males; retrospective | Statistical; LR | NS | NS | NS | 170 (9) | Mean (SD) age: 69.2 (10.9) All male | NS |
| COMHON (2011)^126^ | Yes ^107^ | Spain; ICU ($>$72h); NS; NS | Statistical; NS | NS | NS | NS | NS | NS | NS |
| Compton ICU model (2008)^127^ | Yes ^84 102^ | Germany; ICU; adult; retrospective | Statistical; LR | Median follow-up 6 (IQR 3-14) days | NS | NS | 698 (121) | Median (IQR) age: 66 (56-75.3) Female: 43.8% | C-statistic: 0.82 (95% CI 0.78–0.85) |
| Cubbin & Jackson (1991)^128^ | Yes ^80 84 96 107 109 110^ | UK; ICU; adult; NS | Statistical; NS | NS | NS | Age; weight; general skin condition; mental status; mobility; hemodynamics; nutrition; respiration; incontinence; hygiene | NS | NS | NS |
| Cubbin & Jackson revised (Jackson & Cubbin) (1999)^129^ | Yes ^82 84 103^ | UK; ICU; adult; NS | Statistical; NS | NS | NS | Age; weight; past medical history; general skin condition; mental condition; mobility; haemodynamics; respiration; oxygen requirement; nutrition; incontinence; hygiene | NS | NS | NS |
| Delmore scale (2015)^130^ | No | USA; hospital; any ($\geq$8y); NS | Statistical; NS | NS | NS | NS | NS | NS | NS |
| Douglas (1986)^131^ | Yes ^82 84 103 107 109 110^ | NS; ICU; NS; NS | Statistical; NS | NS | NS | Pain; activity; physical condition; incontinence; steroid therapy; diabetes; cytotoxic therapy; dyspnea | NS | NS | NS |
| DUPA (1995)^132^ | Yes ^84^ | USA; ICU; adult; NS | Statistical; NS | NS | NS | NS | 85 | NS | NS |
| Dutch CBO Score (1992)^133^ | Yes ^82^ | Netherlands; hospital; NS; retrospective | Statistical; NS | NS | NS | NS | NS | NS | NS |
| EMINA (2001)^134^ | Yes ^84 107 110^ | Spain; ICU (long-stay); adult; NS | Statistical; NS | 7 days | NS | NS | 673 (47) | NS | NS |
| EVARUCI scale (2001)^135^ | Yes ^84 107 109^ | Spain; ICU; NS; NS | Statistical; NS | NS | NS | NS | NS | NS | NS |
| Extended Braden (2000)^122^ | Yes ^82 103 107^ | Netherlands; hospital inpatient (medical and surgical); NS; prospective | Statistical; stepwise LR | NS | NS | NS | 320 (47) | Mean age years: 60.9 Female: 48% Ethnicity: white 100% | NS |
| Finnish risk assessment scale (2006)^14^ | No | Finland; long-term care; NS; prospective | Statistical; expert consensus and investigating agreement percentages | NS | NS | Urinary incontinence; activity; mental status; nutrition; mobility; sensory perception; skin condition; appetite; devices section; care methods section | Phase 1^a^: 43 raters, 6 patients Phase 2^b^: 64 (50 analysed) experts | NS | NS |
| Fragmment scale (2002)^136^ | Yes ^82 84 110^ ^102^ | Switzerland; hospital inpatient (acute, medical surgical); adult ($>$16y); prospective | Statistical; LR, Cox regression | Follow-up duration of 3 weeks, with mean follow-up 9 days | CV | NS | 1190 (182) | Mean (range) age: 61 (16-96) Female: 45.4% | C-statistic: 0.80 (95% CI 0.77–0.84) |
| Gosnell (1973)^137^ | Yes ^82 84 107^ | USA; ICU; NS; NS | Statistical; NS | NS | NS | Mental status; continence; movement control; ability to ambulate; process of food intake (evaluation includes recording of vital signs, skin condition and medications, but these are not scored) | NS | NS | NS |
| Hatanaka (2008)^138^ | Yes ^82 102^ | Japan; acute hospital inpatient; elderly; prospective | Statistical; LR, Cox regression | Mean follow-up 33 (range 5-79) days | NS | Haemoglobin; CRP; albumin; age; gender | 149 (38) | Mean (SD) age: 72 (11) Female: 30% | C-statistic: 0.79 (95% CI 0.66–0.88) |
| HPUR (2003)^139^ | Yes ^84^ | Sweden; palliative care; adult; NS | Statistical; NS | NS | NS | NS | 54 | NS | NS |
| Knoll Decubitus Ulcer Potential Scale (1988)^140^ | Yes ^82 84 110^ | USA; long-term care; adult ($>$65y); prospective | Statistical; NS | 28 days | NS | General health; mental health; activity; mobility; incontinence; oral nutrition intake; oral fluid intake; predisposing diseases | 60 (28) | Mean (range) age: 81 (65-97) Female: 80% Ethnicity: white 72%; black 15%; Asian 2%; unknown 11% | NS |
| Maelor score (2000)^141^ | Yes ^91 92^ | UK; NS; NS; NS | Statistical; NS | NS | NS | NS | NS | NS | NS |
| Mainland China | No | NS | NS; NS | NS | NS | NS | NS | NS | NS |
| Medley score (1987) | No | UK; acute medical ward, long-term care; NS; NS | Statistical; NS | NS | NS | NS | NS | NS | NS |
| ML Ahmad (2021)^142^ | Yes ^83^ | USA; inpatient, out-patient, nursing home; NS; prospective | ML; LR, other | NS | 10-fold CV | NS | 713 (NS, 52.3%) | NS | NS |
| ML Alderden [1] (2018)^143^ | Yes ^78 83 89 99 101 108^ | USA; surgical ICU, surgical cardiovascular ICU; adult; retrospective | ML; RF | NS | Split sample; 67% training 33% testing | Hypotension; Glasgow Coma Scale; oxygenation; BMI at admission; laboratory value (albumin, creatinine, glucose, haemoglobin, lactate, prealbumin); surgical time; age | 6376 (1. 516^c^; 2. 257^d^) | Mean (SD) age: 54 (19) Female: 37.7% Ethnicity: white 100% | Accuracy^e^: 0.790  AUC (SD):  1. 0.79  2. 0.79 |
| ML Alderden [2] (2021)^144^ | Yes ^83 108^ ^78^ | USA; surgical ICU; adult (≥18y); retrospective | ML; RF, MLP (ANN), other (AdaBoost, Gradient Boosting, LR) | NS | Split sample; 80% training 20% testing | Minimum albumin; minimum arterial PaO_2_; surgery duration; vasopressin infusion; length of ICU stay prior to HAPI; skin assessment; Braden scale scores | 5101 (NS, 6.5%) | NS | F1 score: 0.34  AUC (SD): 0.8 (0.02) |
| ML Anderson (2021)^145^ | Yes ^83^ ^78 99^ | USA; surgical ICU; adult (≥18y); retrospective | ML; LR, RF, NN, DL | NS | Split sample; 70% testing vs. 30% testing | NS | 23000 (738) | NS | Accuracy^e^: 0.86-0.99  Sensitivity: 0.67-1  Specificity: 0.91-0.99  PPV: 0.82-0.98  NPV: 0.88-1  AUC: 0.71-0.72 |
| ML Borlawsky (2007)^146^ | Yes ^83 102^ | USA; acute hospital inpatient; NS; retrospective | ML; DT | NS | 4-fold CV; tree pruning of the DT tool | NS | 3300 (206) | NS | NS |
| ML Cai (2021)^147^ | Yes ^83 100 101 108^ ^78^ | China; cardiovascular surgery; any; prospective | ML; DT, XGBoost | NS | NS | Age; gender; disease category; weight; duration of surgery; duration of cardiopulmonary bypass procedure | 149 (37) | NS | AUC: 0.81  Sensitivity: 0.08  Specificity: 1  PPV: 1  NPV: 0.77 |
| ML Charon (2022)^148^ | Yes ^83^ | France; nursing home; NS; NS | ML; RF, BN | NS | NS | NS | 3000 | NS | NS |
| ML Chen [1] (2018)^149^ | Yes ^83 89 100 101 108^ | China; cardiovascular surgery; NS; retrospective | ML; ANN | NS | Split sample; 70% training 30% testing | Length of surgery; disease category; age; perioperative corticosteroid administration | 149 (37) | Mean (SD) age: 49.8 (17.7)  Female 46.9% | F1 score: 0.19  Accuracy^e^: 0.815  Accuracy^e^: 0.28  Sensitivity: 0.11  Specificity: 0.79  PPV: 0.67  NPV: 0.21 |
| ML Chen [2] (2019)^150^ | Yes ^100^ | NS; CVD patients; adult; NS; NS | ML; LR | NS | NS | Preoperative haemoglobin value; blood sodium value; preoperative albumin; intraoperative mean body temperature; lowest mean arterial pressure; serum potassium value; smoking frequency; history of hypertension; age | 1163 (67) | NS | NS |
| ML Chen YC (2008)^151^ | Yes ^83^ | Taiwan; post-surgery; NS; retrospective | ML; LR, DT, SVM, other | NS | 3-fold CV | NS | 168 (NS, 4.8%) | NS | NS |
| ML Cheng [1] (2020)^152^ | Yes ^78^ | NS; hospital; NS; NS | ML; LR | NS | Split sample; 83% training 17% testing | Age; movement; sensory perception; response; moisture; perfusion; use of medical devices; compulsive position; hypoalbuminemia; HAPI; surgery | 2341 | NS | AUC: 0.87-0.94 |
| ML Cheng [2] (2021)^153^ | Yes ^83^ | China; hospital inpatient; elderly (>65y); retrospective | ML; RF, SVM, MLP, other | NS | 5-fold CV | NS | 245 (NS, 80%) | NS | NS |
| ML Cho (2011)^154^ | Yes ^83 100^ | Korea; ICU; adult (≥18y); NS | ML; LR, NN | NS | NS | Gender; BMI; transfer from ER; medical diagnosis; systolic blood pressure; diastolic blood pressure; number of urinations; number of defecations; any stompy; number of drain tubes; any catheterization; number of position changes; peripheral sensory; skin condition; albumin; number of sedatives administered; any strain; age; length of stay; any inotropic support; total parenteral nutrition; any sedation; number of analgesics administered | 21,114 for NN model; 21,069 for LR model (3348) | NS | NS |
| ML Choi (2020)^155^ | Yes ^108^ ^78^ | Korea; intubated ICU; adult (≥18y); prospective | ML; LR, BN (Gaussian NB) | NS | CV; split sample; 80% training 20% testing | Oral mucosal; bite-block or airway use; endotracheal tube; holder use; steroid use; vasopressor use; haematocrit; albumin | 27 (NS, 55.6%) | NS | AUC: 0.68-0.82  Sensitivity: 0.60-0.85  Specificity: 0.76-0.89  PPV: 0.23-0.37  NPV: 0.97-0.98 |
| ML Cichosz (2019)^156^ | Yes ^83 108^ | Denmark; any hospital (ICU, medical, surgical); adult (>20y); retrospective | ML; LR | NS | 10-fold CV; split sample; 65% training 35% testing | Gender; up and self-reliant; limitation in activity performance; mobility and willingness; consciousness | 383 (NS, 28.1% in training cohort, 18% in testing cohort) | NS | AUC: 0.82  Sensitivity: 0.43  Specificity: 0.94  PPV: 0.72  NPV: 0.92 |
| ML Cramer (2019)^157^ | Yes ^78 83 99 100 108^ | USA; ICU; adult (≥18y); retrospective | ML; LR, RF, SVM, MLP (ANN), other (GB, EN, NN) | NS | 5-fold CV; split sample 80% training 20% testing | Stage 1 PI within the first 24h; Glasgow Coma Scale; blood urea nitrogen; PaO_2;_ cardiac surgery recovery unit; albumin; medical ICU; pressure reduction device; mechanical ventilation; mean arterial pressure (top 10 predictors in two highest performing models) | 50851 (1690) | NS | Sensitivity: 0.49  PPV: 0.09  NPV: 0.71 |
| ML Delparte (2021)^158^ | Yes ^108^ | Canada; SCI rehabilitation centre; NS; retrospective | ML; LR, DT | NS | NS | PI history; ambulation; FIM toileting scores; FIM bed transfer scores | 807 (NS, 22%) | Mean age: 54 | AUC: 0.78  Sensitivity: 0.93  Specificity: 0.63  PPV: 0.4  NPV: 0.97 |
| ML Deng [1] (2016)^159^ | Yes ^89 99 100^ | China; ICU; adult; retrospective | ML; DT | NS | NS | Age; faecal incontinence; Braden total score; diastolic blood pressure | 468 (94) | Mean (SD) age: 57.81 (16.72) Female: 25.4% | AUC: 0.83 (95% 0.78-0.88)  Sensitivity: 0.81  Specificity: 0.70 |
| ML Deng [2] (2017)^160^ | Yes ^83 100 108^ | China; ICU, medical ICU; adult; retrospective | ML; LR, DT | NS | 10-fold CV | Age; ICU length of stay; diastolic blood pressure; albumin level; mechanical ventilation; total Braden score; faecal incontinence | 468 (94) | Mean (SD) age: 58 (17) Female: 25.4% | AUC: 0.93 (95% CI 0.88-0.97)  Sensitivity: 0.86  Specificity: 0.82  PPV: 0.76  NPV: 0.98 |
| ML Deschepper (2022) ^161^ | Yes ^78^ | NS; ICU; NS; prospective | ML; RF | NS | Split sample; 90% training 10% testing | Age; gender; diagnosis; Braden score; BMI; heart rate; mean arterial pressure; temperature; laboratory results; immunocompromised status | 13254 | NS | Accuracy^e^: 0.830  AUC: 0.785-0.792 |
| ML Do (2022)^162^ | Yes ^78 83 99^ | USA; hospital; adult; retrospective | ML; LR, RF, DT, KNN, NB | NS | 5-fold CV; split sample; 70% training 30% testing | Bed positions; laboratory tests (creatinine, lactate, pre-albumin, and albumin); clinical features; admission weight; BMI; activity; Braden assessment | 6742 (NS, 31.9%) | Average age: 61.5 Female: 47.3% | Accuracy^e^: 0.90-0.97  Sensitivity: 0.86-0.98  Specificity: 0.87-0.97  PPV: 0.81-0.96  NPV: 0.92-0.99  AUC: 0.90-0.97 |
| ML Eshetie (2023)^163^ | Yes ^78^ | NS; residential aged care; elderly; retrospective | ML; Fine-Gray Model | NS | Split sample; 80% training 20% testing | History of PIs; lower care needs mainly mobility; toileting; complex health care; medication assistance | 206540 | NS | AUC: 0.72-0.75 |
| ML Gao (2018)^164^ | Yes ^83 100^ | China; surgical; any; retrospective | ML; LR, DT | NS | NS | Application of external force during operation; lean body mass; time of operation $\geq$6h; prone position operation; cardiopulmonary bypass during operation; intraoperative blood loss | 1963 (48) | NS | NS |
| ML Goodwin (2020)^165^ | Yes ^108^ | USA; ICU; adult (≥16y); retrospective | ML; LR, SVM, CANTRIP, LSTM | NS | NS | NS | 35218 (NS, 39.8%) | NS | F1 score: 0.53  Accuracy^e^: 0.84  AUC: 0.87  Sensitivity: 0.72  Specificity: 0.85  PPV: 0.42 |
| ML Hou (2010)^166^ | Yes ^100^ | NS; hospital; NS; NS | ML; LR | NS | NS | Wet skin conditions; activity conditions; movement conditions; nutrition; mental awareness; bowel control; blood sugar; age; haemoglobin | 303 (46) | NS | NS |
| ML Hu (2020)^167^ | Yes ^78 83 99 100 108^ | China; hospital; adult; retrospective | ML; LR, DT, RF | NS | 10-fold CV; split sample; 50% training 50% testing | Skin integrity; systolic pressure; expression ability; capillary refill time; level of consciousness; eye-opening; level of mobility; emotional responses; diastolic pressure; skin properties; colour in the peripheral limbs; pulse rate | 11838 (161) | NS | F1 score: 0.08  AUC: 0.88-1  Sensitivity: 0.69-1  Specificity: 0.72-0.99  PPV: 0.79-0.99  NPV: 0.82-1 |
| ML Hyun (2019)^168^ | Yes ^83 100^ ^78^ | USA; ICU; adult; retrospective | ML; LR | NS | Split sample; 67% training 33% testing | Age; gender; weight; diabetes; vasopressor; isolation; endotracheal tube; ventilator episode; Braden score; ventilator days | 12654 (753) | NS | Accuracy^e^: 0.81  Sensitivity: 0.65  Specificity: 0.69  PPV: 0.211  NPV: 0.956  AUC: 0.737 |
| ML interRAI PURS (2010)^169^ | Yes ^102^ | Canada; long-term care; elderly; retrospective | ML; LR, DT | Follow-up 91 days | NS | NS | 14083 (503) | Mean (SD) age: 82.2 (10.2) Female: 69.2% | C-statistic: 0.71 (95% CI 0.68–0.73) |
| ML James (2021)^170^ | Yes ^78^ | USA; hospital; NS; retrospective | ML; EBM, DT, LR | NS | NS | Patient history (age and gender); vital signs (heart rate and blood pressure); lab tests (hemoglobin and creatinine levels); length of stay; procedures; medications | 100355 | NS | AUC: 0.60-0.79 |
| ML Jin (2017)^171^ | Yes ^83^ | Korea; hospital; adult; retrospective | ML; LR | NS | Split sample; 80% training 20% testing | NS | 11191 (NS, 20%) | NS | NS |
| ML Kaewprag [1] (2015)^172^ | Yes ^108^ | USA; ICU; adult (≥18y); retrospective | ML; LR, DT, SVM, NB, RF, KNN | NS | 10-fold CV | Braden subscale; medication (18 medication categories); diagnosis (61 comorbid conditions) | 7717 (NS, 7.6%) | NS | g-means: 0.618  AUC: 0.83  Sensitivity: 0.16  Specificity: 0.99  PPV: 0.56  NPV: 0.93 |
| ML Kaewprag [2] (2017)^173^ | Yes ^83 89 99 100 108^ | USA; ICU; adult; retrospective | ML; BN (chosen method), LR, DT, NN, RF, SVM | NS | Split sample; 67% training 33% testing | Braden subscale; medication (18 medication categories); diagnosis (61 comorbid conditions) | 7717 (590) | Mean (SD) age: 57.7 (15.9) Female: 42.6% | Accuracy^e^: 0.819  AUC (SD): 0.83 (0.01)  Sensitivity: 0.478  Specificity: 0.895  Sensitivity (SD): 0.46 (0.03)  Specificity: 0.91 (0.01)  PPV (SD): 0.29 (0.18)  NPV: 0.95 |
| ML Kim [1] (2006)^174^ | Yes ^100^ | USA; hospital (>4d stay); adult (≥18y); NS | ML; DT | NS | NS | Age; race; gender; visit type; diagnoses; procedures; tests/results; drugs | 826 (52) | NS | NS |
| ML Kim [2] (2006)^174^ | Yes ^100^ | USA; hospital (>4d stay); adult (≥18y); NS | ML; LOS | NS | NS | Presence of edema (cardiovascular system); presence of an indwelling foley catheter; presence of nutrition consult triggered for potential or actual nutritional imbalance; use of a wheelchair as an assistive device during the hospital stay; presence of a need for extra nursing care | 2347 (84) | NS | NS |
| ML Ladios-Martin (2020)^175^ | Yes ^78 83 99 100 108^ | Spain; ICU; adult (≥16y); retrospective | ML; LR, DT, RF, SVM, MLP (ANN), BN, other | NS | Split sample; 64% testing 36% testing | Medical service; days of oral antidiabetic agent or insulin therapy; ability to eat; number of red blood cell units transfused; haemoglobin range; PI present on admission; illness severity (total APACHE II score); gender; age; place of birth | 6694 (208) | Mostly aged 65-84 (61.0%)  Female: 34.2% | Accuracy^e^: 0.87  AUC: 0.88  Sensitivity: 0.75  Specificity: 0.88  PPV: 0.22  NPV: 0.99 |
| ML Lee (2021)^176^ | Yes ^83^ | Korea; nursing home; NS; NS | ML; RF, LR, SVM | NS | NS | NS | 60 | NS | NS |
| ML Levy (2022)^177^ | Yes ^83 99^ | USA; hospital inpatient (≥2d stay); adult (≥18y); retrospective | ML; LR, DT, RF, NB, XGBoost | NS | 5-fold CV; 80% training 20% testing | NS | 57227 (241) | Mean (SD) age: 65.5 (15.0) for PI present,  60.1 (18.4) for PI absent  Female: 48.2% | AUC: 0.91 |
| ML Li [1] (2019)^178^ | Yes ^83 89 99 100 108^ | China; hospice/end of life; adult; retrospective | ML; LR, DT, SVM, MLP, ANN | NS | K-fold CV (K not specified) | History of PIs, without cancer, excretion, activity/mobility, and skin condition/circulation | 2062 (1026) | Mean (SD) age: 75.5 (14.4) Female: 45.4% | g-means: 0.770 (DT); 0.779 (NN); 0.798 (SVM)  Accuracy^e^: 0.772 (DT); 0.781 (NN); 0.793 (SVM)  Sensitivity: 0.796 (DT); 0.814 (NN); 0.810 (SVM)  Specificity: 0.748 (DT); 0.749 (NN); 0.788 (SVM) |
| ML Li [2] (2020)^179^ | Yes ^100^ | NS; hospital; NS; NS | ML; SVM, NN | NS | NS | Department category; BMI; skin type; incontinence; poor eating/lack of appetite; feeling restricted; total assessment score; activity score | 554 (345) | NS | NS |
| ML Nakagami (2021)^180^ | Yes ^78 83 99 100 108^ | Japan; any inpatient; adult (≥20y); retrospective | ML; LR, RF, SVM, DT, XGBoost | NS | 5-fold CV; split sample; 70% training 30% testing | Paralysis (difficulty in moving around, difficulty in going up and down stairs, difficulty in transfer, difficulty in standing up, difficulty in keeping standing position); anorexia; age; gender; diagnoses/comorbidities; diet; pain; level of consciousness; skin condition; severity of illness; department type | 75353 (395) | Mean (SD) age: 66.5 (15.9) for PI present,  58.0 (18.5) for PI absent  Female: 47.2% | AUC (SD): 0.80 (0.02)  Sensitivity (SD): 0.78 (0.03)  Specificity (SD): 0.74 (0.04)  PPV: 0.01-0.02  NPV: 0.99 |
| ML Ossai (2021)^181^ | Yes ^83^ | Australia; acute care; any; retrospective | ML; DT, RF, MLP, KNN, LDA | NS | 10-fold CV | NS | 1014 | NS | NS |
| ML Park (2019)^182^ | Yes ^100^ | Korea; acute surgical care; NS; NS | ML; LR | NS | NS | Need for assistance with hygiene; decreased consciousness; Foley catheter; cardiac stimulant; oxygen therapy; low serum albumin; sensory perception impairment; impaired skin integrity; decreased mobility; surgery; nutrition consultation | 400 (80) | NS | NS |
| ML Setoguchi (2016)^183^ | Yes ^83 89 100 108^ | Japan; any hospital; any; retrospective | ML; DT | NS | 10-fold CV | Transfer activity; operation time; BMI | 8286 (NS, 0.62%) | Female: 44.4% | Accuracy^e^: 0.721  Sensitivity: 0.793  Specificity: 0.721 |
| ML Shui (2021)^184^ | Yes ^78^ | NS; hospital, ICU; adult; retrospective | ML; Fine-Gray Model | NS | Split sample; 70% training 30% testing | Age; BMI; lactate serum; Braden score; vasopressor use; antifungal medications | 18019 | NS | AUC: 0.56-0.92 |
| ML Šín (2022)^185^ | Yes ^78 83 99^ | USA; hospital, ICU; NS; retrospective | ML; LR, RF, SVM, MLP, KNN, BN, NB | NS | Split sample; 80% training 20% testing | Age; gender; ethnicity; total intake; total output; length of hospital stays; arterial oxygen saturation; systolic arterial blood pressure; height; daily weight; glucose (whole blood) | 9304 (4652) | NS | Accuracy^e^: 0.84-0.96  PPV: 0.75-0.94  NPV: 0.59-0.91  AUC: 0.77-0.94  F1 score: 0.93 |
| ML Song [1] (2021)^186^ | Yes ^83 99 100 108^ | China; hospital; adult (≥18y); retrospective | ML; DT, RF, SVM, MLP (ANN), KNN, LDA | NS | 10-fold CV; split sample; 50% training 50% testing | Gender; age; height; weight; total intake (mL); total output (mL); body temperature; systolic blood pressure (mmHg); blood glucose; length of stay (days); whether to stay in bed; whether to use restraint bands; surgery; diarrhoea; diabetes; fracture; Norton PI assessment; nutritional assessment; acceptance of passive turning over | 5814 (1673) | Mean (SD) age: 64.3 (18.3) for PI present,  51.9 (17.6) for PI absent  Female: 40.7% | F1 score: 0.83-1  Accuracy^e^: 0.79-1  AUC: 0.95-1  AUPRC: 0.96  Sensitivity: 0.99  Specificity: 0.99  PPV: 0.91-1  NPV: 0.87-1 |
| ML Song [2] (2021)^187^ | Yes ^78 83 99 100 108^ | USA; ICU or ≥24h in acute care; adult; retrospective | ML; LR, RF, SVM, MLP (ANN) | NS | 5-fold CV; split sample; 80% training 20% testing | PI; race; gender; age; Glasgow Coma Scale; level of consciousness; gait/ transferring; activity; pain score; diabetes; peripheral vascular disease; spinal cord injury; stroke; anaemia; albumin; blood urea nitrogen; chloride; potassium; sodium; creatinine; haemoglobin; white blood cell count; platelet blood count | 188512 (6165) | Mean (SD) age: 69.1 (15.5) for cases,  70.3 (7.3) for control  Female: 40.4% for cases, 49.0% for control | F1 score (SD): non-HAPI 0.81 (0.01); HAPI 0.86 (0.02)  Accuracy^e^ (SD): non-HAPI 0.85 (0.02); HAPI 0.88 (0.02)  AUC (SD): non-HAPI 0.92 (0.03); HAPI 0.94 (0.02)  Sensitivity (SD): non-HAPI 0.84 (0.02); HAPI 0.87 (0.03)  Specificity: non-HAPI 0.85 (0.02); HAPI 0.88 (0.02) |
| ML Sotoodeh (2020)^188^ | Yes ^108^ | USA; ICU; adult (≥16y); retrospective | ML; LR, RF, ANN | NS | NS | NS | 24457 | NS | F1 score (SD): 0.79 (0.02)  AUC (SD): 0.95 (0.01) |
| ML Sprigle (2020)^189^ | Yes ^89^ | USA; mobility related disabilities; NS; NS | ML; gradient boosting | NS | NS | Alzheimer’s disease; cerebral palsy; hemiplegia; multiple sclerosis; paraplegia/quadriplegia | 1252313 (NS, 6.9%) | Female: 54.1% | Sensitivity: 0.70  Specificity: 0.92 |
| ML SPURS (2019)^190^ | Yes ^78^ | NS; hospital, surgery; NS; retrospective | ML; LR | NS | Split sample; 70% training 30% testing | Age; female; ASA grade; BMI; Braden score; anemia; respiratory disease; hypertension | 269 | NS | Sensitivity: 0.40  Specificity: 0.70-0.95  PPV: 0.73  NPV: 0.08 |
| ML Su (2012)^191^ | Yes ^83 89 99 100 108^ | China; surgery; adult; prospective | ML; MTS (chosen method), LR, DT, SVM | NS | 4-fold CV; MTS model uses split sample 75% training 25% testing | Sex; age; weight; surgery type; body position during the operation; difference in temperature during surgery; surgical time (mins); air conditioning in operating room; number of electronic knives used in surgery | 168 (8) | Mean (SD) age: 65.4 (7.46) Female: 65.4% | F1 score: 0.38  F-score:  0.38 (MTS); 0.61 (SVM); 0.53 (DT); 0.67 (LR)  g-means: 0.82 (MTS); 0.81 (SVM); 0.7 (DT);  0.79 (LR)  Sensitivity: 0.76 (MTS); 0.67 (SVM); 0.50 (DT); 0.63 (LR)  Specificity: 0.89 (MTS); 0.95 (SVM); 0.98 (DT); 0.99 (LR) |
| ML Sun (2020)^192^ | Yes ^78^ | China; ICU; cancer patients; retrospective | ML; LR | NS | NS | Age; gender; diagnosis; cancer; anti-cancer therapy; Waterlow score; laboratory results; medications; length of stay; mechanical ventilation; APACHE score; blood purification | 486 | NS | Accuracy^e^: 0.83  Sensitivity: 0.66-0.81  Specificity: 0.78-0.96  AUC: 0.82-0.95 |
| ML Tang (2021)^193^ | Yes ^78^ | NS; surgery; NS; prospective | ML; LR | NS | NS | Braden score; preoperative fasting blood glucose level; emergency surgery; types of vasoactive drugs | 648 | NS | Sensitivity: 0.63  Specificity: 0.86  AUC: 0.74 |
| ML Vyas (2020)^194^ | Yes ^83 108^ | USA; ICU; adult (≥16y); retrospective | ML; XGBoost | NS | Split sample; 80% testing 20% testing | Mobility; activity; sensory perception; skin moisture; nutritional state; friction and shear | 13282 (NS, 16.8%) | NS | F1 score: 0.27  Accuracy^e^: 0.95  Sensitivity: 0.84  Specificity: 0.97  PPV: 0.87  NPV: 0.03 |
| ML Walther (2022)^195^ | Yes ^78 83 99^ | Germany; hospital; adult (≥19y); retrospective | ML; LR, BART, LASSO, RF | NS | 10-fold CV; split sample; 80% training 20% testing | Length of anesthesia; wards involved in care; admission reasons; ICU (with/without ventilation); age; sex; comorbidities | 149006 (4663) | Median age: 64  Female: 48.5% | Accuracy^e^: 0.52-0.55 Sensitivity: 0.04-0.10  Specificity: 1 PPV: 0.39-0.58  NPV: 0.98-0.99  AUC: 0.89-0.90 |
| ML Wang (2021)^196^ | Yes ^83^ | China; ICU; NS; retrospective | ML; RF, SVM, other | NS | Split sample; 67% testing 33% testing | NS | 246 (NS, 50%) | NS | NS |
| ML Waterlow 5-item (2000)^197^ | Yes ^102^ | UK; acute hospital; adult (>18y); retrospective | ML; LR, ANN | Follow-up 14 days | Split sample | NS | 422 (69) | Mean (SD) age: 64.8 (17.9) Female: 52.6% | NS |
| ML Xu (2022)^198^ | Yes ^78 83 99^ | China; ICU; adult; retrospective | ML; LR, DT, RF | NS | 5-fold CV; split sample; 70% training 30% testing | Reason for admission; clinical laboratory results; patients’ demographics; medical history; Braden scale | 618 (204) | NS | Accuracy^e^: 0.62-0.78 Sensitivity: 0.38-0.61 Specificity: 0.80-0.89  PPV: 0.54-0.65  NPV: 0.75-0.82  AUC: 0.72-0.88  F1 score: 0.49-0.62 |
| ML Yang (2019)^199^ | Yes ^89 99 100^ | China; tumour hospital; adult; prospective | ML; DT | NS | NS | Braden; inability to turn over; existing/potential damage to the skin; special circumstances | 611 (46) | Mean (SD) age: 61.8 (13.6) Female: 35.5% | Sensitivity: 0.848  Specificity: 0.774  AUC: 0.84 (95% CI 0.81- 0.87) |
| Norton (1962)^200^ | Yes ^79 82 84 87 91 96 97 103 107 109 110^ | UK; hospital; elderly; NS | Statistical; clinical expertise | NS | NS | Physical condition; mental status; activity; mobility; continence | 250 (60) | NS | NS |
| Norton modified by Bale (1995)^201^ | Yes ^82 84 87 110^ | UK; hospice/palliative care; adult; prospective | Statistical; NS | Group A mean (SD) days: 12 (6) Group B mean (SD) days: 13 (5) | NS | General physical condition; mobility; nutritional status; pain continence; special risk factors | 240 (38) | Mean age: 67 Women group A: 45% (group A), 59% (group B) | NS |
| Norton modified by Bienstein (1991)^202^ | Yes ^82 84 103^ | Germany; NS; adult; NS | Statistical; NS | NS | NS | Skin condition; cooperation/motivation; physical condition; additional diseases; mental state; incontinence; activity; mobility; age | NS | NS | NS |
| Norton modified by Ek 87 (1987)^203^ | Yes ^84^ | Sweden; long-term medical ward; adult; NS | Statistical; NS | NS | NS | NS | 367 (55) | Age range: 21-101 | NS |
| Norton modified by Ek 89 (1989)^204^ | No | Sweden; long-term medical ward; NS; NS | Statistical; NS | NS | NS | NS | NS | Mean age: 81 (women), 78 (men) | NS |
| Norton modified by Ek 91 (1991)^205^ | No | Sweden; long-term medical ward; NS; NS | Statistical; NS | NS | NS | NS | NS | Mean age: 80 | NS |
| Norton modified by Ek 97 (1997)^206^ | Yes ^87 110^ | Sweden; NS; NS; NS | NS; NS | NS | NS | NS | NS | NS | NS |
| Norton modified by Stotts (1988)^207^ | Yes ^82^ | USA; surgical (cardiovascular surgery and neurosurgery); adult (>18y); prospective | Statistical; NS | Follow-up to 3 weeks/to discharge | NS | Same items as the standard Norton scale with clarification regarding specific operational definitions | 387 (67) | Mean (range) age: 53 (17-86) Female: 47% | NS |
| Norton scale simplified (1998)^208^ | Yes ^102^ | Switzerland; acute hospital inpatient; adult (>16y); retrospective | Statistical; Cox regression | Follow-up duration/ length of stay: 9 days | NS | NS | 2373 (245) | Mean (SD) age: 63 (19) Female: 49% | NS |
| NOVA-4 | Yes ^84^ | NS; NS; adult; NS | Statistical; NS | NS | NS | NS | 187 | NS | NS |
| NPRU (1991)^209^ | No | NS | Statistical; NS | NS | NS | NS | NS | NS | NS |
| PARA (1994)^210^ | No | UK; ICU; NS; NS | Statistical; NS | NS | NS | NS | NS | NS | NS |
| prePURSE study tool (2006)^211^ | Yes ^102^ | Netherlands; acute hospital inpatient; adult (>18y); prospective | Statistical; LR | Follow-up 12 weeks | Resampling | NS | 1229 (121) | Mean (SD) age: 60.1 (16.7) Female: 54.8% | O/E ratio: 1.0 (95% CI 0.84–1.19)  C-statistic: 0.71 (95% CI 0.66–0.75) |
| Pressure Sore Predictor Scale (1987)^212^ | Yes ^84 110^ | UK; orthopaedic surgery and trauma; NS; NS | Statistical; NS | 3 weeks | NS | NS | 712 (in Lowthian87); 1244 (in '89) | NS | NS |
| Ramstadius (2000)^213^ | Yes ^82 85 91 92 94^ | Australia; hospital; NS; NS | Statistical; NS | NS | NS | Mobility; ability to independently reposition themselves; age; medication; skin integrity; temperature; decreased blood volume; dyspnoea and presence of an existing PI | NS | NS | NS |
| Risk Assessment Pressure Sore (RAPS) (2002)^214^ | Yes ^82 84 107 109 110^ | Sweden; hospital inpatient (medical, surgical, infection, orthopaedic, rehabilitation or geriatric ward); adult (>16y); prospective | Statistical; NS | Maximum follow-up 12 weeks; 50% of patients had ≤8 days follow-up | NS | NS | 488 (62) | Mean (SD) age: 70 (14) Female: 50% | NS |
| Rose/Cohen ICU model (2006)^215 216^ | Yes ^102^ | Canada; ICU; NS; prospective | Statistical; NS | Follow-up 8 days | NS | NS | 111 | NS | NS |
| S.S. (Suriada-Sanada) scale (2008)^217^ | Yes ^84 102 107 109^ | Indonesia; ICU; adult (≥18y); prospective | Statistical; LR, discriminant analysis | Mean follow-up 5.9 (SD 3.49) days | NS | Interface pressure; body temperature; cigarette smoking | 105 (35) | Mean (SD) age: 48.6 (17.5) Female: 31.4% | C-statistic: 0.89 (95% CI 0.83–0.93) |
| SCIPUS (1996)^218^ | No | USA; long-term care for SCI; adult; NS | Statistical; NS | NS | NS | Level of activity; levels of mobility; severity of SCI; complete SCI; autonomic dysreflexia or severe spasticity; urine incontinence or constantly moist; pre-existing conditions (age, tobacco use/smoking, pulmonary disease, cardiac disease or glucose >110mg dl^-1^, renal disease, impaired cognitive function); residence in a nursing home or hospital; nutrition (albumin <3.4 or total protein <6.4, anaemia haematocrit $\leq$36%) | 176 | NS | NS |
| SCIPUS-A (1999)^219^ | No | USA; SCI inpatient; adult; NS | Statistical; NS | NS | NS | Extent of paralysis; incontinence (moisture, continence); mobility; level of activity; nutrition (serum creatinine, albumin); pre-existing conditions (pulmonary disease) | 226 | NS | NS |
| Shannon (1982)^220^ | No | NS | Statistical; NS | NS | NS | NS | NS | NS | NS |
| Stratheden (1996)^221^ | No | UK; NS; elderly; NS | Statistical; NS | NS | NS | NS | NS | NS | NS |
| Sunderland scale (1995)^222^ | Yes ^84 107 109^ | UK; ICU; any; NS | Statistical; NS | NS | NS | NS | 15 | NS | NS |
| Surgical ICU risk assessment scale (2010)^223^ | Yes ^102^ | USA; surgical ICU; NS; prospective | Statistical; LR | NS | NS | NS | 369 (88) | Mean (SD) age years: 58.3 (19.3) Female: 43.6% | NS |
| The Northern Hospital Pressure Ulcer Prevention Plan (TNH-PUPP) model (2011)^224^ | Yes ^82 102^ | Australia; acute hospital inpatient; NS; retrospective | Statistical; LR | Mean follow-up 15.42 (SD 22.29) days | NS | NS | 342 (67) | Mean (SD) age years: 63 (19.8) Female: 54.3% | C-statistic: 0.86 (95% CI 0.81–0.90) |
| Vascular surgery PI risk score (2014)^225^ | Yes ^102^ | USA; cardiovascular surgery; adult; retrospective | Statistical; LR | Mean follow-up 7.08 (SD 7.44) days | Resampling | NS | 849 (101) | Mean (SD) age, years: 68.7 (13.0) | C-statistic: 0.85 (95% CI 0.81–0.89) |
| Waterlow (1985)^226^ | Yes ^82 84 85 90-92 94 96 97 103 104 107 109 110^ | UK; orthopaedic/generic; NS; NS | Statistical; NS | NS | NS | BMI; assessment of the skin; gender; age; malnutrition; incontinence; mobility; tissue malnutrition; neurological deficits; major surgery or trauma; medication | NS | NS | NS |
| Waterlow 12-item (1999)^227^ | Yes ^90^ | UK; hospital (acute and rehabilitation); elderly; NS | Statistical; NS | NS | NS | Skin type; mobility; poor nutrition; build; continence; appetite; medication; age; neurological deficit (including severe rheumatoid arthritis); major surgery; sex | NS | NS | NS |
| Waterlow scale simplified (2002)^228^ | Yes ^102^ | UK; acute hospital inpatient; adult (>65y); prospective | Statistical; LR | Follow-up 14 days | NS | NS | 213 (47) | Mean (SD) age years: 76.7 (8.1) | NS |
| Watkinson (1997)^229^ | Yes ^84^ | UK; NS; adult; NS | Statistical; NS | NS | NS | NS | 185 | NS | NS |

ANN – artificial neural network; ASA – American Society of Anaesthesiologists; AUC – area under curve; AUPRC – area under the precision-recall curve; BN – Bayesian network; BMI – body mass index; CANTRIP – reCurrent Additive Network for Temporal RIsk Prediction; CV – cross-validation; CVD – cardiovascular disease; DT – decision tree; EBM – explainable boosting machine; EN – elastic net; FIM – functional independence measure; GB – gradient boosting; ICU – intensive care unit; KNN – k-nearest neighbours; LDA – linear discriminant analysis; LOS – abbreviation not given by review authors; LR – logistic regression; LSTM – long short-term memory; ML – machine learning; MLP – multilayer perception; MTS – Mahalanobis-Taguchi system; NB – naïve Bayes; NN – neural network; NS – not stated; O/E – observed vs expected; OMPI – oral mucosal pressure injury; RF – random forest; RF – random forest; SCI – spinal cord injury; SD – standard deviation; SVM – support vector machine.

^a^Interrater agreement study.

^b^Expert opinion on clinical utility, reliability, usability, etc.

^c^HAPIs classified as stage 1 to 4, deep-tissue injury, or unstageable.

^d^HAPIs classified as stage 2 to 4, deep-tissue injury, or unstageable.

^e^Measure of accuracy not specified.

#### Table S5. Prognostic model external validation study characteristics

| **Name of model/tool (publication year)** | **Model validation publication author** | **Model validation considered in reviews** | **Country; setting; patients; data source (validation population)** | **N patients (n events)** | **Recalibration of original model** | **Patient characteristics** | **Performance metrics** |
| --- | --- | --- | --- | --- | --- | --- | --- |
| Berlowitz 11-item (1996)^115^ | Berlowitz (1996)^a,115^ | ^102^ | NS; long-term care; NS; retrospective | 17946 (556) | NS | Mean (SD) age: 71.0 (11.4) Female: 3% | O/E ratio: 0.97 (95% CI 0.90–1.06) |
| Berlowitz MDS risk adjustment model (2001)^116^ | Berlowitz (2001) ^230^ | ^102^ | NS; long-term care; NS; retrospective | 13457 (608) | No | NS | C-statistic: 0.73 (95% CI 0.71–0.75)  O/E ratio: 0.91 (95% CI 0.84–0.98) |
| Compton ICU model (2008)^127^ | Compton (2008)^a,127^ | ^102^ | NS; specific acute care; NS; retrospective | 329 (56) | NS | Median age: 67  Female: 45% | C-statistic: 0.80 (95% CI 0.73–0.86) |
| ML interRAI PURS (2010)^169^ | Poss (2010)^a,169^ | ^102^ | NS; long-term care; NS; retrospective | 13062 (1267) | NS | NS | C-statistic: 0.61 (95% CI 0.59–0.62) |
| ML interRAI PURS (2010)^169^ | Poss (2010)^a,169^ | ^102^ | NS; long-term care; NS; retrospective | 73183 (1903) | NS | NS | C-statistic: 0.63 (95% CI 0.62–0.64) |
| ML Ladios-Martin (2020)^231^ | Ladios-Martin (2020)^a,231^ | ^99^ | NS; ICU; NS; NS | NS | NS | NS | NS |
| prePURSE study tool (2006)^211^ | Schoonhoven (2005)^b^ | ^102^ | NS; acute care hospital; NS; prospective | 1440 | NS | NS | NS |
| S.S. (Suriada-Sanada) scale (2008)^217^ | Suriadi 2008^a,217^ | ^102^ | NS; acute care hospital; NS; prospective | 253 (7) | NS | Mean (SD) age: 51.3 (19.4) Female: 37.5% | NS |
| The Northern Hospital Pressure Ulcer Prevention Plan (TNH-PUPP) model (2011)^224^ | Page 2011^a,224^ | ^102^ | NS; acute care hospital; NS; prospective | 165 (7) | NS | Mean (SD) age: 68 (18.4) Female: 47% | C-statistic: 0.90 (95% CI 0.66–0.98) |

ICU – intensive care unit; ML – machine learning; NS – not stated; O/E – observed versus expected; SD – standard deviation. ^a^ Model appears to be externally validated in the same publication as it was developed; ^b^ Model validation citation unclear; year for model development given as 2006 and year for model validation given as 2005.

#### Table S6. Table of Predictors, by tool (predictors were reported for 66 tools), ascertained at review level except in the case of discrepancies between reviews

| **Tool name** | **General Health** | **Mental Status** | **Activity** | **Mobility** | **Ability to ambulate** | **Body position** | **Continence** | **Nutrition** | **Pre-disposing conditions** | **Age** | **Gender** | **Ethnicity or place of birth** | **Body** | **Skin** | **Isolation** | **Braden score** | **Receiving medical Tx/Rx** | **Laboratory values** | **Surgery duration** | **Length of stay** | **Pressure injury** | **Medical unit, ward, visit** | **Friction, shear, pressure** | **Pain** | **Hygiene** | **Smoking** | **Norton or Waterlow score** | **'Special'** (not explained) |
| --- | --- | --- | --- | --- | --- | --- | --- | --- | --- | --- | --- | --- | --- | --- | --- | --- | --- | --- | --- | --- | --- | --- | --- | --- | --- | --- | --- | --- |
| **No. of tools predictor appears in** | 18 | 21 | 21 | 28 | 6 | 3 | 23 | 22 | 32 | 33 | 21 | 5 | 22 | 21 | 2 | 14 | 30 | 27 | 7 | 8 | 8 | 6 | 5 | 3 | 3 | 2 | 2 | 2 |
| Abruzzese^111^ | Y | Y | Y | Y |  |  | Y | **Y x3** | Y |  |  |  |  |  |  |  |  |  |  |  |  |  |  |  |  |  |  |  |
| Braden^117^ |  | Y | Y | Y |  |  |  | Y |  |  |  |  |  | Y |  |  |  |  |  |  |  |  | Y |  |  |  |  |  |
| Braden - modified by Choi & Song^120 121^ | Y | Y | Y | Y |  |  |  | Y |  |  |  |  |  | Y |  |  | **Y x3** |  |  |  |  |  | Y |  |  |  |  |  |
| Braden - modified by Halfens / 4-factor model^122^ |  | Y |  |  |  |  |  |  |  | Y |  |  |  | Y |  |  |  |  |  |  |  |  | Y |  |  |  |  |  |
| Cubbin & Jackson^128^ | **Y x2** | Y |  | Y |  |  | Y | Y |  | Y |  |  | Y | Y |  |  |  |  |  |  |  |  |  |  | Y |  |  |  |
| Cubbin & Jackson (revised) "Jackson-Cubbin"^129^ | **Y x3** | Y |  | Y |  |  | Y | Y | Y | Y |  |  | Y | Y |  |  |  |  |  |  |  |  |  |  | Y |  |  |  |
| Douglas - based on Norton^131^ | Y |  | Y |  |  |  | Y |  | **Y x2** |  |  |  |  |  |  |  | **Y x2** |  |  |  |  |  |  | Y |  |  |  |  |
| Finnish risk assessment scale^14^ |  | **Y x2** | Y | Y |  |  | Y | **Y x2** |  |  |  |  |  | Y |  |  | **Y x2** |  |  |  |  |  |  |  |  |  |  |  |
| Gosnell^137^ |  | Y |  | Y | Y |  | Y | Y |  |  |  |  |  |  |  |  |  |  |  |  |  |  |  |  |  |  |  |  |
| Hatanaka^138^ |  |  |  |  |  |  |  |  |  | Y | Y |  |  |  |  |  |  | **Y x3** |  |  |  |  |  |  |  |  |  |  |
| Hyun^168^ |  |  |  |  |  |  |  |  | Y | Y | Y |  | Y |  | Y | Y | **Y x4** |  |  |  |  |  |  |  |  |  |  |  |
| Knoll Decubitus Ulcer Potential Scale^140^ | Y | Y | Y | Y |  |  | Y | **Y x2** | Y |  |  |  |  |  |  |  |  |  |  |  |  |  |  |  |  |  |  |  |
| ML Alderden [1]^143^ | Y |  |  |  |  |  |  |  | Y | Y |  |  | **Y x2** |  |  |  |  | **Y x6** | Y |  |  |  |  |  |  |  |  |  |
| ML Alderden [2]^144^ |  |  |  |  |  |  |  |  |  |  |  |  |  | Y |  | Y | Y | **Y x2** | Y | Y |  |  |  |  |  |  |  |  |
| ML Cai^147^ |  |  |  |  |  |  |  |  | Y | Y | Y |  | Y |  |  |  |  |  | **Y x2** |  |  |  |  |  |  |  |  |  |
| ML Chen [1]^149^ |  |  |  |  |  |  |  |  | Y | Y |  |  |  |  |  |  | Y |  | Y |  |  |  |  |  |  |  |  |  |
| ML Chen [2]^150^ |  |  |  |  |  |  |  |  | Y | Y |  |  | Y |  |  |  |  | **Y x5** |  |  |  |  |  |  |  | Y |  |  |
| ML Cheng [1]^152^ |  | **Y x2** | Y |  |  | Y |  |  |  | Y |  |  |  | Y |  |  | **Yx3** | Y |  |  | Y |  |  |  |  |  |  |  |
| ML Cho^154^ |  |  |  |  |  |  | **Y x3** | Y | **Y x5** | Y | Y |  | Y | Y |  |  | **Y x6** | **Y x3** |  | Y |  | Y |  |  |  |  |  |  |
| ML Choi^155^ |  |  |  |  |  |  |  |  |  |  |  |  | Y |  |  |  | **Y x4** | **Y x2** |  |  |  |  |  |  |  |  |  |  |
| ML Cichosz^156^ |  | Y | Y | Y | Y |  |  |  |  |  | Y |  |  |  |  |  |  |  |  |  |  |  |  |  |  |  |  |  |
| ML Cramer^157^ |  | Y |  |  |  |  |  |  |  |  |  |  |  |  |  |  | **Y x2** | **Y x3** |  |  | Y | **Y x3** |  |  |  |  |  |  |
| ML Delparte^158^ |  |  |  | Y | Y |  | Y |  |  |  |  |  |  |  |  |  |  |  |  |  | Y |  |  |  |  |  |  |  |
| ML Deng [1]^159^ |  |  |  |  |  |  | Y |  |  | Y |  |  |  |  |  | Y |  | Y |  |  |  |  |  |  |  |  |  |  |
| ML Deng [2]^160^ |  |  |  |  |  |  | Y |  |  | Y |  |  |  |  |  | Y | Y | **Y x2** |  | Y |  |  |  |  |  |  |  |  |
| ML Deschepper^161^ |  |  |  |  |  |  |  |  | **Y x2** | Y | Y |  | **Y x4** |  |  | Y |  | Y |  |  |  |  |  |  |  |  |  |  |
| ML Do^162^ | Y |  | Y |  |  | Y |  |  |  |  |  |  | **Y x2** |  |  | Y |  | **Y x4** |  |  |  |  |  |  |  |  |  |  |
| ML Eshetie^163^ |  |  |  | Y |  |  | Y |  |  |  |  |  |  |  |  |  | **Y x2** |  |  |  | Y |  |  |  |  |  |  |  |
| ML Gao^164^ |  |  |  |  |  |  |  |  |  |  |  |  | Y |  |  |  | **Y x4** |  | Y |  |  |  |  |  |  |  |  |  |
| ML Hou^166^ |  | Y | Y | Y |  |  | Y | Y |  | Y |  |  |  | Y |  |  |  | **Y x2** |  |  |  |  |  |  |  |  |  |  |
| ML Hu^167^ |  | **Y x4** |  | Y |  |  |  |  |  |  |  |  |  | **Y x3** |  |  |  | **Y x4** |  |  |  |  |  |  |  |  |  |  |
| ML Hyun^168^ |  |  |  |  |  |  |  |  | Y | Y | Y |  | Y |  | Y | Y | **Y x3** |  |  | Y |  |  |  |  |  |  |  |  |
| ML James^170^ |  |  |  |  |  |  |  |  |  | Y | Y |  | **Y x2** |  |  |  | **Y x2** | **Y x2** |  | Y |  |  |  |  |  |  |  |  |
| ML Kaewprag [1]^172^ |  |  |  |  |  |  |  |  | **Y x 61** |  |  |  |  |  |  | Y | **Y x 18** |  |  |  |  |  |  |  |  |  |  |  |
| ML Kaewprag [2]^173^ |  |  |  |  |  |  |  |  | **Y x 61** |  |  |  |  |  |  | Y | **Y x 18** |  |  |  |  |  |  |  |  |  |  |  |
| ML Kim [1]^174^ |  |  |  |  |  |  |  |  | Y | Y | Y | Y |  |  |  |  | **Y x2** | Y |  |  |  | Y |  |  |  |  |  |  |
| ML Kim [2]^232^ |  |  |  |  | Y |  |  | Y | Y |  |  |  |  |  |  |  | **Y x2** |  |  |  |  |  |  |  |  |  |  |  |
| ML Ladios-Martin^175^ | Y |  |  |  |  |  |  | Y | Y | Y | Y | Y |  |  |  |  | **Y x3** | Y |  |  | Y |  |  |  |  |  |  |  |
| ML Li [1]^178^ |  |  | Y | Y |  |  | Y |  | Y |  |  |  |  | Y |  |  |  |  |  |  | Y |  |  |  |  |  |  |  |
| ML Li [2]^179^ |  |  | Y | Y |  |  | Y | Y |  |  |  |  | Y | Y |  |  |  |  |  |  |  | Y |  |  |  |  |  |  |
| ML Nakagami^180^ |  | Y |  |  | **Y x5** |  |  | Y | **Y x3** | Y | Y |  |  | Y |  |  |  |  |  |  |  | Y |  | Y |  |  |  |  |
| ML Park^182^ |  | **Y x2** |  | Y |  |  |  | Y |  |  |  |  |  | Y |  |  | **Y x3** | Y |  |  |  |  |  |  | Y |  |  |  |
| ML Setoguchi^183^ |  |  | Y |  |  |  |  |  |  |  |  |  | Y |  |  |  |  |  | Y |  |  |  |  |  |  |  |  |  |
| ML Shui^184^ |  |  |  |  |  |  |  |  |  | Y |  |  | Y |  |  | Y | **Y x2** | Y |  |  |  |  |  |  |  |  |  |  |
| ML Šín^185^ |  |  |  |  |  |  |  | **Y x2** |  | Y | Y | Y | **Y x2** |  |  |  |  | **Y x3** |  | Y |  |  |  |  |  |  |  |  |
| ML Song [1]^186^ | Y |  | Y | **Y x2** |  |  | Y | **Y x3** | **Y x2** | Y | Y |  | **Y x2** |  |  |  | Y | **Y x2** |  | Y |  |  |  |  |  |  | Y |  |
| ML Song [2]^187^ |  | Y | Y |  | Y |  |  |  | **Y x5** | Y | Y | Y |  |  |  |  |  | **Y x9** |  |  | Y |  |  | Y |  |  |  |  |
| ML Sprigle^189^ |  |  |  |  |  |  |  |  | **Y x5** |  |  |  |  |  |  |  |  |  |  |  |  |  |  |  |  |  |  |  |
| ML SPURS^190^ | Y |  |  |  |  |  |  |  | **Y x3** | Y | Y |  | Y |  |  | Y |  |  |  |  |  |  |  |  |  |  |  |  |
| ML Su^191^ |  |  |  |  |  | Y |  |  |  | Y | Y |  | **Y x2** |  |  |  | **Y x3** |  | Y |  |  |  |  |  |  |  |  |  |
| ML Sun^192^ | Y |  |  |  |  |  |  |  | **Y x2** | Y | Y |  |  |  |  |  | **Y x4** | Y |  | Y |  |  |  |  |  |  | Y |  |
| ML Tang^193^ |  |  |  |  |  |  |  |  |  |  |  |  |  |  |  | Y | **Y x2** | Y |  |  |  |  |  |  |  |  |  |  |
| ML Vyas^194^ |  | Y | Y | Y |  |  |  | Y |  |  |  |  |  | Y |  |  |  |  |  |  |  |  | Y |  |  |  |  |  |
| ML Walther^195^ |  |  |  |  |  |  |  |  | Y | Y | Y |  |  |  |  |  | **Y x2** |  |  |  |  | Y |  |  |  |  |  |  |
| ML Xu^198^ |  |  |  |  |  |  |  |  | Y | Y | Y | Y |  |  |  | Y |  | Y |  |  |  |  |  |  |  |  |  |  |
| ML Yang^199^ |  |  |  | Y |  |  |  |  |  |  |  |  |  | Y |  | Y |  |  |  |  |  |  |  |  |  |  |  | Y |
| Norton^200^ | Y | Y | Y | Y |  |  | Y |  |  |  |  |  |  |  |  |  |  |  |  |  |  |  |  |  |  |  |  |  |
| Norton modified by Bale^201^ | Y |  |  | Y |  |  | Y | Y |  |  |  |  |  |  |  |  |  |  |  |  |  |  |  |  |  |  |  | Y |
| Norton modified by Bienstein^202^ | Y | **Y x2** | Y | Y |  |  | Y |  | Y | Y |  |  |  | Y |  |  |  |  |  |  |  |  |  |  |  |  |  |  |
| Norton modified by Stotts^207^ | Y | Y | Y | Y |  |  | Y |  |  |  |  |  |  |  |  |  |  |  |  |  |  |  |  |  |  |  |  |  |
| Ramstadius PI risk assessment^213^ | Y |  |  | **Y x2** |  |  |  |  | Y | Y |  |  |  | Y |  |  | Y | Y |  |  | Y |  |  |  |  |  |  |  |
| S.S. (Suriada-Sanada) scale^217^ | Y |  |  |  |  |  |  |  |  |  |  |  |  |  |  |  |  |  |  |  |  |  | Y |  |  | Y |  |  |
| SCIPUS^218^ |  |  | Y | Y |  |  |  |  | **Y x3** |  |  |  |  |  |  |  |  |  |  |  |  |  |  |  |  |  |  |  |
| SCIPUS-A^219^ |  |  | Y | Y |  |  | **Y x2** | Y | **Y x2** |  |  |  |  |  |  |  |  | **Y x2** |  |  |  |  |  |  |  |  |  |  |
| Waterlow^226^ |  |  |  | Y |  |  | Y | **Y x2** | Y | Y | Y |  | Y | Y |  |  | **Y x2** |  |  |  |  |  |  |  |  |  |  |  |
| Waterlow 12-item^227^ |  |  |  | Y |  |  | Y | **Y x2** | Y | Y | Y |  | Y | Y |  |  | **Y x2** |  |  |  |  |  |  |  |  |  |  |  |
